# Genome-wide association meta-analysis of atrophic gastritis reveals anti–*H. pylori* IgG serostatus-dependent associations and genetic overlap with gastric cancer

**DOI:** 10.64898/2026.11.10.26367681

**Authors:** Tomoki Kyosaka, Akira Narita, Akimitsu Miyake, Yurii Kotsar, Takafumi Ojima, Shinichi Namba, Atsushi Shimizu, Yoichi Sutoh, Masahiro Nakatochi, Yuriko N. Koyanagi, Yunye He, Taiki Yamaji, Yu Sasaki, Takeshi Nishiyama, Asahi Hishida, Naoyuki Takashima, Yumiko Kasugai, Satoshi Makino, Matsuyuki Shirota, Jun Takayama, Chinatsu Gocho, Tomohisa Suzuki, Aye Ko Ko Minn, Kyoga Hiraide, Takamitsu Funayama, Yayoi Otsuka-Yamasaki, Kozo Tanno, Yasuhiko Abe, Osamu Nakajima, Sayaka Yamamoto, Hidemi Ito, Chizu Tanikawa, Masaru Koido, Shiori Nakano, Norie Sawada, Kenji Wakai, Atsushi Hozawa, Kengo Kinoshita, Hiroshi Okuda, Jerzy K Kulski, Keitaro Matsuo, Yoichiro Kamatani, Koichi Matsuda, Motoki Iwasaki, Yoshiyuki Ueno, Yukinori Okada, Shinichi Kuriyama, Motomichi Matsuzaki, Gen Tamiya

**Author notes:** Integrative Genomics and Predictive Medicine, Department of Integrative Genomics, Tohoku Medical Megabank Organization, Tohoku University, Sendai, Japan; One Health Lab (Endowed Research Division), Tohoku Medical Megabank Organization, Tohoku University, Sendai, Japan (Atsushi Shimizu). Corresponding authors: Akira Narita and Gen Tamiya.

## Abstract

**Background:** Atrophic gastritis (AG), characterized by chronic inflammation of the gastric mucosa and glandular atrophy, is an established precursor of gastric cancer (GC) and contributes to iron-deficiency anemia, vitamin B12 deficiency, and pernicious anemia. Although *Helicobacter pylori* infection is a major environmental determinant, not all infected individuals develop AG or GC, suggesting that host genetic factors influence susceptibility to AG and severe atrophic gastritis (SAG). Previous genome-wide association studies (GWAS) of AG have been limited in size and have not adequately compared anti-*H. pylori* IgG serostatus strata, leaving potential genetic heterogeneity unresolved.

**Methods:** We performed GWAS meta-analyses of AG and SAG, stratified by anti-*H. pylori* IgG serostatus, involving 214,747 Japanese participants: 75,074 in the discovery phase and 139,673 in the replication phase. Subsequent analyses included genotype-by-serostatus interactions, HLA allele and ABO blood group associations, genome-wide genetic correlations, multi-trait analysis, local genetic correlations, and structural equation modeling. In addition, cross-ancestry meta-analyses were performed using chronic gastritis GWAS data from FinnGen, UK Biobank, and the Million Veteran Program, comprising 1,357,053 participants.

**Results:** Across six Japanese GWAS meta-analyses, we identified genome-wide significant associations at 17 genomic regions. Strong associations with seropositive AG/SAG were observed at previously reported GC susceptibility loci, including *PSCA* and *MUC1*/*MTX1*, whereas the MHC class II region showed predominant associations with seronegative AG/SAG. *FAP*, *CCR6*, and *ATP4A* were also found to be associated with seronegative SAG. The identified regions included previously reported GC susceptibility regions *MUC1*/*MTX1*, *PRKAA1*/*PTGER4*, *PSCA*, *ABO*, and MHC. Associations at *PSCA* and *MUC1*/*MTX1* were replicated at a genome-wide significance level, supporting a shared genetic basis between AG/SAG and GC. Additional AG-associated candidate regions included *BMP2*, *CCDC80*, *LRRC34*, *CPEB4*, *PGC*/*FRS3*, and *LMTK2*, implicating epithelial regulation, immune responses, transcriptional regulation, and differentiation. A positive genotype-by-serostatus interaction for SAG was observed at *MUC1*/*MTX1*. HLA class II alleles, including *HLA-DRB1*15:01* and *HLA-DQB1*06:02*, showed associations with seronegative AG/SAG. The LDSC genetic correlation between AG and GC was 0.529, and the MTAG estimated Omega correlation was 0.694. Local genetic correlation analysis and structural equation modeling further supported shared genetic components. Cross-ancestry integration of seronegative SAG with external chronic gastritis GWAS data identified an additional association at *TPTEP1*.

**Conclusions:** Our findings characterize the genetic heterogeneity of serologically defined AG/SAG according to anti-*H. pylori* IgG serostatus. They highlight epithelial and mucus barrier-related pathways in seropositive AG/SAG and immune-related pathways in seronegative cases, while also providing evidence of a shared genetic basis between AG and GC. Together, these results clarify the genetic basis of a GC precursor and are consistent with a continuum from chronic inflammation through atrophy to carcinogenesis. These insights may inform prevention strategies and personalized management of the diverse health consequences of AG.

## Introduction

Atrophic gastritis (AG) is characterized by chronic inflammation of the gastric mucosa and atrophy of the gastric glands. In Correa’s multistep model of gastric carcinogenesis, chronic gastritis, atrophy, intestinal metaplasia, dysplasia, and gastric cancer (GC) constitute a sequence of pathological changes [1–3]. In addition to its role as a GC precursor, AG contributes to iron-deficiency anemia, vitamin B12 deficiency, and pernicious anemia through reductions in gastric acid secretion and intrinsic factor production [3,4]. Understanding the genetic determinants of AG development and progression is therefore relevant not only to GC prevention but also to the management of nutritional deficiencies and anemia in aging populations.

The Kyoto Global Consensus Report classified *H. pylori* gastritis as an infectious disease and recognized *H. pylori* infection as a major environmental determinant of AG and GC [5,6]. However, not all infected individuals progress to AG, severe atrophic gastritis (SAG), or GC. This variation suggests the involvement of host genetic factors, immune responses, gastric mucosal barrier function, and acid secretion. A recent large-scale GWAS of anti-*H. pylori* IgG levels, conducted predominantly in Japanese populations, identified host genetic associations at HLA class II and innate immunity-related loci [7]. Previous GWAS of GC have identified susceptibility regions including *PSCA*, *MUC1*, *PRKAA1*, and *ABO* [8–11]. Nevertheless, the stages of the carcinogenic cascade at which these regions act, and the extent of their contribution to AG and SAG, remain incompletely understood. Smaller studies have suggested associations of *PSCA* and *MUC1* with GC and precancerous lesions [12,13]. However, previous AG GWAS have been limited in size and have not adequately compared anti-*H. pylori* IgG serostatus strata, potentially obscuring the genetic heterogeneity of AG/SAG.

To investigate the genetic susceptibility underlying AG development and progression, we performed large-scale GWAS meta-analyses of AG and SAG in Japanese populations, with stratification by anti-*H. pylori* IgG serostatus. These analyses were followed by post-GWAS analyses and cross-ancestry meta-analyses incorporating external chronic gastritis data. We aimed to characterize differences in host genetic effects according to serostatus and to clarify how AG fits genetically within the disease continuum leading to GC.

## Methods

### Study design and participants

We performed Japanese GWAS meta-analyses of AG and SAG, followed by post-GWAS statistical genetic analyses and cross-ancestry meta-analyses incorporating external chronic gastritis GWAS. The Japanese analyses involved 214,747 participants, including 75,074 in the discovery phase and 139,673 in the replication phase. The discovery GWAS used four datasets from the Tohoku Medical Megabank (TMM) Community-Based Cohort Study [14,15]. For the replication phase, data were obtained from the TMM Birth and Three-Generation Cohort Study [16], Yamagata Study [17], Japan Public Health Center-based Prospective Study (JPHC) [18], Japan Multi-Institutional Collaborative Cohort Study (J-MICC) [19], Hospital-based Epidemiologic Research Program at Aichi Cancer Center (HERPACC) [20], and BioBank Japan (BBJ) [21]. The discovery phase included 15,644 AG cases and 59,430 controls, and 4,517 SAG cases and 70,557 controls. The replication phase included 9,548 AG cases and 130,125 controls. The replication analysis of SAG excluded BBJ and included 2,303 cases and 47,661 controls. Details of cohort-specific characteristics and case-control counts are provided in **Supplementary Table 1**. The external chronic gastritis datasets comprised 1,357,053 participants from FinnGen [22], UK Biobank [23], and the Million Veteran Program (MVP) [24]. An overview of the analysis workflow is provided in **Figure 1**.

**Figure 1.**
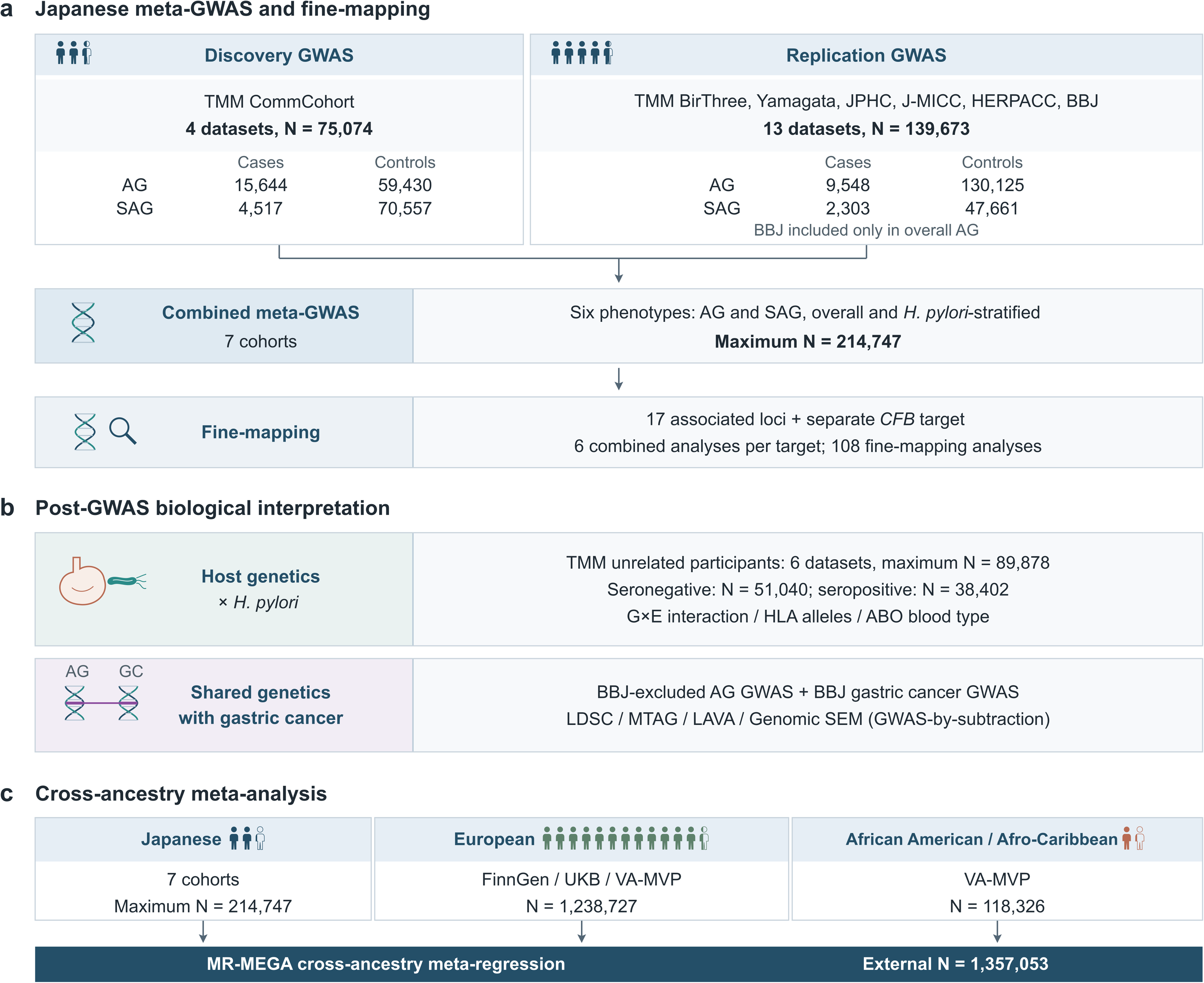
Study design and analytical framework of the AG/SAG GWAS meta-analysis. (a) Japanese GWAS discovery, replication, meta-analysis, and fine-mapping. The discovery stage included 75,074 participants from four TMM CommCohort datasets, and the replication stage included 139,673 participants from TMM BirThree, Yamagata Study, JPHC, J-MICC, HERPACC, and BBJ, yielding up to 214,747 participants from seven Japanese cohorts. Fixed-effect meta-analyses were conducted for overall AG, overall SAG, anti-*H. pylori* IgG-seropositive AG, anti-*H. pylori* IgG-seropositive SAG, anti-*H. pylori* IgG-seronegative AG, and anti-*H. pylori* IgG-seronegative SAG. Seropositive and seronegative strata were defined as anti-*H. pylori* IgG >3 U/mL and <=3 U/mL, respectively. These strata indicate seroreactivity and do not directly establish active infection. BBJ contributed medical diagnosis-based AG data and was included only in the overall AG analysis. FINEMAP was applied across the 17 associated loci and a separate *CFB* target; 35 genome-wide significant locus-analysis combinations are summarized in Supplementary Table 9. (b) Post-GWAS analyses. Post-GWAS analyses were performed in analysis-specific samples of unrelated TMM participants. The six TMM datasets contributing to the estimates in Supplementary Table 10 provided a maximum combined sample size of 89,878. The serostatus-stratified sample sets were restricted to participants retained after relatedness filtering in both the overall sample and the corresponding serostatus stratum. These sample sets included 51,040 seronegative participants (1,075 AG cases and 569 SAG cases) and 38,402 seropositive participants (13,375 AG cases and 3,567 SAG cases). For the shared-genetic analyses, Japanese AG GWAS summary statistics excluding BBJ were analyzed jointly with BBJ gastric cancer GWAS summary statistics to avoid sample overlap. (c) Cross-ancestry meta-analysis. Japanese AG/SAG GWAS results were integrated by MR-MEGA with chronic gastritis GWAS results from FinnGen, UK Biobank, and European and African American or Afro-Caribbean MVP datasets (external N = 1,357,053). Because the external phenotype was chronic gastritis rather than serologically defined AG/SAG, these analyses were interpreted as supportive evidence. AG, atrophic gastritis; BBJ, BioBank Japan; GC, gastric cancer; HLA, human leukocyte antigen; JPHC, Japan Public Health Center-based Prospective Study; J-MICC, Japan Multi-Institutional Collaborative Cohort Study; LAVA, Local Analysis of [co]Variant Association; LDSC, linkage disequilibrium score regression; MTAG, multi-trait analysis of GWAS; MVP, Million Veteran Program; SAG, severe atrophic gastritis; SEM, structural equation modeling; TMM, Tohoku Medical Megabank.

### Ethics

Informed consent was obtained from all participants. Approval was granted by the institutional review boards of Tohoku University for the TMM Project, the National Cancer Center for JPHC, Aichi Cancer Center for J-MICC and HERPACC, and Yamagata University for the Yamagata Study. For BBJ, approval was granted by the ethics committees of the Institute of Medical Science, the University of Tokyo, and the RIKEN Center for Integrative Medical Sciences.

### Phenotype definitions

In the Japanese cohorts other than BBJ, serum pepsinogen (PG) concentrations were used to define AG and SAG [25–27]. AG was defined as PG I ≤70 ng/mL and a PG I/II ratio ≤3, whereas SAG was defined as PG I ≤30 ng/mL and a PG I/II ratio ≤2. In BBJ, AG was defined by medical diagnosis rather than serum PG criteria. BBJ was included only in the overall AG analysis and was not included in the SAG or anti-*H. pylori* IgG serostatus-stratified analyses.

Anti-*H. pylori* IgG serostatus was determined using antibody levels [7,25–27]. Antibody levels were measured using a latex agglutination assay (LIA *H. pylori* latex Seiken) in TMM and HERPACC, an enzyme immunoassay (E-plate Eiken *H. pylori* antibody) in Yamagata Study and JPHC, and both assays in J-MICC. Participants with antibody concentrations >3 U/mL were classified as seropositive, and those with concentrations ≤3 U/mL as seronegative. Individuals with a history of GC were excluded.

### Genotyping and quality control

Genotyping was performed using Japonica Array version 2 and Japonica Array NEO [28], as well as Human660W-Quad, HumanOmniExpressExome, Infinium Asian Screening Array, and HumanCoreExome arrays from Illumina (San Diego, CA, USA). In TMM and Yamagata Study, imputation used a cross-imputed reference panel combining 1000 Genomes Project Phase 3 [29] and 3.5KJPNv2 data [30]. The 1000 Genomes Project Phase 3 reference panel was used in JPHC, J-MICC, HERPACC, and BBJ.

Quality control (QC) procedures were applied before and after imputation according to study-specific criteria. Pre-imputation QC considered sample call rate, sex discrepancies, heterozygosity, SNP call rate, minor allele frequency (MAF), and Hardy-Weinberg equilibrium (HWE). In TMM, phasing was performed using SHAPEIT v2 [31] and imputation using IMPUTE4 [32], followed by filtering based on imputation INFO score and MAF. In the other cohorts, QC, imputation, and GWAS were performed separately for each platform according to the respective analysis protocols. Detailed information on genotyping platforms, reference panels, software, and QC criteria is provided in **Supplementary Table 2**.

### GWAS and meta-analysis

GWAS were performed for six outcomes: overall AG, overall SAG, seronegative AG, seronegative SAG, seropositive AG, and seropositive SAG. Adjustments were made for age, sex, and principal components (PCs) 1-10. In the TMM cohorts, a generalized linear mixed model for binary traits (fastGWA-GLMM), implemented in GCTA version 1.94.0 beta [33,34], was used to account for cohort structure, including related individuals with PI-HAT >0.1875, by jointly modeling fixed effects and random genetic effects. Effect estimates from fastGWA-GLMM were obtained on the log-odds scale. In the other cohorts, GWAS were performed separately for each cohort and genotyping-array platform, with adjustment for age, sex, and PCs 1-10 according to study-specific protocols. REGENIE [35] was used for the three BBJ datasets, BBJ1-180K, BBJ1-12K, and BBJ2-42K. The models used for each dataset are described in **Supplementary Table 2**.

Per-allele log-odds effect estimates and their standard errors from individual cohorts and datasets were combined using inverse variance-weighted fixed-effect meta-analysis with METAL, release 2011-03-25 [36]. Meta-analyses were performed separately for the discovery phase, replication phase, and combined discovery and replication datasets. The BBJ GWAS of medically diagnosed AG was included only in the overall AG analysis. The genome-wide significance level was set at P-value <5 × 10^−8^. Locus annotation and visualization were performed using FUMA [37].

### Fine-mapping

Fine-mapping was performed using summary statistics for the 17 principal Japanese GWAS regions in each of the six combined GWAS analyses. A separate target at *CFB* was additionally evaluated in the same six analyses. Locus-specific fixed genomic windows of 100-350 kb containing the lead variant were analyzed using FINEMAP [38] to estimate posterior inclusion probabilities (PIPs) and credible sets under a Bayesian model. Linkage disequilibrium (LD) was estimated using the 1000 Genomes Project Phase 3 East Asian reference panel [29], and the maximum number of causal components was set to five. To assess correspondence with the GWAS findings, pairwise LD (*r*^2^) between prioritized variants and the GWAS lead variant was evaluated using the same reference panel.

### Post-GWAS analyses

To interpret the primary GWAS associations and evaluate genetic sharing between AG and GC, we performed genotype-by-serostatus interaction analyses, HLA allele and ABO blood group association analyses, and analyses of shared genetic architecture. Data for the interaction, HLA, and ABO analyses were obtained from unrelated TMM participants, defined by PI-HAT <0.1875. The six TMM datasets contributing to the estimates in Supplementary Table 10 provided a maximum combined sample size of 89,878. The serostatus-stratified sample sets were restricted to participants retained after relatedness filtering in both the overall sample and the corresponding serostatus stratum. These sample sets included 51,040 seronegative participants (1,075 AG cases and 569 SAG cases) and 38,402 seropositive participants (13,375 AG cases and 3,567 SAG cases).

### Genotype-by-anti-*H. pylori* IgG serostatus interaction analysis

Genotype, anti-*H. pylori* IgG serostatus, and their interaction were jointly evaluated in logistic regression models for AG or SAG. Serostatus was included as the environmental factor, with anti-*H. pylori* IgG levels >3 U/mL indicating positivity and ≤3 U/mL indicating negativity. This threshold reflects the classification of low antibody titers used to characterize anti-*H. pylori* IgG serostatus [27]. Genotype was represented by effect-allele dosage, and the models were adjusted for age, sex, and PCs 1-10. Analyses were performed using PLINK 2.0 [39]. The --glm interaction option tested the genotype-by-serostatus term, and model terms were specified using --parameters. Covariates, including serostatus, were variance-standardized; the conditional genetic main effect therefore represents the per-allele effect at mean serostatus. The interaction coefficient represents the change in the per-allele log-odds effect per one standard deviation of the binary serostatus indicator, rather than of antibody titer. We estimated marginal and serostatus-stratified associations across the six TMM datasets and calculated approximate model-derived joint ORs using environmental coefficients converted to binary serostatus contrasts within each dataset and pooled by inverse variance. The genome-wide significance threshold for interaction effects was set at P-value <5 × 10^−8^ for each outcome.

### HLA allele association analysis

To examine associations within the MHC at the HLA allele level, classical HLA alleles were imputed using DEEP*HLA [40]. This deep learning method estimates HLA allele dosages from pre-phased single-nucleotide variant data and has been evaluated in Japanese and European HLA reference panels. Pre-imputation QC was applied to variants in the MHC region (chr6:29,677,984-33,485,635; GRCh37/hg19), requiring call rate >0.97, MAF >0.01, and HWE P-value >1 × 10^−5^. The Pan-Asian HLA reference panel comprised 530 individuals and 1,060 haplotypes, including Singapore Chinese, pan-Asian Chinese, Indian, Malaysian, and HapMap Phase II JPT/CHB samples [41].

HLA typing at two- or four-digit resolution covered *HLA-A*, *HLA-B*, *HLA-C*, *HLA-DRB1*, *HLA-DQA1*, *HLA-DQB1*, *HLA-DPA1*, and *HLA-DPB1*, comprising 273 classical HLA alleles. Association analyses were performed using logistic regression in PLINK 2.0 [39], with AG or SAG as the outcome and imputed allele dosage, representing an expected copy number from 0 to 2, as the predictor. The models were adjusted for age, sex, and PCs 1-10. Alleles with an estimated frequency <0.01 were excluded, and the Bonferroni-corrected significance threshold was set at P-value <0.05/273 = 1.83 × 10^−4^.

### ABO blood group association analysis

We examined associations of ABO blood group with AG, SAG, and anti-*H. pylori* IgG seropositivity. Blood groups were inferred using rs505922 as a proxy for the O allele and rs8176746 and rs8176747 to distinguish A/B-related haplotypes, following reports in Japanese populations [42,43]. The rs505922 T allele is strongly correlated with the O allele. Inferred haplotypes were classified as O for OO, A for AO or AA, B for BO or BB, and AB for AB. Blood group O was used as the reference, with three indicator variables representing A, B, and AB.

Associations with AG or SAG were evaluated by logistic regression in the overall population and within serostatus strata, with adjustment for age, sex, and PCs 1-10. Odds ratios (ORs) were estimated relative to blood group O. A three-degree-of-freedom likelihood ratio test was used to assess overall blood group associations by comparing the three non-O categories with the O reference. Anti-*H. pylori* IgG seropositivity was also examined as an outcome. Bonferroni correction was applied separately across the 21 blood-group contrasts and seven omnibus tests.

### Shared genetic architecture of AG and GC

To assess genetic sharing between AG and GC, we performed complementary analyses using GWAS summary statistics. The summary statistics were harmonized to GRCh37/hg19, and strand-ambiguous or discordant variants, variants with MAF <0.01, and variants that could not be matched to the reference panel were excluded. Japanese AG meta-analysis statistics excluding BBJ were used to avoid sample overlap with the GC GWAS. GC summary statistics were obtained from PheWeb.jp for the Japanese BBJ population, comprising 7,921 cases and 159,201 controls [44].

#### Genome-wide genetic correlation

SNP heritability and genome-wide genetic correlations between AG/SAG and GC were estimated using LD score regression (LDSC) version 1.0.1 [45,46]. LD scores were based on the 1000 Genomes Project East Asian reference panel [29]. For each trait pair, we estimated SNP heritability, univariate LDSC intercepts, the cross-trait intercept, genetic correlation, its standard error (SE), and P-value. The LDSC intercepts were used to assess inflation attributable to population structure and sample overlap in relation to the inflation expected from polygenicity. Bonferroni correction was applied across the six AG/SAG–GC genetic correlations.

#### Multi-trait analysis

Multi-trait analysis of GWAS (MTAG) was used to jointly analyze GWAS summary statistics and estimate trait-specific effects by accounting for genetic covariance across traits [47]. Genome build, variant identifiers, and effect alleles were harmonized, and cross-trait genetic covariance was evaluated. Each of the six AG/SAG analyses was paired with GC, with detailed locus-level evaluation of the AG-GC analysis. MTAG-adjusted effect estimates, SEs, and P-values were compared with the primary GWAS findings and interpreted together with the LDSC results.

#### Local genetic correlation

Local genetic correlation analysis was performed using Local Analysis of [co]Variant Association (LAVA), version 0.1.0 [48], to determine whether genetic sharing between AG and GC was widespread or concentrated at specific loci. LAVA transforms marginal SNP effects within a region into joint effects using an LD reference and estimates local SNP heritability and cross-trait genetic correlation. The genome was partitioned into 2,495 approximately independent LD regions [49]. LD information was obtained from the 1000 Genomes Project Phase 3 East Asian reference panel [29]. Local SNP heritability was evaluated for both traits, and bivariate analysis was performed only in regions with nominally significant local heritability in both traits (univariate P-value < 0.05). Local genetic correlations, 95% CIs, and P-values were estimated, with the Bonferroni-corrected significance threshold set at P-value < 0.05/218 = 2.29 × 10^−4^ across all 218 eligible regions, including those with unavailable bivariate estimates.

In post hoc analyses, the MHC was divided into three broader, non-overlapping intervals corresponding to class I (chr6:29,500,000–31,498,002), class III (chr6:31,498,003–32,191,844), and class II (chr6:32,191,845–33,500,000), using GRCh37 coordinates. The class III boundaries were defined by the outer edges of *DDX39B* (formerly *BAT1*) and *NOTCH4*, following the gene-based definition of Xie et al. [50]. Local heritability and genetic correlation were re-estimated from the SNP-level inputs within each interval using the same analytical settings. Bonferroni correction was applied separately across these three regional tests (P-value < 0.05/3 = 0.0167).

#### Genomic structural equation modeling and GWAS-by-subtraction

The genetic covariance structure of AG and GC was evaluated using Genomic Structural Equation Modeling (Genomic SEM) [51]. A latent common-factor model was fitted using genetic covariance and sampling covariance matrices estimated by LDSC. As shown in Figure 3, AG was used as the anchor trait, with its loading on the shared factor fixed to 1 and its residual genetic variance constrained to 0. GC was modeled as a contribution from the shared factor plus a GC-specific residual. We estimated the loading of the shared factor on GC, its SE and P-value, and the proportion of GC genetic variance explained by the factor under the standardized variance parameterization. Model convergence was also assessed.

GWAS-by-subtraction analyses were performed using the Genomic SEM framework [52]. SNP effects were decomposed into a shared component and a GC-specific component remaining after subtraction of the component shared with AG, using a two-trait model based on AG and GC summary statistics. The component GWAS results were evaluated for genome-wide significance, regional associations at principal loci, and overlap with AG/GC-associated regions.

### Cross-ancestry meta-analysis

Japanese AG/SAG GWAS summary statistics were compared and integrated with chronic gastritis GWAS from FinnGen, UK Biobank, and MVP. The external datasets included 436,226 participants from FinnGen, comprising 12,441 cases and 423,785 controls; 361,194 from UK Biobank, with 1,790 cases; 441,307 MVP European participants, with 2,236 cases; and 118,326 MVP African American or Afro-Caribbean participants, with 886 cases. In total, these datasets comprised 1,357,053 participants.

Cross-ancestry meta-analysis was performed using MR-MEGA v0.2 [53]. Ancestry PCs derived from differences in mean allele frequencies between datasets were incorporated into the meta-regression model to account for ancestry-related variation in allelic effects. Ancestry-correlated heterogeneity was estimated for each variant, with significant heterogeneity defined as P-value <5 × 10^−8^. Genome-wide associations were evaluated at P-value <5 × 10^−8^. Because the external datasets assessed chronic gastritis rather than AG/SAG specifically, these analyses were interpreted as external supportive analyses, distinct from the primary Japanese serological AG/SAG GWAS.

## Results

### Study population and analysis workflow

We performed Japanese GWAS meta-analyses involving 214,747 participants (Supplementary Table 1). The discovery phase included 75,074 participants, comprising 15,644 AG cases and 4,517 SAG cases. The replication phase included 139,673 participants, with 9,548 AG cases; the SAG analysis excluded BBJ and included 49,964 participants, with 2,303 SAG cases. The workflow included overall and serostatus-stratified GWAS, genotype-by-serostatus interactions, HLA and ABO association analyses, analyses of shared genetic architecture with GC, and cross-ancestry meta-analysis. An overview of the workflow is provided in **Figure 1**.

### Principal loci identified in the Japanese GWAS

In the overall and serostatus-stratified AG/SAG GWAS, genome-wide significant associations were identified at 17 regions (**Table 1**): *TTLL10*, *MUC1*/*MTX1*, *FAP*, *CCDC80*, *LRRC34*, *PRKAA1*/*PTGER4*, *CPEB4*, MHC class I, MHC class II, *PGC*/*FRS3*, *PRDM1*, *CCR6*, *LMTK2*, *PSCA*, *ABO*, *ATP4A*, and *BMP2*.

**Table 1.** Regional lead variants at 17 genome-wide significant loci identified in the Japanese AG/SAG GWAS meta-analyses.

| Locus | GWS analyses | Strongest analysis | Lead variant | Position (GRCh37) | Effect allele | EAF | OR (95% CI) | P-value | Nearest gene | Annotation |
|---|---|---|---|---|---|---|---|---|---|---|
| 1p36.33 | AG | AG | rs58980341 | chr1:1135107 | T | 0.18 | 1.101 (1.071-1.131) | 3.35 × 10<sup>−11</sup> | TTLL10 | intergenic |
| 1q22 | AG; SAG; HP+ AG; HP+ SAG | HP+ AG | rs760077 | chr1:155178782 | T | 0.854 | 1.252 (1.209-1.297) | 1.40 × 10<sup>−34</sup> | MTX1 | missense |
| 2q24.2 | HP- SAG | HP- SAG | rs11679244 | chr2:163082395 | A | 0.172 | 1.514 (1.305-1.758) | 4.21 × 10<sup>−8</sup> | FAP | intronic |
| 3q13.2 | AG | AG | rs72943992 | chr3:112403577 | C | 0.63 | 1.102 (1.078-1.126) | 8.19 × 10<sup>−18</sup> | CCDC80 | intergenic |
| 3q26.2 | AG | AG | rs28626343 | chr3:169526272 | C | 0.653 | 1.063 (1.040-1.086) | 4.59 × 10<sup>−8</sup> | LRRC34 | intronic |
| 5p13.1 | SAG; HP+ SAG | SAG | rs28540420 | chr5:40687463 | C | 0.421 | 1.130 (1.088-1.173) | 1.03 × 10<sup>−10</sup> | PTGER4 | intronic |
| 5q35.2 | HP+ AG | HP+ AG | rs359456 | chr5:173279808 | C | 0.428 | 1.090 (1.062-1.118) | 2.71 × 10<sup>−11</sup> | CPEB4 | intergenic |
| 6p22.1 | AG; SAG; HP- SAG | AG | rs9260603 | chr6:29922754 | C | 0.592 | 1.084 (1.061-1.108) | 9.20 × 10<sup>−14</sup> | HLA-A | intergenic |
| 6p21.3 | AG; SAG; HP+ AG; HP+ SAG; HP- AG; HP- SAG | HP- SAG | rs9269047 | chr6:32438783 | C | 0.096 | 2.370 (1.991-2.822) | 4.74 × 10<sup>−22</sup> | HLA-DRA | intergenic |
| 6p21.1 | AG; HP+ AG | AG | rs62396739 | chr6:41730412 | A | 0.266 | 1.103 (1.077-1.129) | 4.23 × 10<sup>−16</sup> | FRS3 | intergenic |
| 6q21 | SAG | SAG | rs4946699 | chr6:106263758 | A | 0.17 | 1.428 (1.262-1.615) | 1.81 × 10<sup>−8</sup> | PRDM1 | intergenic |
| 6q27 | HP- SAG | HP- SAG | rs204295 | chr6:167500562 | C | 0.536 | 1.350 (1.217-1.498) | 1.46 × 10<sup>−8</sup> | CCR6 | intergenic |
| 7q21.3 | AG; HP+ AG | AG | rs58880085 | chr7:97836465 | C | 0.278 | 1.070 (1.045-1.096) | 3.05 × 10<sup>−8</sup> | LMTK2 | 3' UTR |
| 8q24.3 | AG; SAG; HP+ AG; HP+ SAG | HP+ AG | rs2920279 | chr8:143762135 | C | 0.641 | 1.335 (1.299-1.372) | 2.48 × 10<sup>−101</sup> | PSCA | intronic |
| 9q34.2 | AG; HP+ AG | HP+ AG | rs494242 | chr9:136145118 | T | 0.419 | 1.093 (1.066-1.121) | 1.03 × 10<sup>−11</sup> | ABO | intronic |
| 19q13.12 | HP- SAG | HP- SAG | rs725986 | chr19:36057622 | T | 0.82 | 1.587 (1.370-1.839) | 8.58 × 10<sup>−10</sup> | ATP4A | intergenic |
| 20p12.3 | AG; HP+ AG | HP+ AG | rs6038583 | chr20:6670473 | G | 0.246 | 1.130 (1.097-1.163) | 6.52 × 10<sup>−16</sup> | BMP2 | intergenic |

Each locus reached genome-wide significance (P < 5 × 10<sup>−8</sup>) in at least one of the six combined Japanese meta-GWAS analyses. GWS analyses lists the combined analyses in which the locus was significant, and Strongest analysis denotes the analysis with the smallest regional lead P value. Statistics are reported for the regional lead variant from that strongest analysis. Gene labels are provided for locus orientation and do not establish causal genes. Anti-*H. pylori* IgG-seropositive (HP+) and seronegative (HP-) strata were defined as >3 U/mL and <=3 U/mL, respectively, and do not directly establish active infection. BBJ was included only in overall AG. Positions are based on GRCh37. AG, atrophic gastritis; BBJ, BioBank Japan; CI, confidence interval; EA, effect allele; EAF, effect allele frequency; GWS, genome-wide significant; OR, odds ratio; SAG, severe atrophic gastritis; SE, standard error.

### Overall AG

In the overall AG analysis, associations were identified at 11 regions (**Figure 2a; Supplementary Table 3**). These included known GC susceptibility regions encompassing *PSCA*, *MUC1/MTX1*, *ABO*, and the MHC. The strongest association was found at 8q24.3, with the lead variant rs2920279 located in an intronic region of *PSCA* (OR = 1.202, 95% confidence interval [CI]: 1.176-1.228, P-value = 3.32 × 10^−61^). An association was also observed at 1q22 for the *MTX1* missense variant rs760077 (OR = 1.134, 95% CI: 1.101-1.168, P-value = 1.45 × 10^−16^). This variant has been reported to be in strong LD with the *MUC1* variant rs4072037 in a Taiwanese population (*r*^2^ = 0.965) [54]. At 9q34.2, the *ABO* intronic variant rs9411372 showed an association (OR = 1.080, 95% CI: 1.053-1.108, P-value = 8.19 × 10^−9^). Within the MHC region, associations were observed for the intergenic variant rs9260603 between *HLA-A* and *HCG9* at 6p22.1 (OR = 1.084, 95% CI: 1.061-1.108, P-value = 9.20 × 10^−14^) and the *HLA-DQA1* intronic variant rs1130153 at 6p21.3 (OR = 1.224, 95% CI: 1.165-1.285, P-value = 2.69 × 10^−16^).

**Figure 2.**
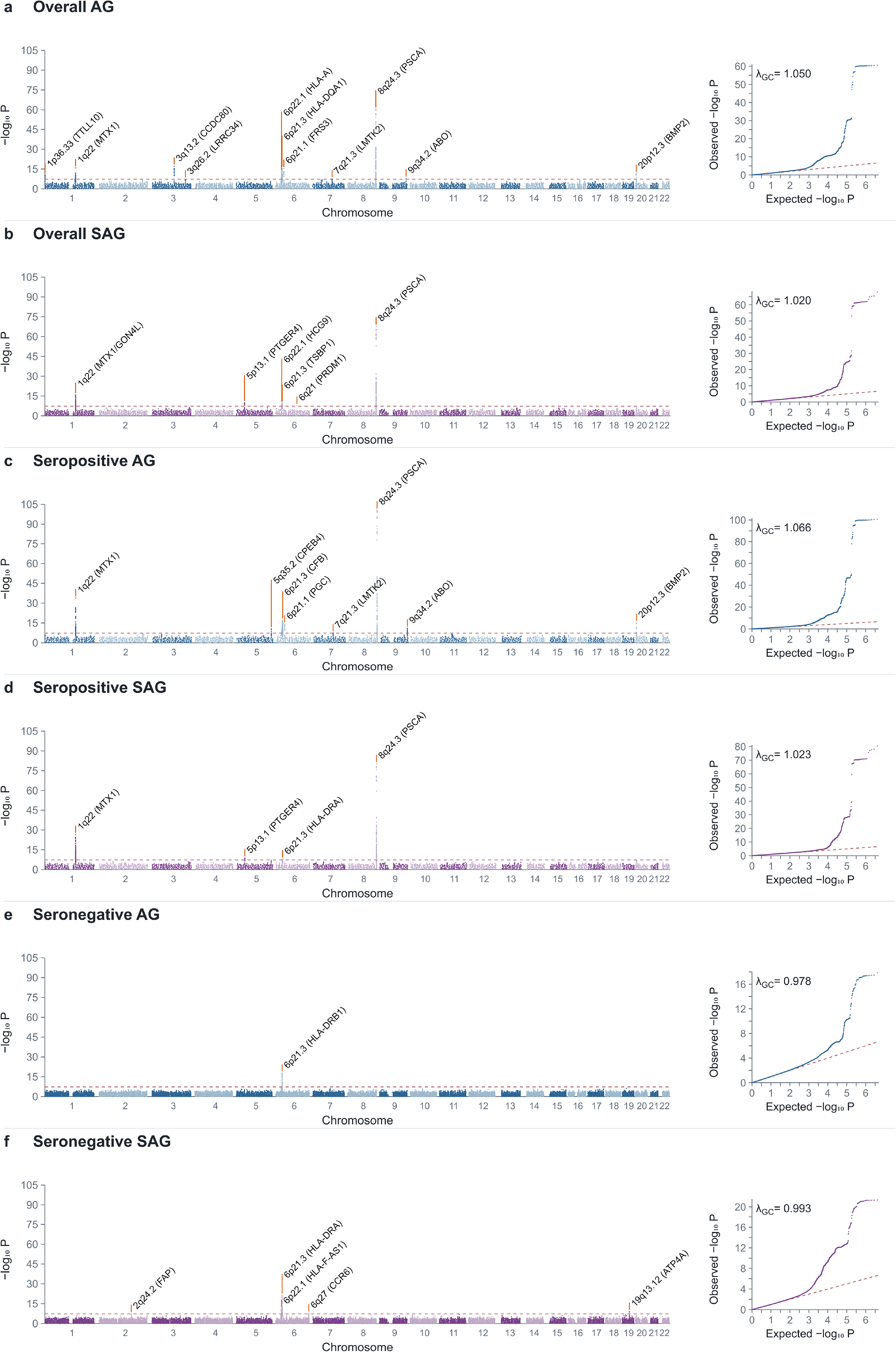
Manhattan and quantile-quantile plots for the combined Japanese AG/SAG GWAS meta-analyses. Manhattan and quantile-quantile plots are shown for (a) overall AG, (b) overall SAG, (c) anti-*H. pylori* IgG-seropositive AG, (d) anti-*H. pylori* IgG-seropositive SAG, (e) anti-*H. pylori* IgG-seronegative AG, and (f) anti-*H. pylori* IgG-seronegative SAG. Manhattan plots show −log_10_(*P*) values by genomic position. The red dashed line indicates the genome-wide significance threshold of *P* = 5 × 10^−8^. Selected genome-wide significant loci are annotated using regional lead variants; Gene labels indicate nearby genes and do not imply causality. Quantile-quantile plots compare observed and expected −log_10_(*P*) values. Genomic positions are based on GRCh37. Seropositive and seronegative strata were defined as anti-*H. pylori* IgG >3 U/mL and <=3 U/mL, respectively, and do not directly establish active infection. Medical diagnosis-based BBJ AG data were included only in panel a. The λ_GC_ values were 1.050, 1.020, 1.066, 1.023, 0.978, and 0.993 for panels a-f, respectively. λ_GC_ was calculated from the median chi-square statistic using reported *P* values in the corresponding source summary-statistics file.

In addition to these findings, associations were identified at regions not known as GC susceptibility loci. These included the *TTLL10*-*TNFRSF18* (*GITR*) intergenic region at 1p36.33 (rs58980341; OR = 1.101, 95% CI: 1.071-1.131, P-value = 3.35 × 10^−11^), the *CASC20*-*BMP2* intergenic region at 20p12.3 (rs6038583; OR = 1.090, 95% CI: 1.064-1.116, P-value = 2.74 × 10^−12^), and an intergenic region downstream of *CCDC80* at 3q13.2 (rs72943992; OR = 1.102, 95% CI: 1.078-1.126, P-value = 8.19 × 10^−18^). Further associations were observed for the *LRRC34* intronic variant rs28626343 at 3q26.2 (OR = 1.063, 95% CI: 1.040-1.086, P-value = 4.59 × 10^−8^), the *PGC*-*FRS3* intergenic variant rs62396739 at 6p21.1 (OR = 1.103, 95% CI: 1.077-1.129, P-value = 4.23 × 10^−16^), and the *LMTK2* 3-prime untranslated region variant rs58880085 at 7q21.3 (OR = 1.070, 95% CI: 1.045-1.096, P-value = 3.05 × 10^−8^).

Associations at the *PSCA* and *MUC1*/*MTX1* regions reached genome-wide significance in both the discovery and replication phases (**Supplementary Figure 1; Supplementary Table 3**). At *PSCA*, the lead variants were rs2978981 in discovery (P-value = 9.23 × 10^−44^) and rs2976384 in replication (P-value = 7.84 × 10^−20^). At *MUC1*/*MTX1*, rs760077 showed associations in both discovery (P-value = 3.32 × 10^−9^) and replication (P-value = 4.92 × 10^−9^). Strong discovery associations were also observed at *CCDC80* and *HLA-DQA1*, although the replication findings provided suggestive evidence below genome-wide significance. The lead variants at *CCDC80* were rs72943992 in discovery (P-value = 2.28 × 10^−12^) and rs12639600 in replication (P-value = 4.89 × 10^−7^), whereas those at *HLA-DQA1* were rs9271399 in discovery (P-value = 1.91 × 10^−15^) and rs1130153 in replication (P-value = 1.65 × 10^−7^). At *BMP2*, the discovery lead rs11087739 showed P-value = 6.24 × 10^−6^, while the replication lead rs6038583 reached P-value = 7.25 × 10^−9^. The *GNG12-AS1* intronic variant rs75563559 at 1p31.3 reached genome-wide significance in discovery (OR = 1.350, 95% CI: 1.217-1.498, P-value = 1.89 × 10^−8^), but the association did not reach this threshold in the combined analysis (P-value = 6.62 × 10^−8^).

### Overall SAG

In the combined SAG analysis, cases were restricted to participants with severe atrophy. Many of the identified regions overlapped with those for overall AG, with stronger associations for SAG at some regions (**Figure 2b; Supplementary Table 4**). The strongest association was observed for the *PSCA* intronic variant rs2920280 (OR = 1.385, 95% CI: 1.335-1.438, P-value = 1.43 × 10^−68^). The *MTX1* missense variant rs760077 showed OR = 1.249 (95% CI: 1.184-1.316, P-value = 1.20 × 10^−16^), and an association was also observed for the *GON4L* intronic variant rs860295 at 1q22 (OR = 1.170, 95% CI: 1.118-1.224, P-value = 2.67 × 10^−12^). At 5p13.1, the *PTGER4* intronic variant rs28540420 showed OR = 1.130 (95% CI: 1.088-1.173, P-value = 1.03 × 10^−10^). This variant is in strong LD with the GC-associated *PRKAA1* intronic variant rs13361707 in Japanese populations (*r*^2^ = 0.961, D′ = 0.981) [10]. The association at this region was more evident for SAG than for AG.

Within the MHC region, associations were observed for rs9268399, located within 2 kb upstream of *TSBP1* in the class II region (OR = 1.147, 95% CI: 1.098-1.197, P-value = 1.98 × 10^−10^), and rs2523945, located within 2 kb upstream of *HCG9* in the class I region (OR = 1.143, 95% CI: 1.095-1.194, P-value = 5.03 × 10^−10^). At 6q21, the *PREP*-*PRDM1* intergenic variant rs4946699 showed OR = 1.428 (95% CI: 1.262-1.615, P-value = 1.81 × 10^−8^).

The *PSCA* and *MUC1*/*MTX1*-*GON4L* associations were replicated at a genome-wide significance level (**Supplementary Figure 2; Supplementary Table 4**). At *PSCA*, the lead variants were rs2920280 in discovery (P-value = 8.98 × 10^−47^) and rs2717609 in replication (P-value = 1.45 × 10^−24^). At *MUC1*/*MTX1*-*GON4L*, the lead variants were rs822478 in discovery (P-value = 1.42 × 10^−8^) and rs760077 in replication (P-value = 1.70 × 10^−11^). In the MHC class II region, the discovery lead rs35656734 showed P-value = 1.63 × 10^−9^, while the replication lead rs77055720 provided suggestive evidence (P-value = 1.53 × 10^−5^). Some regions associated with AG did not reach genome-wide significance for SAG. This may reflect effects at earlier stages of atrophy or smaller effects on progression to severe atrophy. Differences in statistical power due to fewer SAG cases may also contribute.

### Seropositive AG and SAG

Analyses stratified by anti-*H. pylori* IgG serostatus revealed differences in the genetic profiles of AG/SAG. In seropositive AG, stronger associations were observed at some principal AG regions, particularly *PSCA* and *MUC1*/*MTX1* (**Figure 2c; Supplementary Table 5**). The *PSCA* intronic variant rs2920279 showed OR = 1.335 (95% CI: 1.299-1.372, P-value = 2.48 × 10^−101^), and the *MTX1* missense variant rs760077 showed OR = 1.252 (95% CI: 1.209-1.297, P-value = 1.40 × 10^−34^). Other associated variants included *CASC20*-*BMP2* intergenic rs6038583 (OR = 1.130, 95% CI: 1.097-1.163, P-value = 6.52 × 10^−16^), *ABO* intronic rs494242 (OR = 1.093, 95% CI: 1.066-1.121, P-value = 1.03 × 10^−11^), the *PGC*-*FRS3* intergenic variant rs62396738 at 6p21.1 (OR = 1.119, 95% CI: 1.088-1.150, P-value = 9.15 × 10^−15^), and *LMTK2* intronic rs28457167 (OR = 1.087, 95% CI: 1.055-1.119, P-value = 3.10 × 10^−8^). Associations were also observed for the *CPEB4*-*LINC01485* intergenic variant rs359456 at 5q35.2 (OR = 1.090, 95% CI: 1.062-1.118, P-value = 2.71 × 10^−11^) and the *CFB* missense variant rs12614 in the MHC class III region (OR = 1.267, 95% CI: 1.200-1.339, P-value = 9.25 × 10^−18^). Associations at *PSCA*, *MUC1*/*MTX1*, *BMP2*, and *PGC*/*FRS3* reached genome-wide significance in replication, whereas the replication evidence for *CFB* and *ABO* remained below this threshold (**Supplementary Figure 3; Supplementary Table 5**).

In seropositive SAG, associations were observed for *PSCA* rs2920280 (OR = 1.477, 95% CI: 1.420-1.536, P-value = 4.60 × 10^−81^), *MTX1* rs760077 (OR = 1.374, 95% CI: 1.298-1.455, P-value = 4.10 × 10^−27^), and *PTGER4* rs28540420 (OR = 1.132, 95% CI: 1.086-1.180, P-value = 1.78 × 10^−9^) (Figure 2d; Supplementary Table 6). The *HLA-DRA*-*DRB5* intergenic variant rs6940063 showed OR = 1.254 (95% CI: 1.159-1.356, P-value = 1.47 × 10^−8^). Associations at *PSCA* and *MUC1*/*MTX1* were replicated at a genome-wide significance level (**Supplementary Figure 4; Supplementary Table 6**).

### Seronegative AG and SAG

In seronegative AG, a strong association was observed for the *HLA-DRB1*-*DQA1* intergenic variant rs9271065 (OR = 1.799, 95% CI: 1.577-2.051, P-value = 1.40 × 10^−18^) (**Figure 2e; Supplementary Table 7**). The discovery lead rs9469110 showed P-value = 1.43 × 10^−17^, whereas the replication lead rs9269204 provided suggestive evidence at P-value = 1.71 × 10^−6^ (**Supplementary Figure 5; Supplementary Table 7**).

In seronegative SAG, the *HLA-DRA*-*DRB5* intergenic variant rs9269047 showed a strong association (OR = 2.370, 95% CI: 1.991-2.822, P-value = 4.74 × 10^−22^) (**Figure 2f; Supplementary Table 8**), although the replication evidence remained below genome-wide significance (**Supplementary Figure 6; Supplementary Table 8**). A genome-wide significant association was also observed for the *HLA-F-AS1* intronic variant rs9258242 in the MHC class I region (OR = 1.624, 95% CI: 1.416–1.863, P-value = 5.93 × 10^−12^). Additional associations were identified for the *FAP* intronic variant rs11679244 near *IFIH1* at 2q24.2 (OR = 1.514, 95% CI: 1.305-1.758, P-value = 4.21 × 10^−8^), the *FGFR1OP*-*CCR6* intergenic variant rs204295 at 6q27 (OR = 1.350, 95% CI: 1.217-1.498, P-value = 1.46 × 10^−8^), and the *ATP4A*-*LINC01766* intergenic variant rs725986 at 19q13.12 (OR = 1.587, 95% CI: 1.370-1.839, P-value = 8.58 × 10^−10^). In the replication analysis, the *ENPP2*-*TAF2* intergenic variant rs78903306 at 8q24.12 reached genome-wide significance (OR = 5.089, 95% CI: 2.860-9.054, P-value = 3.01 × 10^−8^). However, the lead variant at this region in the combined analysis, rs370391108, did not reach this threshold (OR = 1.879, 95% CI: 1.207-2.927, P-value = 5.27 × 10^−3^).

### Fine-mapping

Fine-mapping was performed for 102 combinations of the 17 principal Japanese GWAS regions and six combined analyses, with six additional analyses targeting *CFB*, giving a total of 108 analyses. The 35 analyses in which the GWAS lead variant reached genome-wide significance are presented in **Supplementary Table 9**. In many analyses, the prioritized credible-set variants were identical to the GWAS lead variant or were in moderate to high LD with it, supporting the corresponding regional associations. The GWAS lead variant itself was prioritized in at least one analysis at *CCDC80*, *LRRC34*, *CPEB4*, *CFB*, and *PRDM1*. Representative variants at *TTLL10*, *MUC1*/*MTX1*, *PGC*/*FRS3*, *CCR6*, and *ATP4A* were in high LD with the corresponding lead variants (*r*^2^ = 0.992, 0.935, 0.918, 0.984, and 0.958, respectively).

On the other hand, lower LD was observed in some MHC analyses, including the class I region in SAG (*r*^2^ = 0.214) and the *HLA-DRA* region in AG (*r*^2^ = 0.129). This may reflect the complex LD structure or the presence of multiple signals. Overall, the fine-mapping findings supported associations within the GWAS regions, although the composition of credible sets varied with the analysis conditions. Many analyses reached the prespecified maximum of five causal components. The results were therefore interpreted as prioritization of candidate variants within associated regions, rather than definitive identification of the number of independent causal signals.

### Genotype-by-anti-*H. pylori* IgG serostatus interactions

The genotype-by-serostatus analyses revealed locus-specific differences in genetic effects (**Supplementary Figure 7; Table 2**). In a logistic regression model without genotype-by-serostatus interaction, the per-allele OR would be the same in seronegative and seropositive participants. Conditional main-effect ORs refer to per-allele genetic associations at mean serostatus and are not stratum-specific estimates. Interaction OR ratios describe the multiplicative change in the per-allele OR per one standard deviation increase in the binary serostatus indicator. Approximate model-derived ORs for the serostatus-by-dosage combinations are shown in **Supplementary Table 10 and Supplementary Figure 8**.

**Table 2.**
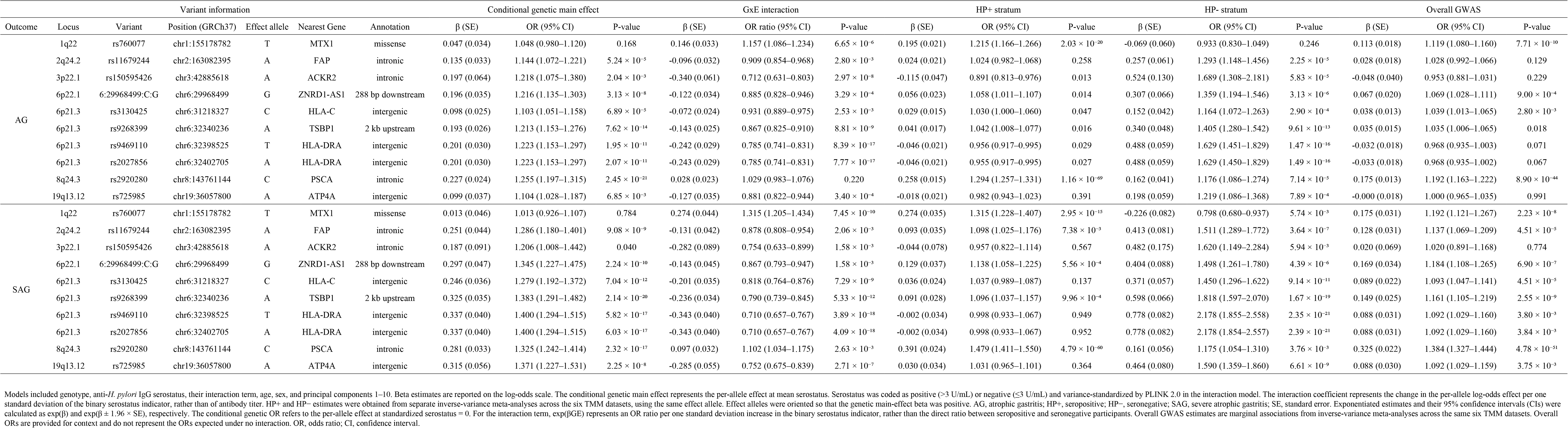
Selected genotype-by-anti-*H. pylori* IgG serostatus interaction results for AG and SAG.

| Variant information |  |  |  |  |  |  | Conditional genetic main effect |  |  | GxE interaction |  |  | HP+ stratum |  |  | HP- stratum |  |  | Overall GWAS |  |  |
|---|---|---|---|---|---|---|---|---|---|---|---|---|---|---|---|---|---|---|---|---|---|
| Outcome | Locus | Variant | Position (GRCh37) | Effect allele | Nearest Gene | Annotation | β (SE) | OR (95% CI) | P-value | β (SE) | OR ratio (95% CI) | P-value | β (SE) | OR (95% CI) | P-value | β (SE) | OR (95% CI) | P-value | β (SE) | OR (95% CI) | P-value |
| AG | 1q22 | rs760077 | chr1:155178782 | T | MTX1 | missense | 0.047 (0.034) | 1.048 (0.980–1.120) | 0.168 | 0.146 (0.033) | 1.157 (1.086–1.234) | 6.65 × 10<sup>-6</sup> | 0.195 (0.021) | 1.215 (1.166–1.266) | 2.03 × 10<sup>-20</sup> | -0.069 (0.060) | 0.933 (0.830–1.049) | 0.246 | 0.113 (0.018) | 1.119 (1.080–1.160) | 7.71 × 10<sup>-10</sup> |
|  | 2q24.2 | rs11679244 | chr2:163082395 | A | FAP | intronic | 0.135 (0.033) | 1.144 (1.072–1.221) | 5.24 × 10<sup>-5</sup> | -0.096 (0.032) | 0.909 (0.854–0.968) | 2.80 × 10<sup>-3</sup> | 0.024 (0.021) | 1.024 (0.982–1.068) | 0.258 | 0.257 (0.061) | 1.293 (1.148–1.456) | 2.25 × 10<sup>-5</sup> | 0.028 (0.018) | 1.028 (0.992–1.066) | 0.129 |
|  | 3p22.1 | rs150595426 | chr3:42885618 | A | ACKR2 | intronic | 0.197 (0.064) | 1.218 (1.075–1.380) | 2.04 × 10<sup>-3</sup> | -0.340 (0.061) | 0.712 (0.631–0.803) | 2.97 × 10<sup>-8</sup> | -0.115 (0.047) | 0.891 (0.813–0.976) | 0.013 | 0.524 (0.130) | 1.689 (1.308–2.181) | 5.83 × 10<sup>-5</sup> | -0.048 (0.040) | 0.953 (0.881–1.031) | 0.229 |
|  | 6p22.1 | 6:29968499:C:G | chr6:29968499 | G | ZNRD1-AS1 | 288 bp downstream | 0.196 (0.035) | 1.216 (1.135–1.303) | 3.13 × 10<sup>-8</sup> | -0.122 (0.034) | 0.885 (0.828–0.946) | 3.29 × 10<sup>-4</sup> | 0.056 (0.023) | 1.058 (1.011–1.107) | 0.014 | 0.307 (0.066) | 1.359 (1.194–1.546) | 3.13 × 10<sup>-6</sup> | 0.067 (0.020) | 1.069 (1.028–1.111) | 9.00 × 10<sup>-4</sup> |
|  | 6p21.3 | rs3130425 | chr6:31218327 | C | HLA-C | intergenic | 0.098 (0.025) | 1.103 (1.051–1.158) | 6.89 × 10<sup>-5</sup> | -0.072 (0.024) | 0.931 (0.889–0.975) | 2.53 × 10<sup>-3</sup> | 0.029 (0.015) | 1.030 (1.000–1.060) | 0.047 | 0.152 (0.042) | 1.164 (1.072–1.263) | 2.90 × 10<sup>-4</sup> | 0.038 (0.013) | 1.039 (1.013–1.065) | 2.80 × 10<sup>-3</sup> |
|  | 6p21.3 | rs9268399 | chr6:32340236 | A | TSBP1 | 2 kb upstream | 0.193 (0.026) | 1.213 (1.153–1.276) | 7.62 × 10<sup>-14</sup> | -0.143 (0.025) | 0.867 (0.825–0.910) | 8.81 × 10<sup>-9</sup> | 0.041 (0.017) | 1.042 (1.008–1.077) | 0.016 | 0.340 (0.048) | 1.405 (1.280–1.542) | 9.61 × 10<sup>-13</sup> | 0.035 (0.015) | 1.035 (1.006–1.065) | 0.018 |
|  | 6p21.3 | rs9469110 | chr6:32398525 | T | HLA-DRA | intergenic | 0.201 (0.030) | 1.223 (1.153–1.297) | 1.95 × 10<sup>-11</sup> | -0.242 (0.029) | 0.785 (0.741–0.831) | 8.39 × 10<sup>-17</sup> | -0.046 (0.021) | 0.956 (0.917–0.995) | 0.029 | 0.488 (0.059) | 1.629 (1.451–1.829) | 1.47 × 10<sup>-16</sup> | -0.032 (0.018) | 0.968 (0.935–1.003) | 0.071 |
|  | 6p21.3 | rs2027856 | chr6:32402705 | A | HLA-DRA | intergenic | 0.201 (0.030) | 1.223 (1.153–1.297) | 2.07 × 10<sup>-11</sup> | -0.243 (0.029) | 0.785 (0.741–0.831) | 7.77 × 10<sup>-17</sup> | -0.046 (0.021) | 0.955 (0.917–0.995) | 0.027 | 0.488 (0.059) | 1.629 (1.450–1.829) | 1.49 × 10<sup>-16</sup> | -0.033 (0.018) | 0.968 (0.935–1.002) | 0.067 |
|  | 8q24.3 | rs2920280 | chr8:143761144 | C | PSCA | intronic | 0.227 (0.024) | 1.255 (1.197–1.315) | 2.45 × 10<sup>-21</sup> | 0.028 (0.023) | 1.029 (0.983–1.076) | 0.220 | 0.258 (0.015) | 1.294 (1.257–1.331) | 1.16 × 10<sup>-69</sup> | 0.162 (0.041) | 1.176 (1.086–1.274) | 7.14 × 10<sup>-5</sup> | 0.175 (0.013) | 1.192 (1.163–1.222) | 8.90 × 10<sup>-44</sup> |
|  | 19q13.12 | rs725985 | chr19:36057800 | A | ATP4A | intergenic | 0.099 (0.037) | 1.104 (1.028–1.187) | 6.85 × 10<sup>-3</sup> | -0.127 (0.035) | 0.881 (0.822–0.944) | 3.40 × 10<sup>-4</sup> | -0.018 (0.021) | 0.982 (0.943–1.023) | 0.391 | 0.198 (0.059) | 1.219 (1.086–1.368) | 7.89 × 10<sup>-4</sup> | -0.000 (0.018) | 1.000 (0.965–1.035) | 0.991 |
| SAG | 1q22 | rs760077 | chr1:155178782 | T | MTX1 | missense | 0.013 (0.046) | 1.013 (0.926–1.107) | 0.784 | 0.274 (0.044) | 1.315 (1.205–1.434) | 7.45 × 10<sup>-10</sup> | 0.274 (0.035) | 1.315 (1.228–1.407) | 2.95 × 10<sup>-15</sup> | -0.226 (0.082) | 0.798 (0.680–0.937) | 5.74 × 10<sup>-3</sup> | 0.175 (0.031) | 1.192 (1.121–1.267) | 2.23 × 10<sup>-8</sup> |
|  | 2q24.2 | rs11679244 | chr2:163082395 | A | FAP | intronic | 0.251 (0.044) | 1.286 (1.180–1.401) | 9.08 × 10<sup>-9</sup> | -0.131 (0.042) | 0.878 (0.808–0.954) | 2.06 × 10<sup>-3</sup> | 0.093 (0.035) | 1.098 (1.025–1.176) | 7.38 × 10<sup>-3</sup> | 0.413 (0.081) | 1.511 (1.289–1.772) | 3.64 × 10<sup>-7</sup> | 0.128 (0.031) | 1.137 (1.069–1.209) | 4.51 × 10<sup>-5</sup> |
|  | 3p22.1 | rs150595426 | chr3:42885618 | A | ACKR2 | intronic | 0.187 (0.091) | 1.206 (1.008–1.442) | 0.040 | -0.282 (0.089) | 0.754 (0.633–0.899) | 1.58 × 10<sup>-3</sup> | -0.044 (0.078) | 0.957 (0.822–1.114) | 0.567 | 0.482 (0.175) | 1.620 (1.149–2.284) | 5.94 × 10<sup>-3</sup> | 0.020 (0.069) | 1.020 (0.891–1.168) | 0.774 |
|  | 6p22.1 | 6:29968499:C:G | chr6:29968499 | G | ZNRD1-AS1 | 288 bp downstream | 0.297 (0.047) | 1.345 (1.227–1.475) | 2.24 × 10<sup>-10</sup> | -0.143 (0.045) | 0.867 (0.793–0.947) | 1.58 × 10<sup>-3</sup> | 0.129 (0.037) | 1.138 (1.058–1.225) | 5.56 × 10<sup>-4</sup> | 0.404 (0.088) | 1.498 (1.261–1.780) | 4.39 × 10<sup>-6</sup> | 0.169 (0.034) | 1.184 (1.108–1.265) | 6.90 × 10<sup>-7</sup> |
|  | 6p21.3 | rs3130425 | chr6:31218327 | C | HLA-C | intergenic | 0.246 (0.036) | 1.279 (1.192–1.372) | 7.04 × 10<sup>-12</sup> | -0.201 (0.035) | 0.818 (0.764–0.876) | 7.29 × 10<sup>-9</sup> | 0.036 (0.024) | 1.037 (0.989–1.087) | 0.137 | 0.371 (0.057) | 1.450 (1.296–1.622) | 9.14 × 10<sup>-11</sup> | 0.089 (0.022) | 1.093 (1.047–1.141) | 4.51 × 10<sup>-5</sup> |
|  | 6p21.3 | rs9268399 | chr6:32340236 | A | TSBP1 | 2 kb upstream | 0.325 (0.035) | 1.383 (1.291–1.482) | 2.14 × 10<sup>-20</sup> | -0.236 (0.034) | 0.790 (0.739–0.845) | 5.33 × 10<sup>-12</sup> | 0.091 (0.028) | 1.096 (1.037–1.157) | 9.96 × 10<sup>-4</sup> | 0.598 (0.066) | 1.818 (1.597–2.070) | 1.67 × 10<sup>-19</sup> | 0.149 (0.025) | 1.161 (1.105–1.219) | 2.55 × 10<sup>-9</sup> |
|  | 6p21.3 | rs9469110 | chr6:32398525 | T | HLA-DRA | intergenic | 0.337 (0.040) | 1.400 (1.294–1.515) | 5.82 × 10<sup>-17</sup> | -0.343 (0.040) | 0.710 (0.657–0.767) | 3.89 × 10<sup>-18</sup> | -0.002 (0.034) | 0.998 (0.933–1.067) | 0.949 | 0.778 (0.082) | 2.178 (1.855–2.558) | 2.35 × 10<sup>-21</sup> | 0.088 (0.031) | 1.092 (1.029–1.160) | 3.80 × 10<sup>-3</sup> |
|  | 6p21.3 | rs2027856 | chr6:32402705 | A | HLA-DRA | intergenic | 0.337 (0.040) | 1.400 (1.294–1.515) | 6.03 × 10<sup>-17</sup> | -0.343 (0.040) | 0.710 (0.657–0.767) | 4.09 × 10<sup>-18</sup> | -0.002 (0.034) | 0.998 (0.933–1.067) | 0.952 | 0.778 (0.082) | 2.178 (1.854–2.557) | 2.39 × 10<sup>-21</sup> | 0.088 (0.031) | 1.092 (1.029–1.160) | 3.84 × 10<sup>-3</sup> |
|  | 8q24.3 | rs2920280 | chr8:143761144 | C | PSCA | intronic | 0.281 (0.033) | 1.325 (1.242–1.414) | 2.32 × 10<sup>-17</sup> | 0.097 (0.032) | 1.102 (1.034–1.175) | 2.63 × 10<sup>-3</sup> | 0.391 (0.024) | 1.479 (1.411–1.550) | 4.79 × 10<sup>-60</sup> | 0.161 (0.056) | 1.175 (1.054–1.310) | 3.76 × 10<sup>-3</sup> | 0.325 (0.022) | 1.384 (1.327–1.444) | 4.78 × 10<sup>-51</sup> |
|  | 19q13.12 | rs725985 | chr19:36057800 | A | ATP4A | intergenic | 0.315 (0.056) | 1.371 (1.227–1.531) | 2.25 × 10<sup>-8</sup> | -0.285 (0.055) | 0.752 (0.675–0.839) | 2.71 × 10<sup>-7</sup> | 0.030 (0.034) | 1.031 (0.965–1.101) | 0.364 | 0.464 (0.080) | 1.590 (1.359–1.860) | 6.61 × 10<sup>-9</sup> | 0.088 (0.030) | 1.092 (1.029–1.159) | 3.75 × 10<sup>-3</sup> |

Models included genotype, anti-*H. pylori* IgG serostatus, their interaction term, age, sex, and principal components 1–10. Beta estimates are reported on the log-odds scale. The conditional genetic main effect represents the per-allele effect at mean serostatus. Serostatus was coded as positive (>3 U/mL) or negative (≤3 U/mL) and variance-standardized by PLINK 2.0 in the interaction model. The interaction coefficient represents the change in the per-allele log-odds effect per one standard deviation of the binary serostatus indicator, rather than of antibody titer. HP+ and HP− estimates were obtained from separate inverse-variance meta-analyses across the six TMM datasets, using the same effect allele. Effect alleles were oriented so that the genetic main-effect beta was positive. AG, atrophic gastritis; HP+, seropositive; HP−, seronegative; SAG, severe atrophic gastritis; SE, standard error. Exponentiated estimates and their 95% confidence intervals (CIs) were calculated as exp(β) and exp(β ± 1.96 × SE), respectively. The conditional genetic OR refers to the per-allele effect at standardized serostatus = 0. For the interaction term, exp(βGE) represents an OR ratio per one standard deviation increase in the binary serostatus indicator, rather than the direct ratio between seropositive and seronegative participants. Overall GWAS estimates are marginal associations from inverse-variance meta-analyses across the same six TMM datasets. Overall ORs are provided for context and do not represent the ORs expected under no interaction. OR, odds ratio; CI, confidence interval.

At MUC1/MTX1, the rs760077 T allele showed a genome-wide significant positive interaction for SAG (interaction OR ratio = 1.31, 95% CI, 1.20–1.43, P-value = 7.45 × 10⁻¹⁰), with evidence of a positive interaction also observed for AG (interaction OR ratio = 1.16, 95% CI, 1.09–1.23, P-value = 6.65 × 10⁻⁶). The conditional genetic main effects were not significant for either AG (OR = 1.05, 95% CI, 0.98–1.12, P-value = 0.168) or SAG (OR = 1.01, 95% CI, 0.93–1.11, P-value = 0.784). In the TMM stratified meta-analyses, the rs760077 T-allele OR for AG was 0.93 (95% CI, 0.83–1.05) in seronegative participants and 1.22 (95% CI, 1.17–1.27) in seropositive participants. The corresponding ORs for SAG were 0.80 (95% CI, 0.68–0.94) and 1.31 (95% CI, 1.23–1.41), respectively. The T allele was associated with higher odds of AG and SAG in seropositive participants. In seronegative participants, the T allele showed no clear association or was associated with lower odds of AG/SAG. This pattern is consistent with the positive interaction terms, particularly for SAG.

At PSCA, strong conditional main effects were observed for the rs2920280 C allele in AG (OR = 1.25, 95% CI, 1.20–1.31, P-value = 2.45 × 10⁻²¹) and SAG (OR = 1.32, 95% CI, 1.24–1.41, P-value = 2.32 × 10⁻¹⁷). The interaction terms did not reach genome-wide significance, with an interaction OR ratio of 1.03 (95% CI, 0.98–1.08, P-value = 0.220) for AG and 1.10 (95% CI, 1.03–1.17, P-value = 2.63 × 10⁻³) for SAG. Although associations were more robust in the seropositive strata, the direction of association was consistent across strata, including seronegative AG (OR = 1.18, 95% CI, 1.09–1.27, P-value = 7.14 × 10⁻⁵).

At HLA-DRA, negative interactions were observed for the rs9469110 T allele in AG (interaction OR ratio = 0.78, 95% CI, 0.74–0.83, P-value = 8.39 × 10⁻¹⁷) and SAG (interaction OR ratio = 0.71, 95% CI, 0.66–0.77, P-value = 3.89 × 10⁻¹⁸). The conditional main effects were positive for both AG (OR = 1.22, 95% CI, 1.15–1.30, P-value = 1.95 × 10⁻¹¹) and SAG (OR = 1.40, 95% CI, 1.29–1.52, P-value = 5.82 × 10⁻¹⁷). A similar pattern was observed for the rs2027856 A allele. In the TMM stratified meta-analyses, the rs9469110 T-allele OR for AG was 1.63 (95% CI, 1.45–1.83) in seronegative participants and 0.96 (95% CI, 0.92–1.00) in seropositive participants. The corresponding ORs for SAG were 2.18 (95% CI, 1.85–2.56) and 1.00 (95% CI, 0.93–1.07), respectively. The T allele was associated with higher odds of AG and SAG in seronegative participants. In seropositive participants, the T allele showed weak or no clear associations with AG/SAG. This pattern is consistent with the negative interaction terms.

In the ACKR2/KRBOX1-overlapping region, the rs150595426 A allele showed a negative interaction for AG (interaction OR ratio = 0.71, 95% CI, 0.63–0.80, P-value = 2.97 × 10⁻⁸), whereas the interaction for SAG did not reach genome-wide significance (interaction OR ratio = 0.75, 95% CI, 0.63–0.90, P-value = 1.58 × 10⁻³). The conditional main-effect ORs were 1.22 (95% CI, 1.07–1.38, P-value = 2.04 × 10⁻³) for AG and 1.21 (95% CI, 1.01–1.44, P-value = 0.040) for SAG. Genome-wide significance was not reached in either stratum-specific analysis, although suggestive evidence was observed in seronegative AG (OR = 1.69, 95% CI, 1.31–2.18, P-value = 5.83 × 10⁻⁵). At this locus, ACKR2 encodes an atypical chemokine receptor that regulates the localization and concentration of inflammatory CC chemokines [55], whereas KRBOX1 (also known as KRABD1) encodes a KRAB-domain-containing protein with a predicted role in transcriptional regulation [56]. These annotations suggest chemokine regulation or transcriptional control as potential processes underlying the observed interaction, although the causal gene and mechanism remain unclear.

Conditional genetic main effects reached genome-wide significance at *FAP*, *ZNRD1-AS1*, and *ATP4A*, whereas the corresponding interaction terms did not. At *FAP*, which encodes a fibroblast-associated serine protease implicated in GC progression [57], the rs11679244 A allele was associated with SAG (OR = 1.286, 95% CI, 1.180–1.401, P-value = 9.08 × 10⁻⁹). At *ZNRD1-AS1*, a long noncoding RNA implicated in GC cell proliferation and metastasis [58], the G allele of 6:29968499:C:G was associated with both AG (OR = 1.216, 95% CI, 1.135–1.303, P-value = 3.13 × 10⁻⁸) and SAG (OR = 1.345, 95% CI, 1.227–1.475, P-value = 2.24 × 10⁻¹⁰). At *ATP4A*, which encodes the gastric proton-pump α-subunit targeted in autoimmune gastritis [59], the rs725985 A allele was associated with SAG (OR = 1.371, 95% CI, 1.227–1.531, P-value = 2.25 × 10⁻⁸).

### HLA allele associations

HLA allele association analysis revealed differences according to anti-*H. pylori* IgG serostatus (**Table 3**). In seronegative AG, strong associations were identified for *HLA-DRB1*15:01* (OR = 1.485, 95% CI: 1.337-1.649, P-value = 1.44 × 10^−13^) and *HLA-DQB1*06:02* (OR = 1.537, 95% CI: 1.364-1.733, P-value = 2.10 × 10^−12^). These associations were stronger in seronegative SAG, with OR = 1.777 (95% CI: 1.549-2.038, P-value = 1.90 × 10^−16^) for *HLA-DRB1*15:01* and OR = 1.901 (95% CI: 1.632-2.215, P-value = 1.72 × 10^−16^) for *HLA-DQB1*06:02*. The two alleles may represent a shared DR15-DQ6 haplotypic signal. Related HLA associations have been reported in multiple sclerosis and narcolepsy [60,61].

**Table 3.** Associations of classical HLA alleles with AG/SAG overall and by anti-*H. pylori* IgG serostatus.

| Subgroup | Trait | Allele | Case frequency | Control frequency | Beta | SE | OR | 95% CI | P-value |
|---|---|---|---|---|---|---|---|---|---|
| total | AG | HLA-DRB1*13:02 | 0.053 | 0.062 | -0.172 | 0.029 | 0.842 | 0.7959-0.8913 | 3.01 × 10<sup>−9</sup> |
|  |  | HLA-B*44:03 | 0.050 | 0.059 | -0.177 | 0.030 | 0.838 | 0.7894-0.8893 | 5.61 × 10<sup>−9</sup> |
|  |  | HLA-C*14:03 | 0.047 | 0.056 | -0.180 | 0.031 | 0.835 | 0.7857-0.8879 | 8.43 × 10<sup>−9</sup> |
|  |  | HLA-DQB1*06:04 | 0.046 | 0.055 | -0.178 | 0.031 | 0.837 | 0.7879-0.8897 | 1.01 × 10<sup>−8</sup> |
|  |  | HLA-DPA1*01:03 | 0.417 | 0.432 | -0.069 | 0.014 | 0.933 | 0.9078-0.9590 | 7.58 × 10<sup>−7</sup> |
|  |  | HLA-B*44 | 0.054 | 0.062 | -0.144 | 0.029 | 0.866 | 0.8181-0.9170 | 8.02 × 10<sup>−7</sup> |
|  |  | HLA-DPA1*02:02 | 0.499 | 0.484 | 0.061 | 0.013 | 1.063 | 1.0357-1.0919 | 5.04 × 10<sup>−6</sup> |
|  |  | HLA-DQA1*03:01 | 0.364 | 0.351 | 0.062 | 0.014 | 1.064 | 1.0353-1.0932 | 8.01 × 10<sup>−6</sup> |
|  |  | HLA-DPB1*05 | 0.413 | 0.398 | 0.060 | 0.014 | 1.062 | 1.0335-1.0909 | 1.29 × 10<sup>−5</sup> |
|  |  | HLA-DPA1*01 | 0.475 | 0.488 | -0.060 | 0.014 | 0.941 | 0.9160-0.9673 | 1.33 × 10<sup>−5</sup> |
|  |  | HLA-A*33:03 | 0.053 | 0.060 | -0.129 | 0.030 | 0.879 | 0.8286-0.9317 | 1.53 × 10<sup>−5</sup> |
|  |  | HLA-DPA1*02 | 0.522 | 0.509 | 0.060 | 0.014 | 1.062 | 1.0331-1.0909 | 1.60 × 10<sup>−5</sup> |
|  |  | HLA-A*24 | 0.342 | 0.330 | 0.060 | 0.014 | 1.062 | 1.0329-1.0912 | 2.05 × 10<sup>−5</sup> |
|  |  | HLA-A*33 | 0.053 | 0.060 | -0.127 | 0.030 | 0.881 | 0.8306-0.9339 | 2.16 × 10<sup>−5</sup> |
|  |  | HLA-DQA1*03 | 0.360 | 0.347 | 0.058 | 0.014 | 1.060 | 1.0316-1.0894 | 2.63 × 10<sup>−5</sup> |
|  |  | HLA-DPB1*05:01 | 0.410 | 0.396 | 0.057 | 0.014 | 1.059 | 1.0308-1.0877 | 3.13 × 10<sup>−5</sup> |
|  |  | HLA-A*24:02 | 0.299 | 0.289 | 0.061 | 0.015 | 1.063 | 1.0315-1.0948 | 6.03 × 10<sup>−5</sup> |
|  |  | HLA-DPB1*02:01 | 0.285 | 0.296 | -0.058 | 0.015 | 0.944 | 0.9163-0.9718 | 1.11 × 10<sup>−4</sup> |
|  |  | HLA-DRB1*04 | 0.234 | 0.223 | 0.059 | 0.016 | 1.060 | 1.0284-1.0933 | 1.79 × 10<sup>−4</sup> |
|  | SAG | HLA-A*26 | 0.161 | 0.142 | 0.152 | 0.031 | 1.164 | 1.0949-1.2378 | 1.21 × 10<sup>−6</sup> |
|  |  | HLA-A*26:01 | 0.171 | 0.153 | 0.138 | 0.030 | 1.148 | 1.0817-1.2191 | 5.93 × 10<sup>−6</sup> |
|  |  | HLA-DQB1*06:03 | 0.018 | 0.013 | 0.374 | 0.091 | 1.454 | 1.2160-1.7386 | 4.02 × 10<sup>−5</sup> |
|  |  | HLA-DQB1*06:04 | 0.044 | 0.054 | -0.220 | 0.055 | 0.802 | 0.7204-0.8934 | 5.95 × 10<sup>−5</sup> |
|  |  | HLA-DRB1*13:02 | 0.051 | 0.061 | -0.205 | 0.051 | 0.814 | 0.7366-0.9003 | 6.00 × 10<sup>−5</sup> |
|  |  | HLA-B*44:03 | 0.048 | 0.057 | -0.205 | 0.053 | 0.814 | 0.7333-0.9041 | 1.18 × 10<sup>−4</sup> |
| HP-positive | AG | HLA-C*14:03 | 0.047 | 0.059 | -0.242 | 0.034 | 0.785 | 0.7344-0.8391 | 1.08 × 10<sup>−12</sup> |
|  |  | HLA-B*44:03 | 0.050 | 0.061 | -0.230 | 0.033 | 0.795 | 0.7448-0.8480 | 3.68 × 10<sup>−12</sup> |
|  |  | HLA-DRB1*13:02 | 0.052 | 0.064 | -0.218 | 0.032 | 0.804 | 0.7558-0.8562 | 7.86 × 10<sup>−12</sup> |
|  |  | HLA-DQB1*06:04 | 0.046 | 0.056 | -0.219 | 0.034 | 0.803 | 0.7515-0.8590 | 1.40 × 10<sup>−10</sup> |
|  |  | HLA-B*44 | 0.054 | 0.065 | -0.195 | 0.032 | 0.823 | 0.7735-0.8762 | 1.01 × 10<sup>−9</sup> |
|  |  | HLA-A*33:03 | 0.053 | 0.064 | -0.197 | 0.033 | 0.821 | 0.7704-0.8757 | 1.72 × 10<sup>−9</sup> |
|  |  | HLA-A*33 | 0.053 | 0.064 | -0.195 | 0.033 | 0.823 | 0.7714-0.8769 | 2.25 × 10<sup>−9</sup> |
|  |  | HLA-A*24 | 0.341 | 0.325 | 0.077 | 0.016 | 1.081 | 1.0478-1.1144 | 8.44 × 10<sup>−7</sup> |
|  |  | HLA-A*24:02 | 0.299 | 0.284 | 0.083 | 0.017 | 1.087 | 1.0511-1.1236 | 9.68 × 10<sup>−7</sup> |
|  |  | HLA-C*14 | 0.116 | 0.125 | -0.092 | 0.023 | 0.912 | 0.8721-0.9544 | 6.49 × 10<sup>−5</sup> |
|  | SAG | HLA-DQB1*06:03 | 0.019 | 0.013 | 0.406 | 0.098 | 1.501 | 1.2397-1.8176 | 3.16 × 10<sup>−5</sup> |
| HP-negative | AG | HLA-DRB1*15:01 | 0.192 | 0.134 | 0.395 | 0.053 | 1.485 | 1.3369-1.6488 | 1.44 × 10<sup>−13</sup> |
|  |  | HLA-DQB1*06:02 | 0.140 | 0.091 | 0.430 | 0.061 | 1.537 | 1.3635-1.7332 | 2.10 × 10<sup>−12</sup> |
|  |  | HLA-DQA1*01:02 | 0.228 | 0.192 | 0.200 | 0.052 | 1.222 | 1.1041-1.3522 | 1.07 × 10<sup>−4</sup> |
|  | SAG | HLA-DQB1*06:02 | 0.168 | 0.091 | 0.642 | 0.078 | 1.901 | 1.6317-2.2153 | 1.72 × 10<sup>−16</sup> |
|  |  | HLA-DRB1*15:01 | 0.221 | 0.133 | 0.575 | 0.070 | 1.777 | 1.5493-2.0377 | 1.90 × 10<sup>−16</sup> |
|  |  | HLA-DRB1*15:02 | 0.085 | 0.135 | -0.542 | 0.109 | 0.581 | 0.4695-0.7198 | 6.44 × 10<sup>−7</sup> |
|  |  | HLA-B*35:01 | 0.141 | 0.095 | 0.428 | 0.087 | 1.534 | 1.2939-1.8191 | 8.46 × 10<sup>−7</sup> |
|  |  | HLA-DPB1*09 | 0.036 | 0.074 | -0.799 | 0.164 | 0.450 | 0.3265-0.6198 | 1.04 × 10<sup>−6</sup> |
|  |  | HLA-DPB1*09:01 | 0.037 | 0.075 | -0.782 | 0.160 | 0.457 | 0.3339-0.6262 | 1.07 × 10<sup>−6</sup> |
|  |  | HLA-DQA1*01:02 | 0.247 | 0.192 | 0.318 | 0.069 | 1.374 | 1.2004-1.5732 | 4.04 × 10<sup>−6</sup> |
|  |  | HLA-DQB1*06:01 | 0.114 | 0.161 | -0.422 | 0.094 | 0.655 | 0.5450-0.7882 | 7.06 × 10<sup>−6</sup> |
|  |  | HLA-B*52 | 0.057 | 0.091 | -0.579 | 0.133 | 0.560 | 0.4318-0.7272 | 1.32 × 10<sup>−5</sup> |
|  |  | HLA-DPA1*02:01 | 0.051 | 0.086 | -0.598 | 0.139 | 0.550 | 0.4185-0.7223 | 1.74 × 10<sup>−5</sup> |
|  |  | HLA-A*26 | 0.189 | 0.141 | 0.321 | 0.075 | 1.379 | 1.1905-1.5962 | 1.75 × 10<sup>−5</sup> |
|  |  | HLA-C*12:02 | 0.063 | 0.098 | -0.543 | 0.127 | 0.581 | 0.4534-0.7450 | 1.83 × 10<sup>−5</sup> |
|  |  | HLA-C*12 | 0.063 | 0.096 | -0.543 | 0.128 | 0.581 | 0.4523-0.7459 | 2.07 × 10<sup>−5</sup> |
|  |  | HLA-B*35 | 0.134 | 0.094 | 0.374 | 0.088 | 1.453 | 1.2227-1.7277 | 2.25 × 10<sup>−5</sup> |
|  |  | HLA-DQB1*03:02 | 0.188 | 0.144 | 0.311 | 0.075 | 1.365 | 1.1774-1.5823 | 3.71 × 10<sup>−5</sup> |
|  |  | HLA-B*52:01 | 0.063 | 0.095 | -0.521 | 0.127 | 0.594 | 0.4632-0.7621 | 4.17 × 10<sup>−5</sup> |
|  |  | HLA-A*26:01 | 0.198 | 0.152 | 0.292 | 0.073 | 1.340 | 1.1603-1.5471 | 6.79 × 10<sup>−5</sup> |
|  |  | HLA-DRB1*08:02 | 0.067 | 0.042 | 0.498 | 0.127 | 1.645 | 1.2831-2.1101 | 8.69 × 10<sup>−5</sup> |
|  |  | HLA-C*03:03 | 0.194 | 0.150 | 0.290 | 0.075 | 1.337 | 1.1540-1.5490 | 1.11 × 10<sup>−4</sup> |

The table includes HLA alleles meeting the Bonferroni-corrected significance threshold of P < 0.05/273 (approximately 1.83 × 10<sup>−4</sup>). Each imputed HLA allele was analyzed as an expected allele dosage from 0 to 2 in a logistic regression model adjusted for age, sex, and principal components 1-10. Alleles with an estimated frequency <0.01 were excluded. Beta estimates are on the log-odds scale; odds ratios and 95% confidence intervals were calculated as exp(beta) and exp(beta +/- 1.96 x SE), respectively. Anti-*H. pylori* IgG seropositivity and seronegativity were defined as >3 U/mL and <=3 U/mL, respectively. Correlated alleles, different resolutions of the same allele group, and alleles marking the same haplotypic background should not be interpreted as independent signals. AG, atrophic gastritis; CI, confidence interval; HLA, human leukocyte antigen; OR, odds ratio; SAG, severe atrophic gastritis; SE, standard error.
In a Pan-Asian reference panel (530 individuals; 1,060 haplotypes), 33 of 38 haplotypes carrying DQB1\*06:02 also carried DRB1\*15:01 (r<sup>2</sup> = 0.249, D′ = 0.854).

In seropositive AG, *HLA-C*14:03* (OR = 0.785, 95% CI: 0.734-0.839, P-value = 1.08 × 10^−12^) and *HLA-DRB1*13:02* (OR = 0.804, 95% CI: 0.756-0.856, P-value = 7.86 × 10^−12^) were associated with lower odds. An association with seropositive SAG was observed for *HLA-DQB1*06:03* (OR = 1.501, 95% CI: 1.240-1.818, P-value = 3.16 × 10^−5^), whereas the associations of *DRB1*15:01* and *DQB1*06:02* were less evident in seropositive participants. In the overall AG analysis, lower odds were associated with *HLA-DRB1*13:02* (OR = 0.842, 95% CI: 0.796-0.891, P-value = 3.01 × 10^−9^) and *HLA-C*14:03* (OR = 0.835, 95% CI: 0.786-0.888, P-value = 8.43 × 10^−9^). Overall SAG showed associations with *HLA-A*26:01* (OR = 1.148, 95% CI: 1.082-1.219, P-value = 5.93 × 10^−6^), which was also associated with seronegative SAG, and *HLA-DQB1*06:03* (OR = 1.454, 95% CI: 1.216-1.739, P-value = 4.02 × 10^−5^), which was also associated with seropositive SAG. These results indicate that stratification helped distinguish HLA signals combined in the overall analyses.

### ABO blood group associations

Associations of ABO blood group were examined using O as the reference group (**Supplementary Table 11**). In seropositive AG, the highest OR was observed for AB (1.219, 95% CI: 1.123-1.324, P-value = 2.27 × 10^−6^), followed by A (OR = 1.139, 95% CI: 1.081-1.201, P-value = 1.27 × 10^−6^) and B (OR = 1.080, 95% CI: 1.017-1.147, P-value = 0.012). The three-degree-of-freedom likelihood ratio test showed P-value = 1.72 × 10^−7^. Associations with overall AG were also observed for AB (OR = 1.131, 95% CI: 1.055-1.212, P-value = 5.52 × 10^−4^) and A (OR = 1.087, 95% CI: 1.038-1.137, P-value = 3.40 × 10^−4^), with an omnibus P-value = 3.04 × 10^−5^. The A and AB associations and the omnibus tests for overall and seropositive AG remained significant after Bonferroni correction. In contrast, no individual blood group showed a clear association with seronegative AG, although a nominal omnibus association was observed (P-value = 0.049).

A similar tendency toward higher odds for AB was observed in the SAG analyses, with nominally significant associations for overall and seronegative SAG: OR = 1.136 (95% CI: 1.009-1.279, P-value = 0.035) for overall SAG, OR = 1.133 (95% CI: 0.992-1.294, P-value = 0.065) for seropositive SAG, and OR = 1.350 (95% CI: 1.021-1.786, P-value = 0.035) for seronegative SAG. The corresponding omnibus P-values were 0.157, 0.186, and 0.019, respectively. Thus, a nominal overall blood group association was observed only in the seronegative SAG stratum.

When anti-*H. pylori* IgG seropositivity, defined using the 3 U/mL cutoff, was used as the outcome, no clear association was observed for A (OR = 0.994, 95% CI: 0.954-1.035, P-value = 0.765). By contrast, AB (OR = 0.913, 95% CI: 0.856-0.973, P-value = 5.46 × 10^−3^) and B (OR = 0.949, 95% CI: 0.906-0.994, P-value = 0.027) were nominally associated with lower odds. The omnibus P-value was 8.15 × 10^−3^. Previous epidemiological studies have reported associations of blood group A with gastritis or GC [62]. In our study, however, AB showed a higher OR for AG than A, while also showing lower odds of seropositivity. These findings suggest that the associations of ABO blood group with anti-*H. pylori* IgG serostatus and with atrophy are not necessarily in the same direction.

### Shared genetic architecture with GC

To avoid sample overlap with the BBJ GC GWAS, Japanese AG meta-analysis statistics excluding BBJ were used (**Supplementary Figure 9; Supplementary Table 12**). GC summary statistics were obtained from PheWeb.jp [44]. The overall AG associations were largely preserved after excluding BBJ: all 11 lead variants showed concordant effect directions, and 10 remained genome-wide significant.

#### Genome-wide genetic correlation

In the LDSC analysis, positive genetic correlations with GC were observed for overall AG (*r*_g_ = 0.5286, SE = 0.1368, P-value = 1.0 × 10^−4^), overall SAG (*r*_g_ = 0.4359, SE = 0.1834, −3 P-value = 0.018), seropositive AG (*r*_g_ = 0.4474, SE = 0.1616, P-value = 5.6 × 10^-3^), and seropositive SAG (*r*_g_ = 0.4921, SE = 0.2073, P-value = 0.018) (**Supplementary Table 13**). The correlations for overall and seropositive AG remained significant after Bonferroni correction; those for overall and seropositive SAG were also nominally significant. On the other hand, no clear genetic correlation was observed for seronegative AG (*r*_g_ = 0.0791, SE = 0.1776, P-value = 0.656) or seronegative SAG (*r*_g_ = −0.1447, SE = 0.1565, P-value = 0.355). These latter findings may reflect biological differences as well as limited statistical power due to fewer seronegative cases and uncertainty in the heritability estimates.

#### Multi-trait analysis

Pairwise MTAG analyses with GC showed increases in mean chi-squared statistics for overall AG, overall SAG, seropositive AG, and seropositive SAG (**Supplementary Table 14**). The maximum GWAS-equivalent sample size increased by 38.9%, 36.8%, 32.6%, and 52.1%, respectively, with the greatest increase observed for seropositive SAG. The estimated Omega correlations were 0.694, 0.514, 0.593, and 0.586, respectively. In contrast, no meaningful increase in mean chi-squared statistics or equivalent sample size was observed for seronegative AG or SAG, and the Omega correlations could not be estimated for these analyses.

In the overall AG-GC analysis, LDSC SNP heritability increased after MTAG from 0.0311 (SE = 0.0051) to 0.0423 (SE = 0.0073) for AG and from 0.0201 (SE = 0.0053) to 0.0308 (SE = 0.0061) for GC. For AG, regional lead P-values decreased after MTAG, indicating stronger evidence of association (**Supplementary Figure 10; Supplementary Table 15**). P-values changed from 1.14 × 10^−17^ to 9.29 × 10^−35^ at *MUC1*/*MTX1* and from 1.95 × 10^−65^ to 2.93 × 10^−94^ at *PSCA*. The *PRKAA1*/*PTGER4* region reached genome-wide significance in MTAG-adjusted AG (P-value = 3.59 × 10^−21^). These results support genetic sharing between AG and GC and are consistent with contributions along the disease continuum.

#### Local genetic correlation

Local genetic correlation analysis identified significant positive correlations between AG and GC in five genomic regions outside the MHC after Bonferroni correction across 218 eligible regions (P-value < 2.29 × 10^−4^). Strong positive correlations were observed in a local genomic region containing *PSCA* (chr8:143,611,462–144,934,288; ρ = 0.937, 95% CI: 0.876–0.999, P-value = 4.30 × 10^−64^) and a region containing *MUC1* and *MTX1* (chr1:154,685,546–156,813,845; ρ = 1.000, 95% CI: 0.907–1.000, P-value = 2.84 × 10^−32^). Additional positive correlations were observed at 3p21.31 (ρ = 1.000, 95% CI: 0.498–1.000, P-value = 8.48 × 10^−6^), at 5p13.1 in an interval containing *PTGER4*, *PRKAA1*, and *CARD6* (ρ = 0.844, 95% CI: 0.489–1.000, P-value = 2.26 × 10^−5^), and at 8q24.3 in an interval containing *TSNARE1* and overlapping *ADGRB1* (chr8:142,611,890–143,611,461; ρ = 0.853, 95% CI: 0.465–1.000, P-value = 1.42 × 10^−4^). The latter interval was adjacent to, but distinct from, the interval containing *PSCA*.

The 3p21.31 interval contains genes previously implicated in GC cell invasion and migration (*LZTFL1*) [63], gastric tumorigenesis in mice (*SETD2*) [64], and immune tolerance during *H. pylori* infection in mice (*CCR2*) [65]. The 8q24.3 interval overlaps *ADGRB1* (formerly *BAI1*), which has been implicated in the recognition and engulfment of apoptotic gastric epithelial cells by gastric phagocytes [66].

In separate post hoc analyses of broader MHC intervals, positive local genetic correlations were observed for class I (ρ = 0.819, 95% CI: 0.681–0.963, P-value = 3.23 × 10^−19^), class III (ρ = 0.615, 95% CI: 0.418–0.821, P-value = 1.33 × 10^−7^), and class II (ρ = 0.689, 95% CI: 0.547–0.843, P-value = 2.06 × 10^−15^), all meeting the significance threshold corrected for three tests. The five non-MHC results and the three broader MHC regional estimates are presented in **Supplementary Table 16**. The correlation estimates for the interval containing *MUC1* and *MTX1* and for 3p21.31 reached the upper boundary of the correlation scale and should not be interpreted as establishing perfect genetic sharing.

#### Genomic structural equation modeling

In the Genomic SEM model, AG was used as the anchor trait, with its loading on the shared factor fixed to 1 and its residual genetic variance constrained to 0. The loading of the shared genetic factor on GC was 0.5267 (SE = 0.1372, P-value = 1.24 × 10^−4^), corresponding to a model-dependent estimate that the AG-anchored shared component captured 27.7% of GC SNP genetic variance (**Figure 3**). The model converged to a numerically stable estimate. This proportion describes genetic sharing under the specified model and does not quantify a causal contribution of AG to GC.

**Figure 3.**
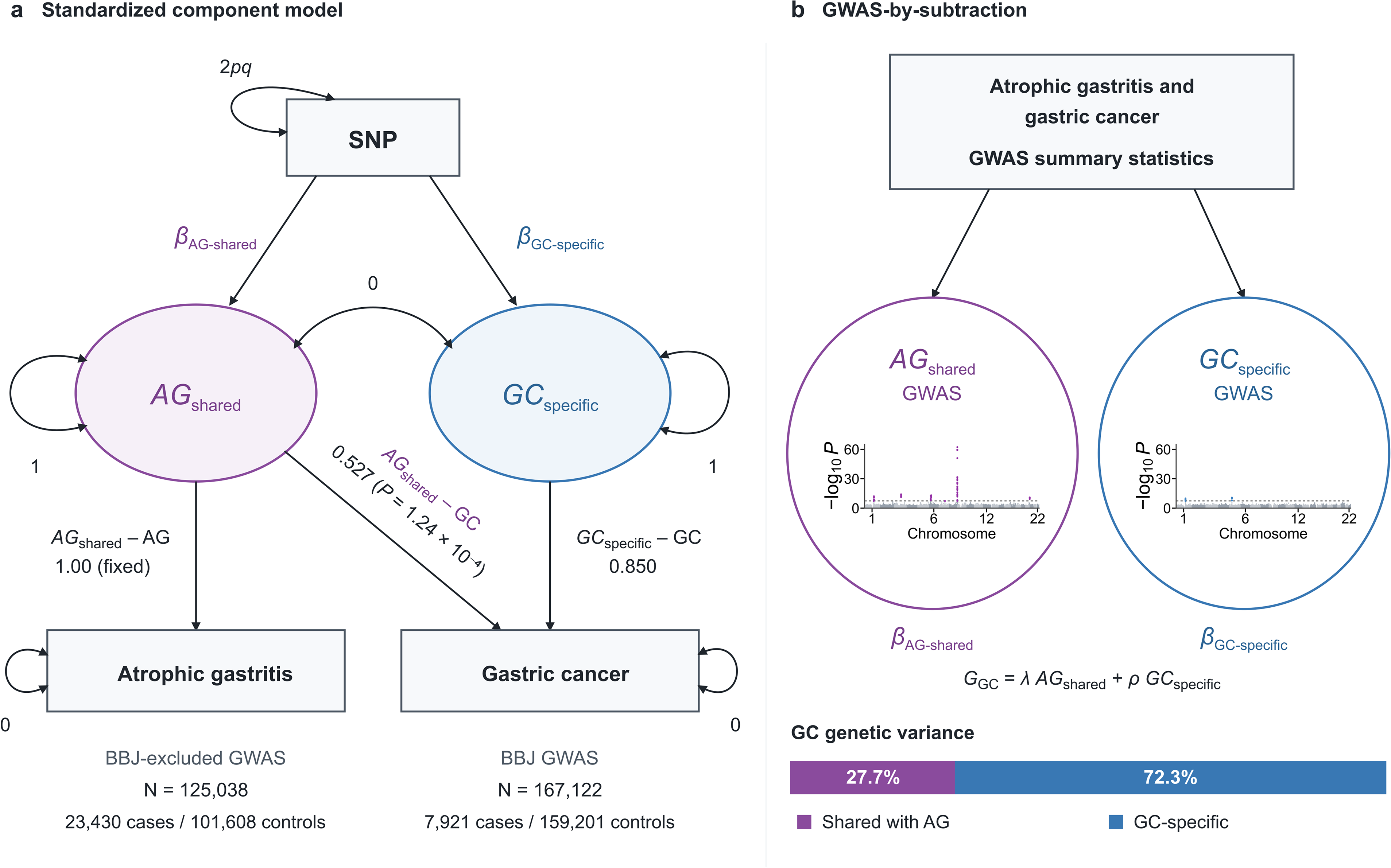
Genomic SEM model of the shared genetic architecture of atrophic gastritis and gastric cancer. (a) Standardized representation of the fitted two-trait Genomic SEM model. BBJ-excluded AG GWAS summary statistics (N = 125,038) and BBJ gastric cancer GWAS summary statistics (7,921 cases and 159,201 controls; total N = 167,122) were used to avoid sample overlap. AG was specified as the anchor trait: its loading on the shared genetic factor *F* was fixed to 1 and its residual genetic variance was constrained to 0. Gastric cancer was modeled as G_GC_ = λ × *F* + ε_GC_. In the standardized diagram, *F* is labeled AG-shared and ε_GC_ = ρ × GC-specific, with orthogonal unit-variance components; ρ = 0.850 is the square root of the GC-specific variance (0.723). The model converged stably. The estimated loading of *F* on gastric cancer was 0.5267 (SE = 0.1372, *P* = 1.24 × 10^−4^). Under the standardized variance parameterization, *F* explained 0.277 (27.7%) of gastric cancer genetic variance, leaving a model-defined GC-specific residual component of 0.723 (72.3%). These percentages describe genome-wide variance decomposition, not the proportions of genome-wide significant loci. SNP variance is 2*pq*, where *p* is an allele frequency and *q* = 1 - *p*. (b) GWAS-by-subtraction. The fitted model was applied at the SNP level to estimate association with the AG-anchored factor *F* (shared component GWAS) and association with the residual gastric cancer component after accounting for λ *F* (GC-specific component GWAS). The shared component is AG-anchored rather than a symmetric intersection of AG and gastric cancer, and the GC-specific component should not be interpreted as variants completely unrelated to AG. In the component Manhattan plots, the horizontal dashed lines indicate *P* = 5 × 10^−8^. Component GWAS results are shown in Supplementary Figure 11 and Supplementary Table 17.

#### GWAS-by-subtraction

GWAS-by-subtraction identified nine genome-wide significant loci associated with the AG-GC shared component (**Supplementary Figure 11; Supplementary Table 17**). These included the *PSCA* 3-prime untranslated region variant rs2976394 (β = 0.2796, SE = 0.0167, P-value = 5.21 × 10^−63^), the *THBS3* intronic variant rs2075571 at *MUC1*/*MTX1* (β = 0.1496, SE = 0.0208, P-value = 5.60 × 10^−13^), and the *CCDC80*-*RP11-572C15.3* intergenic variant rs1249910 (β = −0.1280, SE = 0.0164, P-value = 4.98 × 10^−15^). Further shared-component associations were observed for the *HLA-DQA1*-*DQB1* intergenic variant rs17843604 (β = 0.1179, SE = 0.0161, P-value = 2.32 × 10^−13^), the *USP49* intronic variant rs11753329 (β = 0.1299, SE = 0.0174, P-value = 7.64 × 10^−14^), and the *BMP2*-*CASC20* intergenic variant rs6054471 (β = 0.1272, SE = 0.0187, P-value = 9.67 × 10^−12^).

Two loci showed associations with the GC-specific component: the *PRKAA1* intronic variant rs13361707 (β = −0.1359, SE = 0.0202, P-value = 1.72 × 10^−11^) and the *THBS3* intronic variant rs2075571 (β = 0.1658, SE = 0.0257, P-value = 1.13 × 10^−10^). The GC-specific component represents residual GC genetic variation after accounting for the component shared with AG. Here, “GC-specific” is a model-based designation and does not imply that the underlying genetic effects are unrelated to AG. These effects may contribute to gastric atrophy itself, the transition from atrophy to cancer, or subsequent tumor progression.

### Cross-ancestry meta-analysis

In the cross-ancestry analysis incorporating chronic gastritis GWAS from FinnGen, UK Biobank, and MVP, the major associations identified in the Japanese GWAS were generally maintained without marked attenuation (**Figure 4; Supplementary Figure 12; Supplementary Table 18**). An additional genome-wide significant association was identified for rs2186377 near *TPTEP1* in the analysis combining seronegative SAG with external chronic gastritis data (chr22:17,128,707; beta_0 = 1.316, StdErr_0 = 0.259, P-value = 3.79 × 10^−8^; heterogeneity P-value = 0.064). Here, beta_0 and StdErr_0 denote the intercept effect estimate and its SE in the meta-regression model.

**Figure 4.**
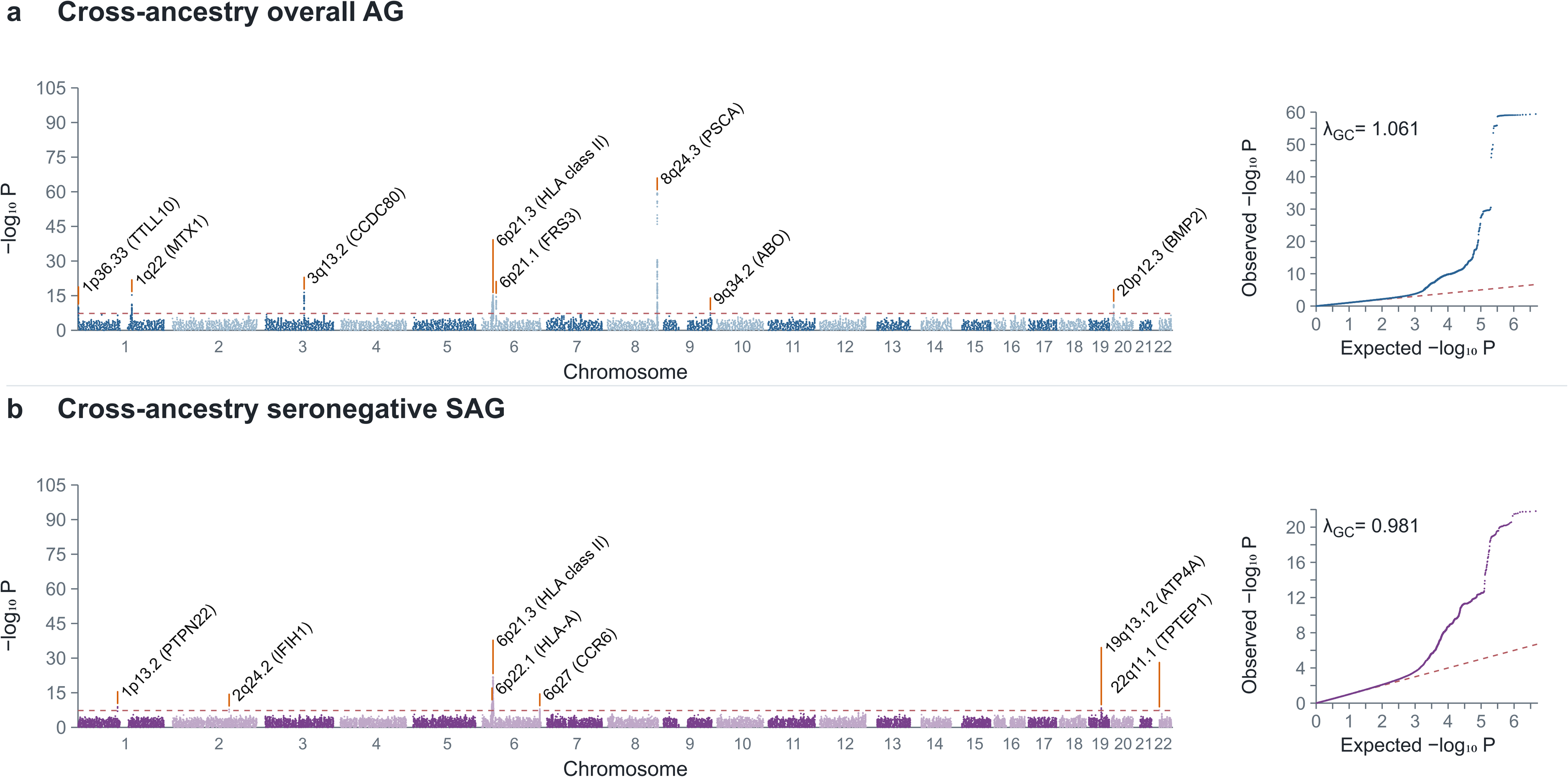
Cross-ancestry meta-analyses integrating Japanese AG/SAG and external chronic gastritis GWAS. Manhattan and quantile-quantile plots show MR-MEGA results obtained by integrating Japanese GWAS summary statistics with chronic gastritis GWAS summary statistics from FinnGen, UK Biobank, and European and African American or Afro-Caribbean MVP datasets: (a) overall AG, including BBJ in the Japanese input, and (b) anti-*H. pylori* IgG-seronegative SAG. The external datasets comprised 1,357,053 participants. Manhattan plots show −log_10_(*P*) values by genomic position, and the red dashed line indicates *P* = 5 × 10^−8^. Selected genome-wide significant loci are annotated. Genomic positions are based on GRCh37. Anti-*H. pylori* IgG seronegativity was defined as <=3 U/mL and does not directly establish absence of current or previous infection. Because the external phenotype was chronic gastritis rather than serologically defined AG/SAG, these results were treated as supportive cross-ancestry evidence. The λ_GC_ values were 1.061 and 0.981 for panels a and b, respectively. λ_GC_ was calculated from the median chi-square statistic using all reported *P* values in the corresponding source summary-statistics file.

*TPTEP1* is a *TPTE*-derived pseudogene expressed as a non-protein-coding long noncoding RNA. Previous studies have reported tumor-suppressive roles mediated through microRNAs and RNA-binding proteins [67,68], although its direct function in AG remains unclear. The interpretation of this association requires consideration of the external phenotype, which was chronic gastritis rather than AG/SAG. Accordingly, it belongs to a different evidential tier from the 17 principal Japanese loci and was interpreted cautiously as an additional locus from an external supportive analysis of the Japanese AG/SAG GWAS.

*PTPN22* rs2476601 also reached genome-wide significance in several cross-ancestry analyses. *PTPN22* encodes a tyrosine phosphatase that regulates lymphocyte antigen receptor signaling and immune tolerance [69]. This signal was also detected by MR-MEGA in Japanese datasets alone, suggesting that its detection may reflect differences from fixed-effect meta-analysis rather than the addition of other ancestry groups.

## Discussion

To our knowledge, we performed the largest GWAS meta-analysis of AG/SAG to date. Associations were identified at 17 principal regions in the Japanese analyses, with an additional *TPTEP1* region identified in the supportive cross-ancestry analysis. By focusing on gastric atrophy as a precursor to GC, this study provides insights into genetic susceptibility at an earlier stage of disease that are difficult to obtain from GWAS of GC alone. In particular, the associations shared with GC, together with the differences by anti-*H. pylori* IgG serostatus, help clarify the biological pathways linking gastric atrophy to the development of GC.

The regions identified in this study included previously reported GC susceptibility loci, such as *PSCA*, *MUC1*/*MTX1*, *PRKAA1*/*PTGER4*, *ABO*, and MHC. *PSCA* has been associated with gastric epithelial biology and GC susceptibility. Its association with AG/SAG suggests that GC-related epithelial susceptibility may act before cancer develops [8,12,13]. *MUC1* encodes a membrane-bound mucin involved in the gastric mucus barrier and mucosal defense against *H. pylori* [9,70]. The *MTX1* variant rs760077 has been reported to be in strong LD with *MUC1* rs4072037 in a Taiwanese population (*r*^2^ = 0.965), supporting the interpretation that the 1q22 signal is related to *MUC1*, although this does not determine which gene or variant is causal [54]. *PTGER4* provides a plausible link to inflammation, mucosal repair, and immune responses, whereas *PRKAA1* provides a plausible link to cellular energy metabolism and stress responses [71,72]. *ABO* encodes glycosyltransferase activity that determines blood group antigens and may influence gastric mucosal glycans, *H. pylori*-related susceptibility, and atrophy [42,62,73]. Within the MHC, class II molecules play a central role in antigen presentation and may contribute to differences in immune responses in both *H. pylori*-related and autoimmune gastritis [74]. In addition, the associations observed in the class I and class III/*CFB* regions suggest possible involvement of cellular immune responses and complement, alongside class II antigen presentation [74,75].

Candidate genes at several additional regions identified in our analyses are involved in immune regulation, epithelial maintenance, and cellular differentiation. The *TTLL10*/*TNFRSF18* (*GITR*) region provides a candidate link to T-cell activation and mucosal immune regulation [76]. *GNG12* encodes a gamma subunit of heterotrimeric G proteins and participates in G protein-coupled receptor signaling and cellular and immune responses [77,78]. *PRDM1* regulates B- and T-cell differentiation and inflammatory responses, including terminal lymphocyte differentiation [79]. In addition, BMP2 signaling contributes to homeostasis at gastric epithelial transition zones and tumor suppression [80]. *CCDC80*, also known as *DRO1*, has been linked to suppression of gastrointestinal cancer cell growth and downregulation in response to oncogenic signaling. These findings make it a candidate for epithelial growth regulation and tissue responses [81]. The *LRRC34* region has been associated with telomere length, suggesting its potential relevance to cellular senescence and mucosal maintenance [82]. *CPEB4* regulates translation, proliferation, and differentiation [83], while *LMTK2* has been implicated in cell-cycle and beta-catenin-related pathways [84].

Both genes are therefore candidates for epithelial responses in atrophic mucosa. *PGC* encodes pepsinogen C, which is expressed in the stomach and is biologically connected to the serum PG-based classification of AG/SAG [85]. Because AG/SAG was primarily defined using serum PG, the association at this region may partly reflect genetic influences on PG II levels and the PG I/II ratio used for case classification, in addition to potential effects on gastric atrophy [86].

The functions of candidate genes at regions associated with anti-*H. pylori* IgG seronegative SAG are consistent with an autoimmune-like component of atrophy. *IFIH1* mediates innate immune recognition of viral RNA [87], *FAP* is associated with fibroblast activation and tissue remodeling [88], and *CCR6* contributes to the migration of immune cells, including Th17 cells [89]. The gastric parietal cell H+/K+-ATPase subunit encoded by *ATP4A* is involved in acid secretion and is a major autoantigen in autoimmune gastritis and pernicious anemia [90]. The roles of these candidate genes in serostatus-dependent atrophy warrant further investigation.

In seronegative AG/SAG, prominent associations were observed in the MHC class II region and for *HLA-DRB1*15:01* and *HLA-DQB1*06:02*. The relationships of these signals with immune-mediated conditions, such as multiple sclerosis and narcolepsy, provide context for considering antigen presentation and immune regulation [60,61]. Together, these findings suggest that serologically defined atrophy may encompass different pathological components rather than a single uniform precancerous phenotype.

Seronegative gastric atrophy is also relevant to GC risk: a prospective Japanese study showed that negative *H. pylori* serology does not preclude elevated risk when pepsinogen-defined atrophy is present [26]. GC has also been documented in *H. pylori*-naïve patients with autoimmune gastritis, although the magnitude of risk in this setting remains uncertain [91,92]. The immune-related associations identified here provide a basis for investigating whether distinct pathways underlying seronegative atrophy also contribute to gastric carcinogenesis. Integrating genetic findings with infection and eradication histories, gastric autoantibodies, and histology could help distinguish these pathways.

In seropositive AG/SAG, associations at *PSCA* and *MUC1*/*MTX1* were prominent, consistent with contributions from epithelial susceptibility and mucus barrier responses in the context of host–pathogen interactions. The effects of *PSCA* were directionally consistent across strata, indicating that its association was not restricted to one serostatus group. On the other hand, the positive *MUC1*/*MTX1* interaction for SAG suggests a stronger role for mucosal defense or epithelial responses in seropositive severe atrophy.

Chinese GC studies have reported interactions with *H. pylori* serostatus at *MUC1* rs4072037 (P-value = 4.9 × 10⁻⁵) [93] and *PSCA* rs2294008 (interaction OR ratio = 3.05, 95% CI: 1.43–6.53; P-value = 0.004) [94], although evidence for the *MUC1* interaction varied across studies. In our SAG analysis, a positive interaction was observed at *MUC1*/*MTX1* rs760077 (P-value = 7.45 × 10⁻¹⁰), whereas *PSCA* rs2920280 showed stronger evidence for a conditional genetic main effect (P-value = 2.32 × 10⁻¹⁷) than for interaction (P-value = 2.63 × 10⁻³). Our analyses extend this line of investigation to AG/SAG, providing insight into serostatus-dependent genetic associations within the broader disease continuum from gastric inflammation through atrophy to GC.

Gene–environment analyses can reveal context-dependent effects at established GWAS loci, as illustrated by our *MUC1*/*MTX1* and HLA findings, in addition to identifying loci with weak marginal associations. Integrating these context-dependent effects with marginal associations may also advance our understanding of the potential contribution of gene–environment interactions to missing heritability [95–97].

Complementary analyses provided evidence of genetic sharing between AG/SAG and GC, with clearer sharing for overall and seropositive AG/SAG in the LDSC and MTAG analyses. LAVA and Genomic SEM further supported genetic sharing between AG and GC. The repeated identification of *PSCA* and *MUC1*/*MTX1* across these analyses suggests that some GC susceptibility regions contribute at the precursor stage.

GWAS-by-subtraction identified both shared and residual GC components. The *PTGER4*/*PRKAA1* region was associated with SAG and with the residual GC component. Possible roles in advanced atrophy or subsequent progression remain hypotheses requiring further investigation. These analyses do not establish when, or through which mechanisms, the associated variants act.

The Kyoto consensus recognizes *H. pylori* gastritis as an infectious disease and places infection diagnosis and eradication in the context of GC prevention [5]. Evidence from long-term randomized trials and syntheses of trial findings supports a preventive effect of eradication on GC [98–100]. Although eradication was not directly evaluated in our study, our findings provide complementary host genetic support for the biological rationale of GC prevention strategies, including *H. pylori* eradication.

Several limitations of this study should be considered. First, AG and SAG were primarily defined using serum PG rather than endoscopic or histological findings, whereas BBJ used medically diagnosed AG. These differences may introduce phenotypic heterogeneity. Second, anti-*H. pylori* IgG does not distinguish active infection, past infection, prior eradication, and low-titer states. Seropositivity should therefore not be equated directly with current infection. In particular, seronegative AG/SAG may include both autoimmune gastritis and advanced *H. pylori*-related atrophy with spontaneous loss of detectable antibodies [101]. Host genetic influences on antibody responses may also affect serostatus classification and thereby influence the stratified associations [102]. Differences in anti-*H. pylori* IgG assays across cohorts may also have contributed to heterogeneity in serostatus classification. Third, the external cross-ancestry datasets assessed chronic gastritis rather than the same serological AG/SAG phenotypes, providing supportive rather than directly equivalent evidence. Fourth, the complex LD structure of the MHC limits the ability to determine the independence of locus- and HLA allele-level associations. Fifth, the interaction analyses were restricted to unrelated TMM participants, which may limit generalisability, and the small numbers of seronegative cases, particularly SAG cases, may have reduced statistical power and precision.

In conclusion, our findings characterize the heterogeneous genetic basis of AG/SAG. Anti-*H. pylori* IgG seropositive disease was characterized by epithelial and mucus barrier-related GC susceptibility regions, whereas prominent HLA class II and immune-related associations were observed in seronegative disease. In addition, genetic sharing with GC places AG within a broader disease continuum. These results provide a basis for future genetic risk stratification and personalized approaches to prevention.

## Data availability

GWAS summary statistics generated in this study will be made available through the jMorp database (https://jmorp.megabank.tohoku.ac.jp/) upon publication of this work, subject to the data-sharing policies of the participating cohorts. Individual-level genotype and phenotype data from the participating Japanese cohorts are not publicly available because of privacy, ethical, and institutional restrictions. Access to these data is subject to the consent conditions and approval procedures of the respective cohorts.

Publicly available gastric cancer GWAS summary statistics from BioBank Japan were obtained from PheWeb.jp (https://pheweb.jp/). Chronic gastritis GWAS summary statistics were obtained from FinnGen Release 12 (https://finngen.gitbook.io/documentation/data-download), the Neale Lab UK Biobank resource (https://www.nealelab.is/uk-biobank/), and the Million Veteran Program (https://phenomics.va.ornl.gov/web/). Access to these external datasets is governed by the terms of the respective resources.

## Funding

This work was supported by Programs for Bridging the gap between R&D and the IDeal society (society 5.0) and Generating Economic and social value (BRIDGE; JPJ012525 to G.T.) and the Japan Society for the Promotion of Science (JSPS) KAKENHI (JP25K00278 to T.K.). Additional support was provided by the RIKEN Center for Advanced Intelligence Project through the project “Genome Statistical Analysis Using Machine Learning and Artificial Intelligence Technologies” (to G.T.) and by the Tohoku Kaihatsu Memorial Foundation (to T.K.). Y.O. was supported by JSPS KAKENHI (25H01057), the Japan Agency for Medical Research and Development (AMED; JP223fa627001, JP223fa627010, JP223fa627011, JP22zf0127008, JP23tm0524002, JP24wm0625504, JP24gm1810011, JP25kk0305022, JP26tm0135237, JP26ek0410158), the Japan Science and Technology Agency (JST) Moonshot R&D Program (JPMJMS2021, JPMJMS2024), Ono Pharmaceutical Foundation for Oncology, Immunology, and Neurology, Baelz Research Grant, and the RIKEN TRIP initiative (AGIS). T.O. was supported by AMED (JP25tm042423), JSPS KAKENHI (JP25K23789, 26K20570), and the Takeda Science Foundation. K. Matsuda was supported by AMED (JP23tm0624002, JP22zf0127009, JP223fa627011, JP24gm2010003, and JP25ama221544) and JST AI for Science (JPMJAR2607). K. Matsuda and C.T. were supported by AMED (JP26ck0106091 and JP23ck0106877) and the Takeda Science Foundation. C.T. was supported by the Hirose Foundation.

The J-MICC Study was supported by Grants-in-Aid for Scientific Research from the Ministry of Education, Culture, Sports, Science and Technology (MEXT) (Priority Areas of Cancer: 17015018; Innovative Areas: 221S0001) and JSPS KAKENHI (JP16H06277 and JP22H04923[CoBiA]). It was also partially funded by the BBJ Project from AMED (since April 2015) and by MEXT (from April 2003 to March 2015).

The Tohoku Medical Megabank Project was supported by AMED and MEXT (grant number: JP26tm0235007). Computational resources were provided by the Tohoku Medical Megabank Organization supercomputer system, which is supported by AMED (grant number: JP26tm0135602).

The Iwate Tohoku Medical Megabank Organization (IMM) was supported by AMED under Grant Numbers JP15km0105004, JP21tm0124006, and JP26tm0235008.

The HERPACC Study was supported by Grants-in-Aid for Scientific Research from MEXT (Priority Areas of Cancer: 17015018; Innovative Areas: 221S0001) and JSPS KAKENHI (JP16H06277[CoBiA], JP26253041, JP20K10463, JP23K16316, and JP24K02697), a Grant-in-Aid for the Third Term Comprehensive 10-year Strategy for Cancer Control from the Ministry of Health, Labour and Welfare (MHLW) of Japan, as well as the Aichi Cancer Center Joint Research Project (1st, 2nd, and 3rd) on Priority Areas.

This work was also supported by the National Cancer Center Research and Development Fund (28-A-19, 31-A-18, 2022-A-20, and 2025-A-23). The JPHC Study was supported by the National Cancer Center Research and Development Fund (23-A-31[toku], 26-A-2, 29-A-4, 2020-J-4, 2023-J-4, and 2026-J-4), and Grants-in-Aid for Cancer Research from MHLW of Japan (from 1989 to 2010).

## Acknowledgements

We thank the participants, investigators, and staff of the Tohoku Medical Megabank Project, including ToMMo and IMM, the Yamagata Study, the JPHC study, the J-MICC Study, HERPACC, and BioBank Japan for their contributions to this study. We also thank the participants and investigators of FinnGen, UK Biobank, and the Million Veteran Program for their contributions to the publicly available GWAS resources used in this study.

The ToMMo members are listed at https://www.megabank.tohoku.ac.jp/english/a250901/ (as of September 1, 2025). The current and past members of the JPHC Study Group are listed at https://epi.ncc.go.jp/en/jphc/781/9509.html (2024 list). Contributors to the J-MICC Study are listed at https://jmicc.com/en/contributors (updated December 20, 2025).

## Author contributions

T.K. performed the statistical analyses and drafted the manuscript. A.N. and G.T. provided scientific guidance during the study and manuscript preparation.

A.M., Y. Kotsar, S.M., M.S., J.T., C.G., T.S., A.K.K.M., K.H., T.F., H.O., and M.M. contributed to bioinformatics analyses. A. Hozawa, K.K., and S.K. contributed to the management of ToMMo data. A.S., Y. Sutoh, Y. Otsuka-Yamasaki, and K.T. contributed to the management of IMM data. Y. Sasaki, Y.A., O.N., and Y.U. contributed to the management of Yamagata Study data. T.Y. contributed to bioinformatics analyses of JPHC data, and S. Nakano, N.S., and M.I. contributed to the management of JPHC data. M.N. contributed to bioinformatics analyses of J-MICC and HERPACC data. T.N., A. Hishida, N.T., Y. Kasugai, and K.W. contributed to the management of J-MICC data. Y.N.K., S.Y., H.I., and K. Matsuo contributed to the management of HERPACC data. Y.H. contributed to bioinformatics analyses of BBJ data. C.T., M.K., Y. Kamatani, and K. Matsuda contributed to the management of BBJ data.

T.K., A.N., and G.T. contributed to study design. A.N., T.O., S. Namba, J.K.K., K. Matsuo, Y. Okada, and G.T. contributed to interpretation and critical revision of the manuscript.

## Competing interests

The authors declare no competing interests.

## Notes

### Competing Interest Statement

The authors have declared no competing interest.

### Author Declarations

The institutional review board of Tohoku University gave ethical approval for the Tohoku Medical Megabank Project. The institutional review board of the National Cancer Center gave ethical approval for the Japan Public Health Center-based Prospective Study. The institutional review board of Aichi Cancer Center gave ethical approval for the Japan Multi-Institutional Collaborative Cohort Study and the Hospital-based Epidemiologic Research Program at Aichi Cancer Center. The institutional review board of Yamagata University gave ethical approval for the Yamagata Study. The ethics committees of the Institute of Medical Science, the University of Tokyo, and the RIKEN Center for Integrative Medical Sciences gave ethical approval for BioBank Japan. Informed consent was obtained from all participants.

## References

1. Correa P, Piazuelo MB. The gastric precancerous cascade. J Dig Dis. 2012;13(1):2–9.

2. Correa P. Human gastric carcinogenesis: a multistep and multifactorial process--First American Cancer Society Award Lecture on Cancer Epidemiology and Prevention. Cancer Res. 1992;52:6735–6740.

3. Park YH, Kim N. Review of atrophic gastritis and intestinal metaplasia as a premalignant lesion of gastric cancer. J Cancer Prev. 2015;20:25–40.

4. Pimentel-Nunes P, Libânio D, Marcos-Pinto R, et al. Management of epithelial precancerous conditions and lesions in the stomach (MAPS II): European Society of Gastrointestinal Endoscopy guideline update. Endoscopy. 2019;51:365–388.

5. Sugano K, Tack J, Kuipers EJ, et al. Kyoto global consensus report on *Helicobacter pylori* gastritis. Gut. 2015;64:1353–1367.

6. Uemura N, Okamoto S, Yamamoto S, et al. *Helicobacter pylori* infection and the development of gastric cancer. N Engl J Med. 2001;345:784–789.

7. Kyosaka T, Narita A, Kulski JK, et al. Genome-wide association and Mendelian randomization analyses link *Helicobacter pylori* infection to Human Leukocyte Antigen polymorphisms and autoimmune diseases. medRxiv. 2026;2026.03.27.26349357.

8. Sakamoto H, Yoshimura K, Saeki N, et al. Genetic variation in PSCA is associated with susceptibility to diffuse-type gastric cancer. Nat Genet. 2008;40:730–740.

9. Saeki N, Saito A, Choi IJ, et al. A functional single nucleotide polymorphism in MUC1, at chromosome 1q22, determines susceptibility to diffuse-type gastric cancer. Gastroenterology. 2011;140:892–902.

10. Tanikawa C, Kamatani Y, Toyoshima O, et al. Genome-wide association study identifies gastric cancer susceptibility loci at 12q24.11-12 and 20q11.21. Cancer Sci. 2018;109:4015–4024.

11. Hu N, Wang Z, Song X, et al. Genome-wide association study of gastric adenocarcinoma in Asia: a comparison of associations between cardia and non-cardia tumours. Gut. 2016;65:1611–1618.

12. Hishida A, Ugai T, Fujii R, et al. GWAS analysis reveals a significant contribution of PSCA to the risk of *Helicobacter pylori*-induced gastric atrophy. Carcinogenesis. 2019;40:661–668.

13. Kupcinskas J, Wex T, Link A, et al. PSCA and MUC1 gene polymorphisms are associated with gastric cancer and pre-malignant gastric conditions. Anticancer Res. 2014;34:7167–7175.

14. Hozawa A, et al. Study profile of the Tohoku Medical Megabank Community-Based Cohort Study. J Epidemiol. 2021;31:65–76.

15. Kuriyama S, et al. The Tohoku Medical Megabank Project: design and mission. J Epidemiol. 2016;26:493–511.

16. Kuriyama S, et al. Cohort profile: Tohoku Medical Megabank Project Birth and Three-Generation Cohort Study (TMM BirThree Cohort Study): rationale, progress and perspective. Int J Epidemiol. 2020;49:18–19.

17. Saito T, et al. Factors associated with community residents’ preference for living at home at the end of life: The Yamagata Cohort Survey. Glob Health Med. 2024;6:70–76.

18. Tsugane S, Sawada N. The JPHC study: design and some findings on the typical Japanese diet. Jpn J Clin Oncol. 2014;44:777–782.

19. Takeuchi K, et al. Study profile of the Japan Multi-institutional Collaborative Cohort (J-MICC) Study. J Epidemiol. 2021;31:660–668.

20. Tajima K, et al. A model of practical cancer prevention for out-patients visiting a hospital: the Hospital-based Epidemiologic Research Program at Aichi Cancer Center (HERPACC). Asian Pac J Cancer Prev. 2000;1:35–47.

21. Nagai A, et al. Overview of the BioBank Japan Project: Study design and profile. J Epidemiol. 2017;27:S2–S8.

22. Kurki MI, Karjalainen J, Palta P, et al. FinnGen provides genetic insights from a well-phenotyped isolated population. Nature. 2023;613:508–518.

23. Sudlow C, Gallacher J, Allen N, et al. UK Biobank: an open access resource for identifying the causes of a wide range of complex diseases of middle and old age. PLoS Med. 2015;12:e1001779.

24. Gaziano JM, Concato J, Brophy M, et al. Million Veteran Program: a mega-biobank to study genetic influences on health and disease. J Clin Epidemiol. 2016;70:214–223.

25. Miki K. Gastric cancer screening by combined assay for serum anti-*Helicobacter pylori* IgG antibody and serum pepsinogen levels--“ABC method”. Proc Jpn Acad Ser B Phys Biol Sci. 2011;87:405–414.

26. Watabe H, Mitsushima T, Yamaji Y, et al. Predicting the development of gastric cancer from combining *Helicobacter pylori* antibodies and serum pepsinogen status: a prospective endoscopic cohort study. Gut. 2005;54:764–768.

27. Inoue M, Sawada N, Goto A, Shimazu T, Yamaji T, Iwasaki M, Tsugane S; JPHC Study Group. High-negative anti-*Helicobacter pylori* IgG antibody titers and long-term risk of gastric cancer: results from a large-scale population-based cohort study in Japan. Cancer Epidemiol Biomarkers Prev. 2020;29:420–426.

28. Sakurai-Yageta M, Kumada K, Gocho C, et al. Japonica Array NEO with increased genome-wide coverage and abundant disease risk SNPs. J Biochem. 2021;170(3):399–410.

29. 1000 Genomes Project Consortium. A global reference for human genetic variation. Nature. 2015;526:68–74.

30. Tadaka S, Katsuoka F, Ueki M, et al. 3.5KJPNv2: an allele frequency panel of 3552 Japanese individuals including the X chromosome. Hum Genome Var. 2019;6:28.

31. Delaneau O, Zagury JF, Marchini J. Improved whole-chromosome phasing for disease and population genetic studies. Nat Methods. 2013;10:5–6.

32. Bycroft C, Freeman C, Petkova D, et al. The UK Biobank resource with deep phenotyping and genomic data. Nature. 2018;562:203–209.

33. Yang J, Lee SH, Goddard ME, Visscher PM. GCTA: a tool for genome-wide complex trait analysis. Am J Hum Genet. 2011;88:76–82.

34. Jiang L, Zheng Z, Fang H, Yang J. A generalized linear mixed model association tool for biobank-scale data. Nat Genet. 2021;53:1616–1621.

35. Mbatchou J, Barnard L, Backman J, et al. Computationally efficient whole-genome regression for quantitative and binary traits. Nat Genet. 2021;53:1097–1103.

36. Willer CJ, Li Y, Abecasis GR. METAL: fast and efficient meta-analysis of genomewide association scans. Bioinformatics. 2010;26:2190–2191.

37. Watanabe K, Taskesen E, van Bochoven A, Posthuma D. Functional mapping and annotation of genetic associations with FUMA. Nat Commun. 2017;8:1826.

38. Benner C, Spencer CCA, Havulinna AS, Salomaa V, Ripatti S, Pirinen M. FINEMAP: efficient variable selection using summary data from genome-wide association studies. Bioinformatics. 2016;32:1493–1501.

39. Chang CC, Chow CC, Tellier LC, Vattikuti S, Purcell SM, Lee JJ. Second-generation PLINK: rising to the challenge of larger and richer datasets. Gigascience. 2015;4:7.

40. Naito T, Suzuki K, Hirata J, et al. A deep learning method for HLA imputation and trans-ethnic MHC fine-mapping of type 1 diabetes. Nat Commun. 2021;12:1639.

41. Okada Y, Kim K, Han B, et al. Risk for ACPA-positive rheumatoid arthritis is driven by shared HLA amino acid polymorphisms in Asian and European populations. Hum Mol Genet. 2014;23(25):6916–6926.

42. Yamamoto F, Clausen H, White T, Marken J, Hakomori S. Molecular genetic basis of the histo-blood group ABO system. Nature. 1990;345:229–233.

43. Nakao M, Matsuo K, Hosono S, et al. ABO blood group alleles and the risk of pancreatic cancer in a Japanese population. Cancer Sci. 2011;102(5):1076–1080.

44. Sakaue S, Kanai M, Tanigawa Y, et al. A cross-population atlas of genetic associations for 220 human phenotypes. Nat Genet. 2021;53:1415–1424.

45. Bulik-Sullivan BK, Loh PR, Finucane HK, et al. LD Score regression distinguishes confounding from polygenicity in genome-wide association studies. Nat Genet. 2015;47:291–295.

46. Bulik-Sullivan B, Finucane HK, Anttila V, et al. An atlas of genetic correlations across human diseases and traits. Nat Genet. 2015;47:1236–1241.

47. Turley P, Walters RK, Maghzian O, et al. Multi-trait analysis of genome-wide association summary statistics using MTAG. Nat Genet. 2018;50:229–237.

48. Werme J, van der Sluis S, Posthuma D, de Leeuw CA. An integrated framework for local genetic correlation analysis. Nat Genet. 2022;54:274–282.

49. Berisa T, Pickrell JK. Approximately independent linkage disequilibrium blocks in human populations. Bioinformatics. 2016;32:283–285.

50. Xie T, Rowen L, Aguado B, et al. Analysis of the gene-dense major histocompatibility complex class III region and its comparison to mouse. Genome Res. 2003;13(12):2621–2636.

51. Grotzinger AD, Rhemtulla M, de Vlaming R, et al. Genomic structural equation modelling provides insights into the multivariate genetic architecture of complex traits. Nat Hum Behav. 2019;3:513–525.

52. Demange PA, Malanchini M, Mallard TT, et al. Investigating the genetic architecture of noncognitive skills using GWAS-by-subtraction. Nat Genet. 2021;53:35–44.

53. Magi R, Horikoshi M, Sofer T, et al. Trans-ethnic meta-regression of genome-wide association studies accounting for ancestry increases power for discovery and improves fine-mapping resolution. Hum Mol Genet. 2017;26:3639–3650.

54. Teng MS, Wu S, Hsu LA, Chou HH, Ko YL. Pleiotropic effects of functional MUC1 variants on cardiometabolic, renal, and hematological traits in the Taiwanese population. Int J Mol Sci. 2021;22(19):10641.

55. Nibbs RJ, Graham GJ. Immune regulation by atypical chemokine receptors. Nat Rev Immunol. 2013;13:815–829.

56. Urrutia R. KRAB-containing zinc-finger repressor proteins. Genome Biol. 2003;4:231.

57. Liu J, Huang C, Peng C, et al. Stromal fibroblast activation protein alpha promotes gastric cancer progression via epithelial-mesenchymal transition through Wnt/β-catenin pathway. BMC Cancer. 2018;18:1099.

58. Ba MC, Ba Z, Gong YF, et al. Knockdown of lncRNA ZNRD1-AS1 suppresses gastric cancer cell proliferation and metastasis by targeting the miR-9-5p/HSP90AA1 axis. Aging (Albany NY). 2021;13(13):17285–17301.

59. Lahner E, Brigatti C, Marzinotto I, et al. Luminescent immunoprecipitation system (LIPS) for detection of autoantibodies against ATP4A and ATP4B subunits of gastric proton pump H+,K+-ATPase in atrophic body gastritis patients. Clin Transl Gastroenterol. 2017;8(1):e215.

60. International Multiple Sclerosis Genetics Consortium. Analysis of immune-related loci identifies 48 new susceptibility variants for multiple sclerosis. Nat Genet. 2013;45:1353–1360.

61. Mignot E, Lin L, Rogers W, et al. Complex HLA-DR and -DQ interactions confer risk of narcolepsy-cataplexy in three ethnic groups. Am J Hum Genet. 2001;68:686–699.

62. Nakao M, Matsuo K, Ito H, et al. ABO genotype and the risk of gastric cancer, atrophic gastritis, and *Helicobacter pylori* infection. Cancer Epidemiol Biomarkers Prev. 2011;20(8):1665–1672.

63. Wang L, Guo J, Wang Q, et al. LZTFL1 suppresses gastric cancer cell migration and invasion through regulating nuclear translocation of β-catenin. J Cancer Res Clin Oncol. 2014;140(12):1997–2008.

64. Feng W, Ma C, Rao H, et al. Setd2 deficiency promotes gastric tumorigenesis through inhibiting the SIRT1/FOXO pathway. Cancer Lett. 2023;579:216470.

65. Sun X, Zhang M, El-Zaatari M, Huffnagle GB, Kao JY. CCR2 mediates Helicobacter pylori-induced immune tolerance and contributes to mucosal homeostasis. Helicobacter. 2017;22(2):e12366.

66. Das S, Sarkar A, Ryan KA, et al. Brain angiogenesis inhibitor 1 is expressed by gastric phagocytes during infection with Helicobacter pylori and mediates the recognition and engulfment of human apoptotic gastric epithelial cells. FASEB J. 2014;28(5):2214–2224.

67. Cao F, Wang Z, Feng Y, et al. lncRNA TPTEP1 competitively sponges miR-328-5p to inhibit the proliferation of non-small cell lung cancer cells. Oncol Rep. 2020;43:1606–1618.

68. Feng Y, Zhang Z, Yang H, et al. The lncRNA TPTEP1 suppresses PI3K/AKT signalling and inhibits ovarian cancer progression by interacting with PTBP1. J Cell Mol Med. 2024;28:e70106.

69. Salmond RJ, Brownlie RJ, Morrison VL, et al. The tyrosine phosphatase PTPN22 discriminates weak self peptides from strong agonist TCR signals. Nat Immunol. 2014;15:875–883.

70. McGuckin MA, Every AL, Skene CD, et al. MUC1 mucin limits both *Helicobacter pylori* colonization of the murine gastric mucosa and associated gastritis. Gastroenterology. 2007;133:1210–1218.

71. Sugimoto Y, Narumiya S. Prostaglandin E receptors. J Biol Chem. 2007;282:11613–11617.

72. Hardie DG. AMP-activated protein kinase: maintaining energy homeostasis at the cellular and whole-body levels. Annu Rev Nutr. 2014;34:31–55.

73. Ilver D, Arnqvist A, Ogren J, et al. *Helicobacter pylori* adhesin binding fucosylated histo-blood group antigens revealed by retagging. Science. 1998;279:373–377.

74. Neefjes J, Jongsma ML, Paul P, Bakke O. Towards a systems understanding of MHC class I and MHC class II antigen presentation. Nat Rev Immunol. 2011;11:823–836.

75. Ricklin D, Hajishengallis G, Yang K, Lambris JD. Complement: a key system for immune surveillance and homeostasis. Nat Immunol. 2010;11:785–797.

76. Nocentini G, Ronchetti S, Cuzzocrea S, Riccardi C. GITR/GITRL: more than an effector T cell co-stimulatory system. Eur J Immunol. 2007;37:1165–1169.

77. Hurowitz EH, Melnyk JM, Chen YJ, Kouros-Mehr H, Simon MI, Shizuya H. Genomic characterization of the human heterotrimeric G protein alpha, beta, and gamma subunit genes. DNA Res. 2000;7:111–120.

78. Wettschureck N, Offermanns S. Mammalian G proteins and their cell type-specific functions. Physiol Rev. 2005;85:1159–1204.

79. Kallies A, Nutt SL. Terminal differentiation of lymphocytes depends on Blimp-1. Curr Opin Immunol. 2007;19:156–162.

80. Bleuming SA, He XC, Kodach LL, et al. Bone morphogenetic protein signaling suppresses tumorigenesis at gastric epithelial transition zones in mice. Cancer Res. 2007;67:8149–8155.

81. Bommer GT, Jager C, Durr EM, et al. DRO1, a gene down-regulated by oncogenes, mediates growth inhibition in colon and pancreatic cancer cells. J Biol Chem. 2005;280:7962–7975.

82. Codd V, Nelson CP, Albrecht E, et al. Identification of seven loci affecting mean telomere length and their association with disease. Nat Genet. 2013;45:422–427.

83. Ortiz-Zapater E, Pineda D, Martinez-Bosch N, et al. Key contribution of CPEB4-mediated translational control to cancer progression. Nat Med. 2012;18:83–90.

84. Han X, Wang DZ, Yuan M, Bai WJ. Lemur tyrosine kinase 2 silencing inhibits the proliferation of gastric cancer cells by regulating GSK-3β phosphorylation and β-catenin nuclear translocation. Bioengineered. 2022;13(3):6231–6243.

85. Taggart RT, Cass LG, Mohandas TK, Derby P, Barr PJ, Pals G. Human pepsinogen C (progastricsin). Isolation of cDNA clones, localization to chromosome 6, and sequence homology with pepsinogen A. J Biol Chem. 1989;264:375–379.

86. Yamagata Z, Iijima S, Asaka A, Kobayashi K. Effects of pepsinogen C gene polymorphisms on serum pepsinogen I and serum pepsinogen II levels. Nihon Eiseigaku Zasshi. 1995;49:998–1003.

87. Nejentsev S, Walker N, Riches D, Egholm M, Todd JA. Rare variants of IFIH1, a gene implicated in antiviral responses, protect against type 1 diabetes. Science. 2009;324:387–389.

88. Park JE, et al. Fibroblast activation protein, a dual specificity serine protease expressed in reactive human tumor stromal fibroblasts. J Biol Chem. 1999;274:36505–36512.

89. Reboldi A, Coisne C, Baumjohann D, et al. C-C chemokine receptor 6-regulated entry of TH-17 cells into the CNS through the choroid plexus is required for EAE. Nat Immunol. 2009;10:514–523.

90. Lenti MV, Rugge M, Lahner E, et al. Autoimmune gastritis. Nat Rev Dis Primers. 2020;6:56.

91. Takasu A, Hirasawa T, Higashi Y, et al. Comparison of clinicopathological features of gastric carcinoma in autoimmune gastritis with and without *Helicobacter pylori* infection. Gastric Cancer. 2025;28(6):1182–1189.

92. Rugge M, Bricca L, Guzzinati S, et al. Autoimmune gastritis: long-term natural history in naïve *Helicobacter pylori*-negative patients. Gut. 2023;72(1):30–38.

93. Li M, Huang L, Qiu H, et al. *Helicobacter pylori* infection synergizes with three inflammation-related genetic variants in the GWASs to increase risk of gastric cancer in a Chinese population. PLoS One. 2013;8(9):e74976.

94. Cai M, Dai S, Chen W, et al. Environmental factors, seven GWAS-identified susceptibility loci, and risk of gastric cancer and its precursors in a Chinese population. Cancer Med. 2017;6(3):708–720.

95. Manolio TA, Collins FS, Cox NJ, et al. Finding the missing heritability of complex diseases. Nature. 2009;461(7265):747–753.

96. Thomas D. Gene–environment-wide association studies: emerging approaches. Nat Rev Genet. 2010;11(4):259–272.

97. Herrera-Luis E, Benke K, Volk H, et al. Gene–environment interactions in human health. Nat Rev Genet. 2024;25(11):768–784.

98. Li WQ, Zhang JY, Ma JL, et al. Effects of *Helicobacter pylori* treatment and vitamin and garlic supplementation on gastric cancer incidence and mortality: follow-up of a randomized intervention trial. BMJ. 2019;366:l5016.

99. Yan L, Chen Y, Chen F, et al. Effect of *Helicobacter pylori* eradication on gastric cancer prevention: updated report from a randomized controlled trial with 26.5 years of follow-up. Gastroenterology. 2022;163:154–162.e3.

100. Ford AC, Forman D, Hunt RH, Yuan Y, Moayyedi P. *Helicobacter pylori* eradication therapy to prevent gastric cancer in healthy asymptomatic infected individuals: systematic review and meta-analysis. BMJ. 2014;348:g3174.

101. Kokkola A, Kosunen TU, Puolakkainen P, et al. Spontaneous disappearance of Helicobacter pylori antibodies in patients with advanced atrophic corpus gastritis. APMIS. 2003;111:619–624.

102. Mayerle J, den Hoed CM, Schurmann C, et al. Identification of genetic loci associated with Helicobacter pylori serologic status. JAMA. 2013;309:1912–1920.

