## Supplementary Figures 1-12 for "Genome-wide association meta-analysis of atrophic gastritis reveals anti–*H. pylori* IgG serostatus-dependent associations and genetic overlap with gastric cancer"

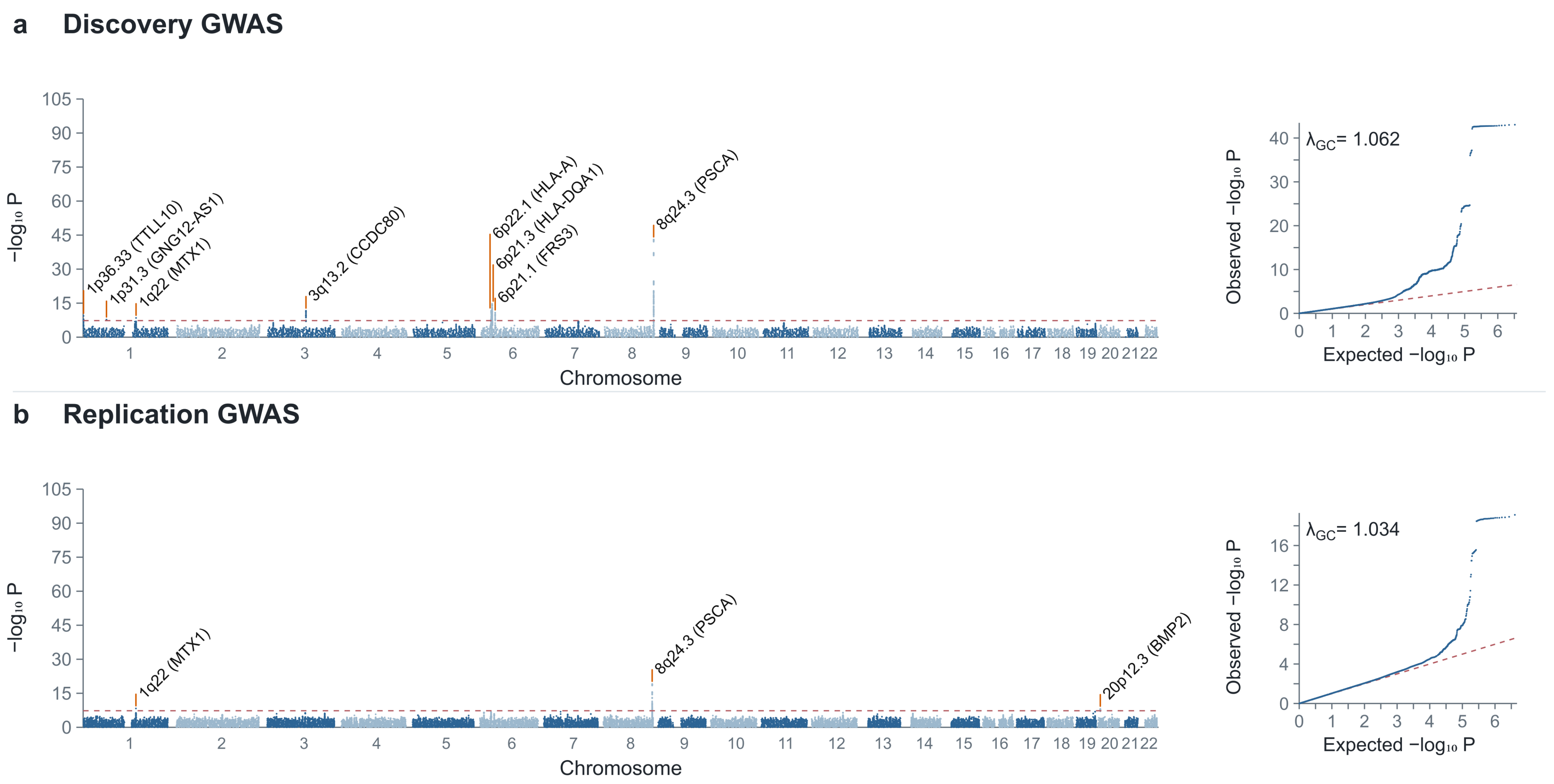
a Discovery GWAS
−log₁₀ P
1p36.33 (TTLL10)
1p31.3 (GNG12-AS1)
1q22 (MTX1)
3q13.2 (CCDC80)
6p22.1 (HLA-A)
6p21.3 (HLA-DQA1)
6p21.1 (FRS3)
8q24.3 (PSCA)
Chromosome
λGC= 1.062
Observed −log₁₀ P
Expected −log₁₀ P
b Replication GWAS
−log₁₀ P
1q22 (MTX1)
8q24.3 (PSCA)
20p12.3 (BMP2)
Chromosome
λGC= 1.034
Observed −log₁₀ P
Expected −log₁₀ P

**Supplementary Figure 1. Manhattan and quantile-quantile plots for the discovery and replication overall AG GWAS.**

Panels show (a) discovery and (b) replication GWAS results. Manhattan plots show $-\log_{10}(P)$ values by genomic position; the red dashed line indicates $P = 5 \times 10^{-8}$. Selected genome-wide significant loci are annotated. Quantile-quantile plots compare observed and expected $-\log_{10}(P)$ values. Genomic positions are based on GRCh37. BBJ was included in the replication meta-analysis. The $\lambda_{GC}$ values were 1.062 and 1.034 for panels a and b, respectively. $\lambda_{GC}$ was calculated from the median chi-square statistic using all reported *P* values in the corresponding source summary-statistics file.

### a Discovery GWAS

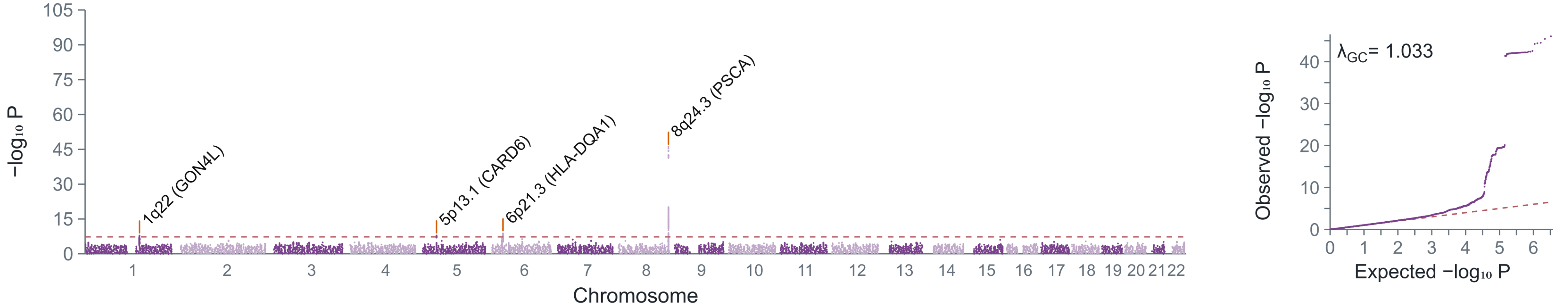


### b Replication GWAS

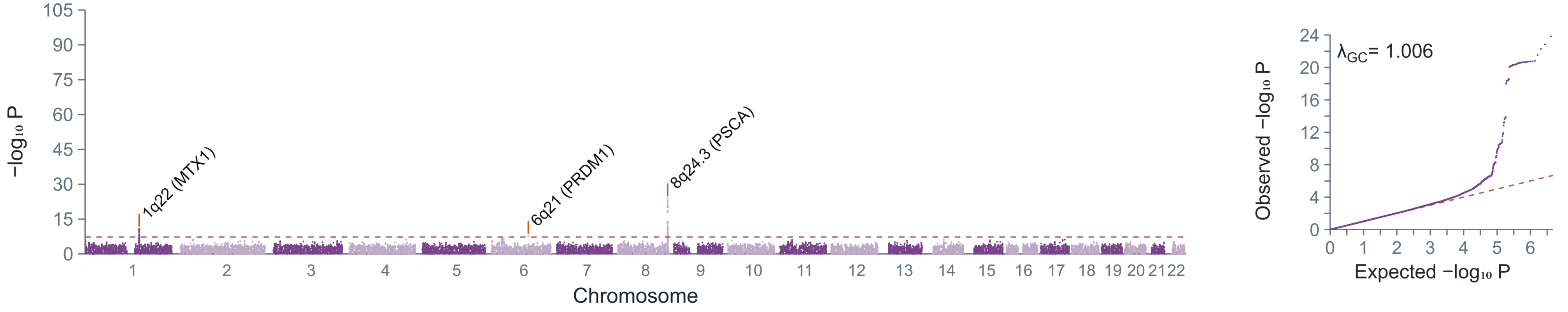

### Supplementary Figure 2. Manhattan and quantile-quantile plots for the discovery and replication overall SAG GWAS

Panels show (a) discovery and (b) replication GWAS results. Manhattan plots show $-\log_{10}(P)$ values by genomic position; the red dashed line indicates $P = 5 \times 10^{-8}$. Selected genome-wide significant loci are annotated. Quantile-quantile plots compare observed and expected $-\log_{10}(P)$ values. Genomic positions are based on GRCh37. The $\lambda_{GC}$ values were 1.033 and 1.006 for panels a and b, respectively. $\lambda_{GC}$ was calculated from the median chi-square statistic using all reported $P$ values in the corresponding source summary-statistics file.

Supplementary Figure 3

### a Discovery GWAS

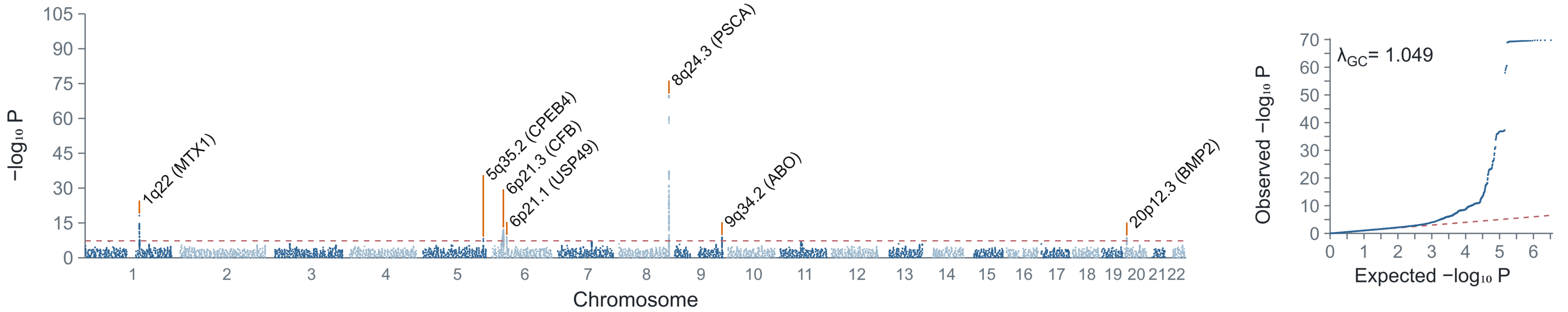


### b Replication GWAS

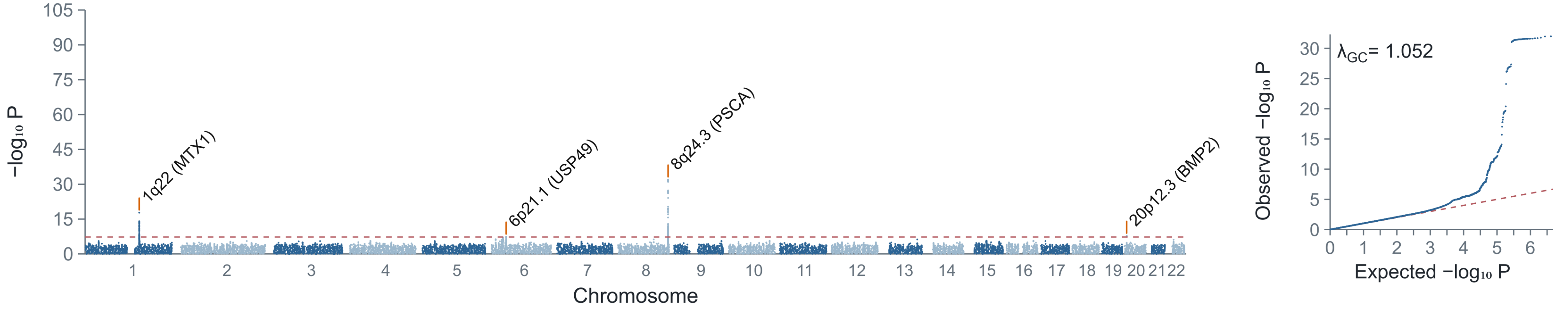

**Supplementary Figure 3. Manhattan and quantile-quantile plots for the discovery and replication anti-*H. pylori* IgG-seropositive AG GWAS.**

Panels show (a) discovery and (b) replication GWAS results. Manhattan plots show $-\log_{10}(P)$ values by genomic position; the red dashed line indicates $P = 5 \times 10^{-8}$. Selected genome-wide significant loci are annotated. Quantile-quantile plots compare observed and expected $-\log_{10}(P)$ values. Genomic positions are based on GRCh37. Anti-*H. pylori* IgG seropositivity was defined as >3 U/mL and does not directly establish active infection. The $\lambda_{GC}$ values were 1.049 and 1.052 for panels a and b, respectively. $\lambda_{GC}$ was calculated from the median chi-square statistic using all reported *P* values in the corresponding source summary-statistics file.

Supplementary Figure 4

### a Discovery GWAS

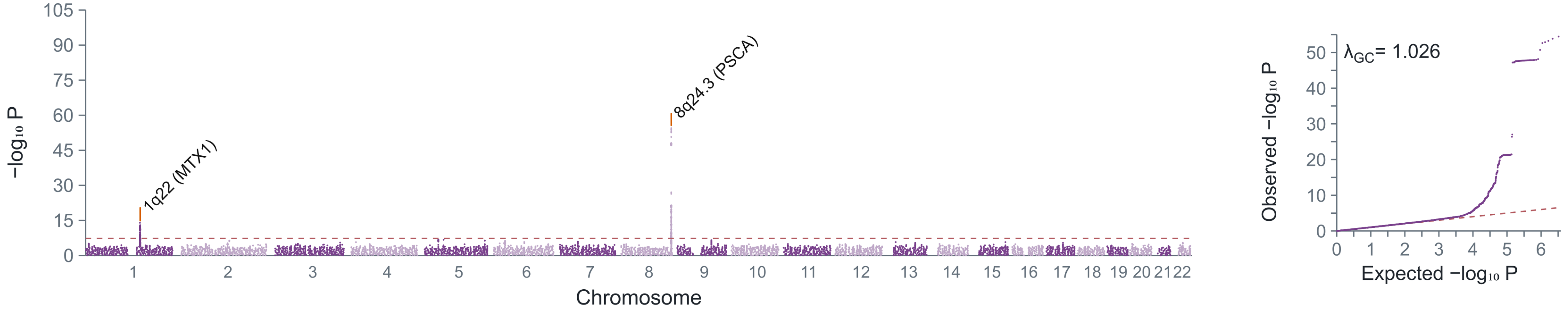


### b Replication GWAS

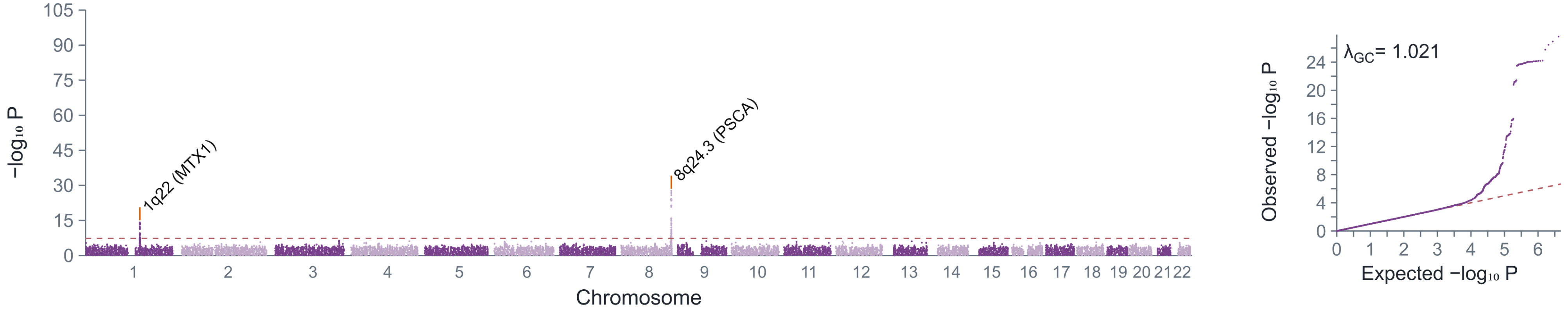

### Supplementary Figure 4. Manhattan and quantile-quantile plots for the discovery and replication anti-*H. pylori* IgG-seropositive SAG GWAS.

Panels show (a) discovery and (b) replication GWAS results. Manhattan plots show $-\log_{10}(P)$ values by genomic position; the red dashed line indicates $P = 5 \times 10^{-8}$. Selected genome-wide significant loci are annotated. Quantile-quantile plots compare observed and expected $-\log_{10}(P)$ values. Genomic positions are based on GRCh37. Anti-*H. pylori* IgG seropositivity was defined as >3 U/mL and does not directly establish active infection. The $\lambda_{GC}$ values were 1.026 and 1.021 for panels a and b, respectively. $\lambda_{GC}$ was calculated from the median chi-square statistic using all reported *P* values in the corresponding source summary-statistics file.

### a Discovery GWAS

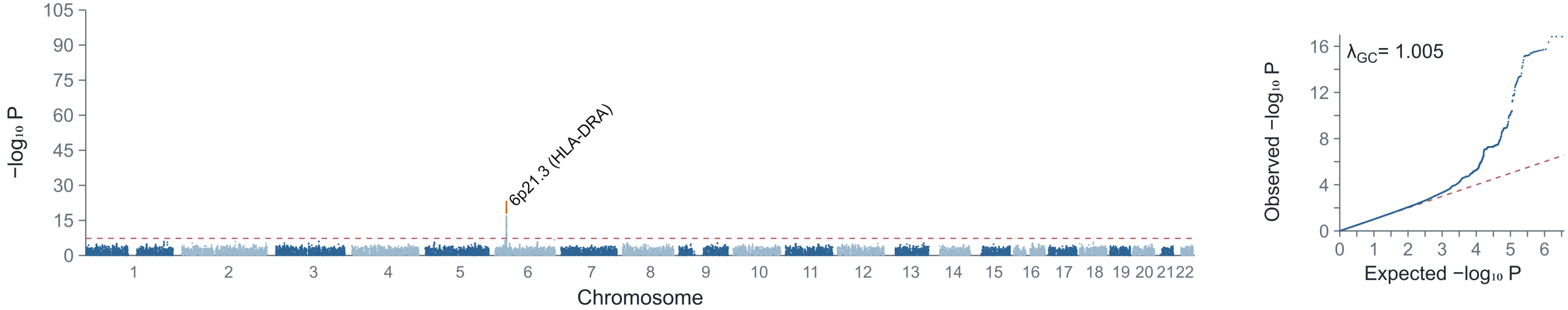


### b Replication GWAS

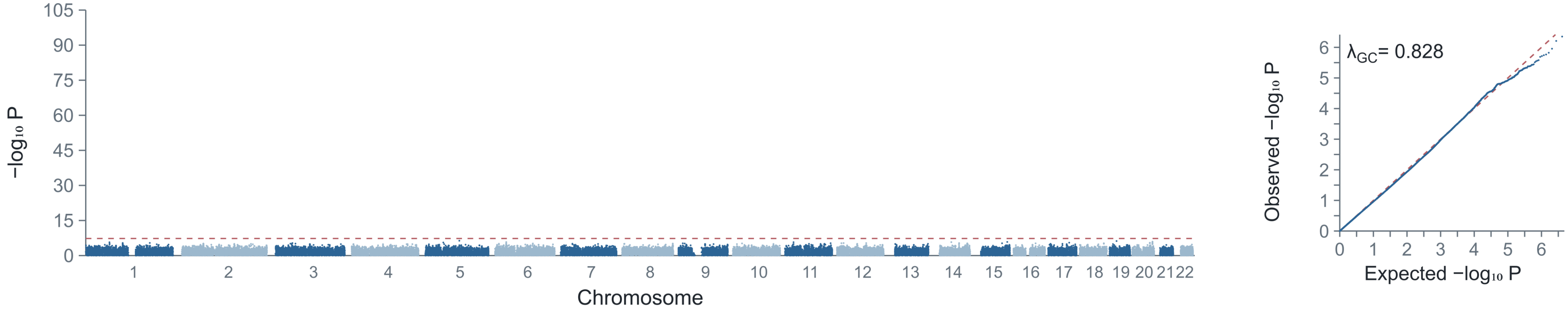

**Supplementary Figure 5. Manhattan and quantile-quantile plots for the discovery and replication anti-*H. pylori* IgG-seronegative AG GWAS.**

Panels show (a) discovery and (b) replication GWAS results. Manhattan plots show $-\log_{10}(P)$ values by genomic position; the red dashed line indicates $P = 5 \times 10^{-8}$. Selected genome-wide significant loci are annotated. Quantile-quantile plots compare observed and expected $-\log_{10}(P)$ values. Genomic positions are based on GRCh37. Anti-*H. pylori* IgG seronegativity was defined as <=3 U/mL and does not directly establish absence of current or previous infection. The $\lambda_{GC}$ values were 1.005 and 0.828 for panels a and b, respectively. $\lambda_{GC}$ was calculated from the median chi-square statistic using all reported *P* values in the corresponding source summary-statistics file.

Supplementary Figure 6

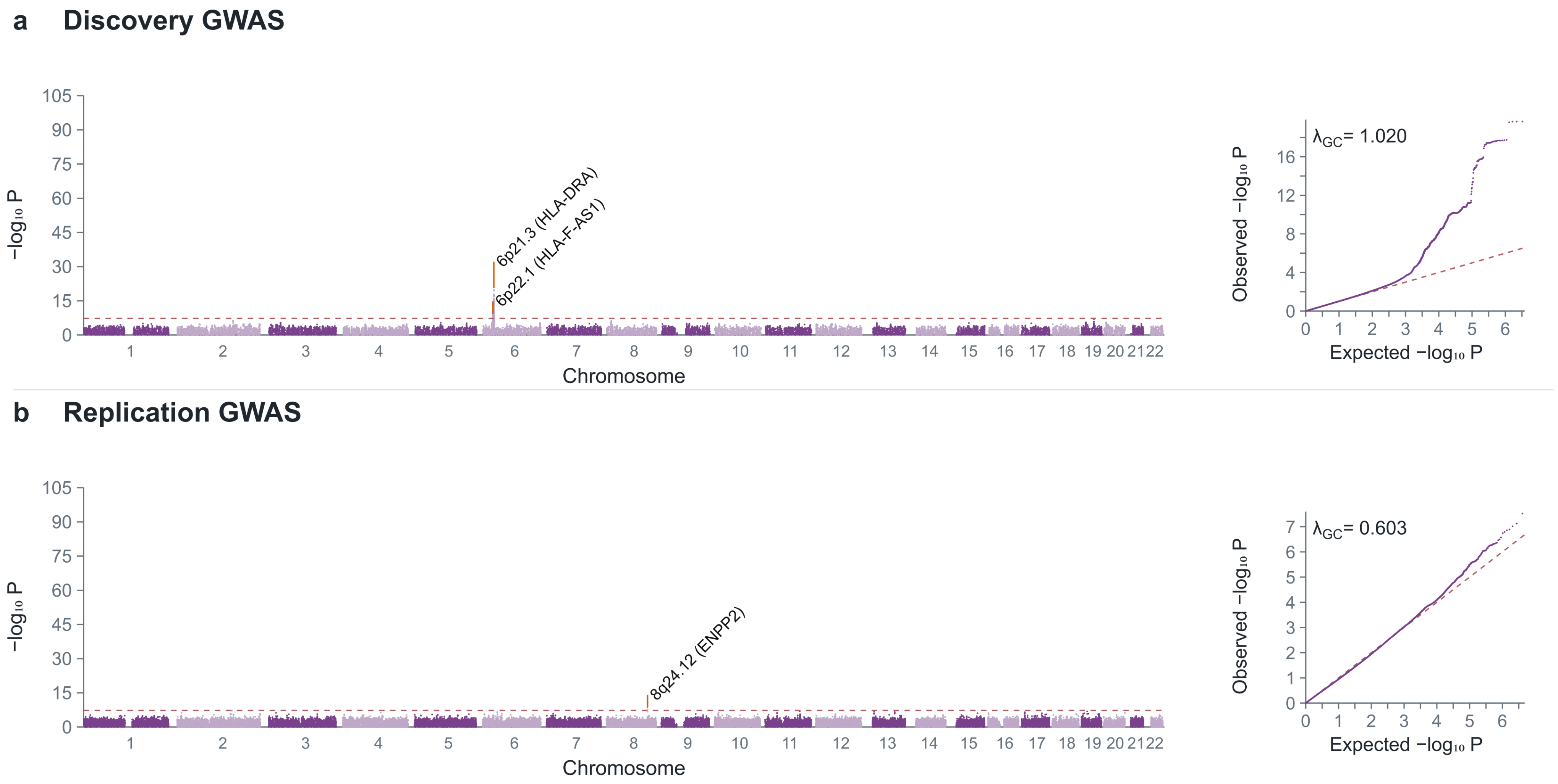

### Supplementary Figure 6. Manhattan and quantile-quantile plots for the discovery and replication anti-*H. pylori* IgG-seronegative SAG GWAS.

Panels show (a) discovery and (b) replication GWAS results. Manhattan plots show $-\log_{10}(P)$ values by genomic position; the red dashed line indicates $P = 5 \times 10^{-8}$. Selected genome-wide significant loci are annotated. Quantile-quantile plots compare observed and expected $-\log_{10}(P)$ values. Genomic positions are based on GRCh37. Anti-*H. pylori* IgG seronegativity was defined as <=3 U/mL and does not directly establish absence of current or previous infection. The $\lambda_{GC}$ values were 1.020 and 0.603 for panels a and b, respectively. $\lambda_{GC}$ was calculated from the median chi-square statistic using all reported *P* values in the corresponding source summary-statistics file.

Supplementary Figure 7

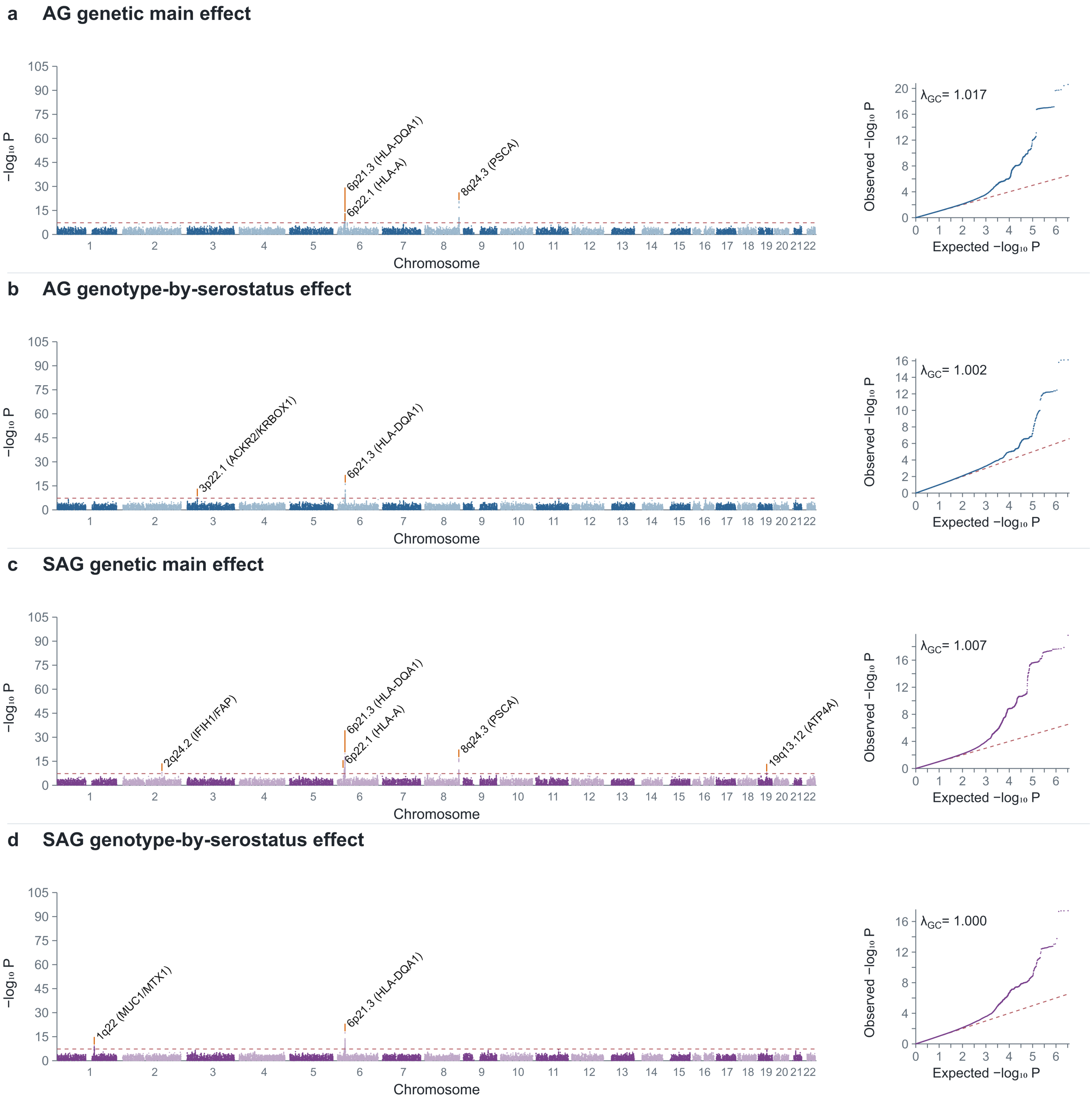

### Supplementary Figure 7. Genome-wide genotype-by-anti-*H. pylori* IgG serostatus interaction analyses for AG and SAG.

Genome-wide genotype-by-anti-*H. pylori* IgG serostatus interaction analyses were performed for AG and SAG. Panels show conditional genetic main effects and interaction effects for AG (a and b) and SAG (c and d). Logistic regression models were adjusted for age, sex, and principal components 1–10. Covariates, including serostatus, were variance-standardized in PLINK 2.0; conditional main effects are evaluated at mean serostatus. Seropositivity and seronegativity were defined as anti-*H. pylori* IgG >3 U/mL and ≤3 U/mL, respectively. The red dashed line indicates $P = 5 \times 10^{-8}$. The $\lambda_{GC}$ values were 1.017, 1.002, 1.007, and 1.000 for panels a–d, respectively, calculated from all reported *P* values in the corresponding source files. Positions are based on GRCh37.

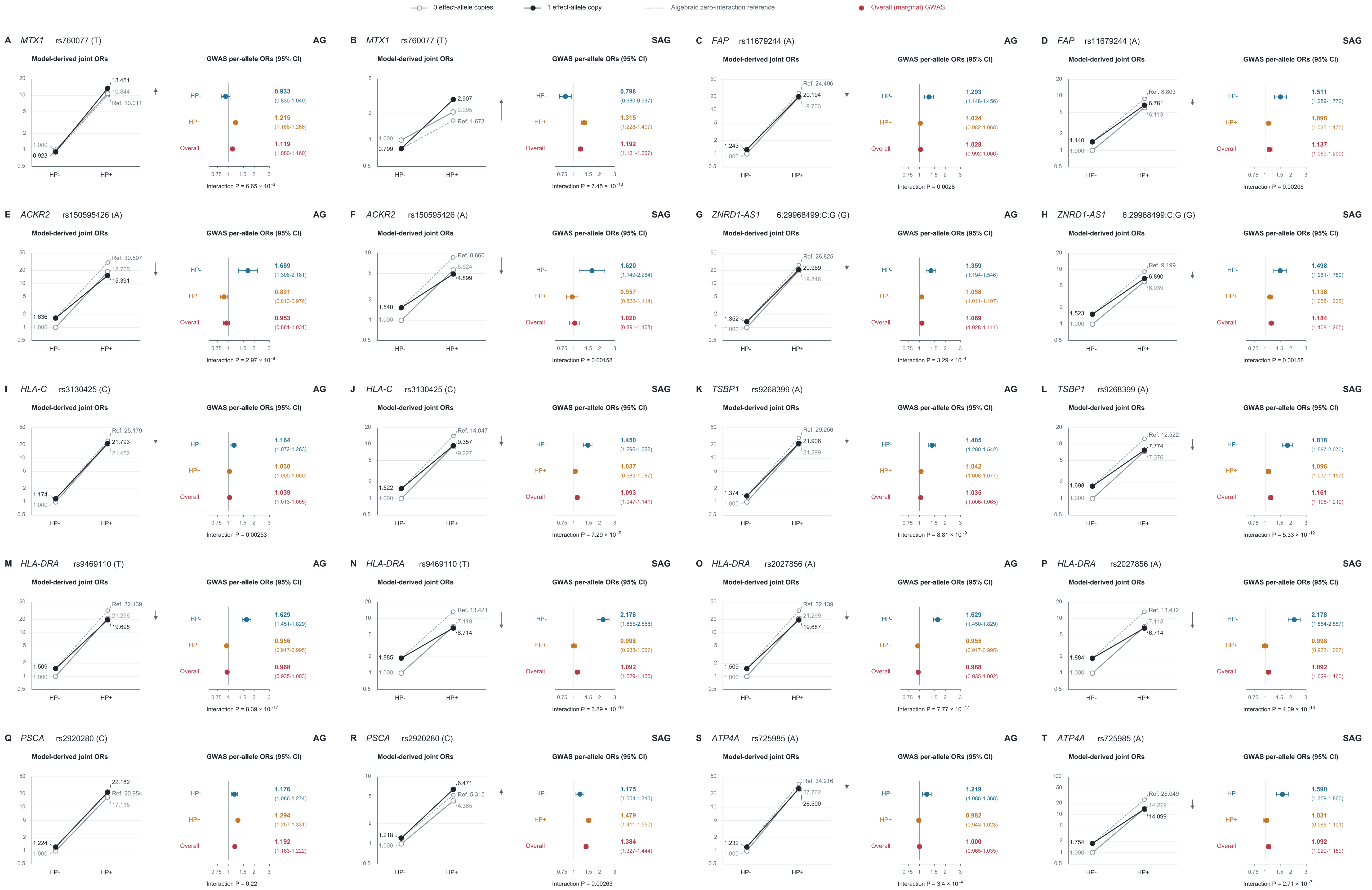

0 effect-allele copies
1 effect-allele copy
Algebraic zero-interaction reference
Overall (marginal) GWAS
A MTX1 rs760077 (T) AG
Model-derived joint ORs
13.451
10.844
Ref. 10.011
1.000
0.923
GWAS per-allele ORs (95% CI)
HP- 0.933 (0.830-1.049)
HP+ 1.215 (1.166-1.266)
Overall 1.119 (1.080-1.160)
Interaction P = 6.65 × 10 -6
B MTX1 rs760077 (T) SAG
Model-derived joint ORs
2.907
2.095
Ref. 1.673
1.000
0.799
GWAS per-allele ORs (95% CI)
HP- 0.798 (0.680-0.937)
HP+ 1.315 (1.228-1.407)
Overall 1.192 (1.121-1.267)
Interaction P = 7.45 × 10 -10
C FAP rs11679244 (A) AG
Model-derived joint ORs
Ref. 24.496
20.194
19.703
1.243
1.000
GWAS per-allele ORs (95% CI)
HP- 1.293 (1.148-1.456)
HP+ 1.024 (0.982-1.068)
Overall 1.028 (0.992-1.066)
Interaction P = 0.0028
D FAP rs11679244 (A) SAG
Model-derived joint ORs
Ref. 8.803
6.761
6.113
1.440
1.000
GWAS per-allele ORs (95% CI)
HP- 1.511 (1.289-1.772)
HP+ 1.098 (1.025-1.176)
Overall 1.137 (1.069-1.209)
Interaction P = 0.00206
E ACKR2 rs150595426 (A) AG
Model-derived joint ORs
Ref. 30.597
18.705
15.391
1.636
1.000
GWAS per-allele ORs (95% CI)
HP- 1.689 (1.308-2.181)
HP+ 0.891 (0.813-0.976)
Overall 0.953 (0.881-1.031)
Interaction P = 2.97 × 10 -8
F ACKR2 rs150595426 (A) SAG
Model-derived joint ORs
Ref. 8.660
5.624
4.899
1.540
1.000
GWAS per-allele ORs (95% CI)
HP- 1.620 (1.149-2.284)
HP+ 0.957 (0.822-1.114)
Overall 1.020 (0.891-1.168)
Interaction P = 0.00158
G ZNRD1-AS1 6:29968499:C:G (G) AG
Model-derived joint ORs
Ref. 26.825
20.969
19.846
1.352
1.000
GWAS per-allele ORs (95% CI)
HP- 1.359 (1.194-1.546)
HP+ 1.058 (1.011-1.107)
Overall 1.069 (1.028-1.111)
Interaction P = 3.29 × 10 -4
H ZNRD1-AS1 6:29968499:C:G (G) SAG
Model-derived joint ORs
Ref. 9.199
6.890
6.039
1.523
1.000
GWAS per-allele ORs (95% CI)
HP- 1.498 (1.261-1.780)
HP+ 1.138 (1.058-1.225)
Overall 1.184 (1.108-1.265)
Interaction P = 0.00158
I HLA-C rs3130425 (C) AG
Model-derived joint ORs
Ref. 25.179
21.793
21.452
1.174
1.000
GWAS per-allele ORs (95% CI)
HP- 1.164 (1.072-1.263)
HP+ 1.030 (1.000-1.060)
Overall 1.039 (1.013-1.065)
Interaction P = 0.00253
J HLA-C rs3130425 (C) SAG
Model-derived joint ORs
Ref. 14.047
9.357
9.227
1.522
1.000
GWAS per-allele ORs (95% CI)
HP- 1.450 (1.296-1.622)
HP+ 1.037 (0.989-1.087)
Overall 1.093 (1.047-1.141)
Interaction P = 7.29 × 10 -9
K TSBP1 rs9268399 (A) AG
Model-derived joint ORs
Ref. 29.256
21.906
21.299
1.374
1.000
GWAS per-allele ORs (95% CI)
HP- 1.405 (1.280-1.542)
HP+ 1.042 (1.008-1.077)
Overall 1.035 (1.006-1.065)
Interaction P = 8.81 × 10 -9
L TSBP1 rs9268399 (A) SAG
Model-derived joint ORs
Ref. 12.522
7.774
7.376
1.698
1.000
GWAS per-allele ORs (95% CI)
HP- 1.818 (1.597-2.070)
HP+ 1.096 (1.037-1.157)
Overall 1.161 (1.105-1.219)
Interaction P = 5.33 × 10 -12
M HLA-DRA rs9469110 (T) AG
Model-derived joint ORs
Ref. 32.139
21.296
19.695
1.509
1.000
GWAS per-allele ORs (95% CI)
HP- 1.629 (1.451-1.829)
HP+ 0.956 (0.917-0.995)
Overall 0.968 (0.935-1.003)
Interaction P = 8.39 × 10 -17
N HLA-DRA rs9469110 (T) SAG
Model-derived joint ORs
Ref. 13.421
7.119
6.714
1.885
1.000
GWAS per-allele ORs (95% CI)
HP- 2.178 (1.855-2.558)
HP+ 0.998 (0.933-1.067)
Overall 1.092 (1.029-1.160)
Interaction P = 3.89 × 10 -18
O HLA-DRA rs2027856 (A) AG
Model-derived joint ORs
Ref. 32.139
21.299
19.687
1.509
1.000
GWAS per-allele ORs (95% CI)
HP- 1.629 (1.450-1.829)
HP+ 0.955 (0.917-0.995)
Overall 0.968 (0.935-1.002)
Interaction P = 7.77 × 10 -17
P HLA-DRA rs2027856 (A) SAG
Model-derived joint ORs
Ref. 13.412
7.118
6.714
1.884
1.000
GWAS per-allele ORs (95% CI)
HP- 2.178 (1.854-2.557)
HP+ 0.998 (0.933-1.067)
Overall 1.092 (1.029-1.160)
Interaction P = 4.09 × 10 -18
Q PSCA rs2920280 (C) AG
Model-derived joint ORs
22.182
Ref. 20.954
17.115
1.224
1.000
GWAS per-allele ORs (95% CI)
HP- 1.176 (1.086-1.274)
HP+ 1.294 (1.257-1.331)
Overall 1.192 (1.163-1.222)
Interaction P = 0.22
R PSCA rs2920280 (C) SAG
Model-derived joint ORs
6.471
Ref. 5.315
4.365
1.218
1.000
GWAS per-allele ORs (95% CI)
HP- 1.175 (1.054-1.310)
HP+ 1.479 (1.411-1.550)
Overall 1.384 (1.327-1.444)
Interaction P = 0.00263
S ATP4A rs725985 (A) AG
Model-derived joint ORs
Ref. 34.216
27.762
26.500
1.232
1.000
GWAS per-allele ORs (95% CI)
HP- 1.219 (1.086-1.368)
HP+ 0.982 (0.943-1.023)
Overall 1.000 (0.965-1.035)
Interaction P = 3.4 × 10 -4
T ATP4A rs725985 (A) SAG
Model-derived joint ORs
Ref. 25.049
14.279
14.099
1.754
1.000
GWAS per-allele ORs (95% CI)
HP- 1.590 (1.359-1.860)
HP+ 1.031 (0.965-1.101)
Overall 1.092 (1.029-1.159)
Interaction P = 2.71 × 10 -7
HP-
HP+

### Supplementary Figure 8. Genotype-by-serostatus associations in AG and SAG.

Genotype-by-anti-*H. pylori* IgG serostatus associations at ten variants in AG and SAG. Joint odds ratios (ORs) are approximate model-derived estimates relative to seronegative participants with 0 effect-allele copies (Supplementary Table 10). Environmental coefficients were allele-aligned, converted to binary-serostatus contrasts using dataset-specific standardization, and pooled by inverse variance. Dashed lines show the algebraic zero-interaction reference, $OR_{11}(ref) = OR_{01} \times OR_{10}/OR_{00}$, without refitting. Right, separate TMM marginal and serostatus-stratified GWAS per-allele ORs with 95% CIs (Table 2). Marginal estimates are red. Interaction P values refer to the standardized-serostatus G×E term. Arrows indicate contrasts, not significance. Coefficient covariances were unavailable, so joint ORs have no CIs. Both OR axes are logarithmic, with panel-specific joint-OR ranges. Ref., reference OR; HP−/HP+, anti-*H. pylori* IgG seronegative/seropositive; AG, atrophic gastritis; SAG, severe atrophic gastritis.

### BBJ-excluded overall AG

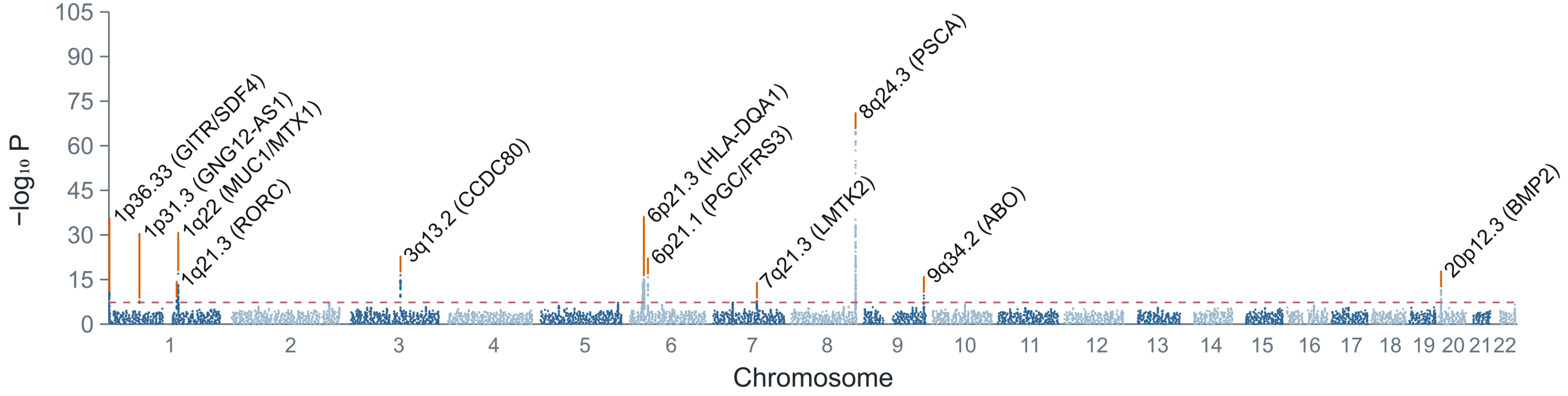


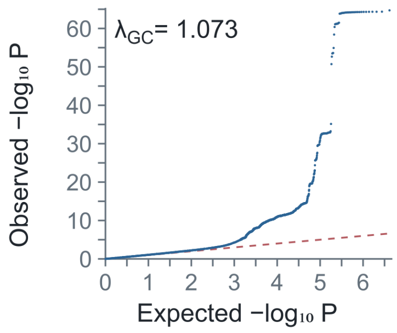

### Supplementary Figure 9. Manhattan and quantile-quantile plots for the BBJ-excluded overall AG meta-analysis.

The BBJ-excluded Japanese overall AG meta-analysis was used in shared-genetic analyses with the BBJ gastric cancer GWAS to avoid sample overlap. The red dashed line indicates $P = 5 \times 10^{-8}$. Genomic positions are based on GRCh37. The $\lambda_{GC}$ value was 1.073. $\lambda_{GC}$ was calculated from the median chi-square statistic using all reported $P$ values in the corresponding source summary-statistics file.

**a** **MTAG-adjusted AG summary statistics**

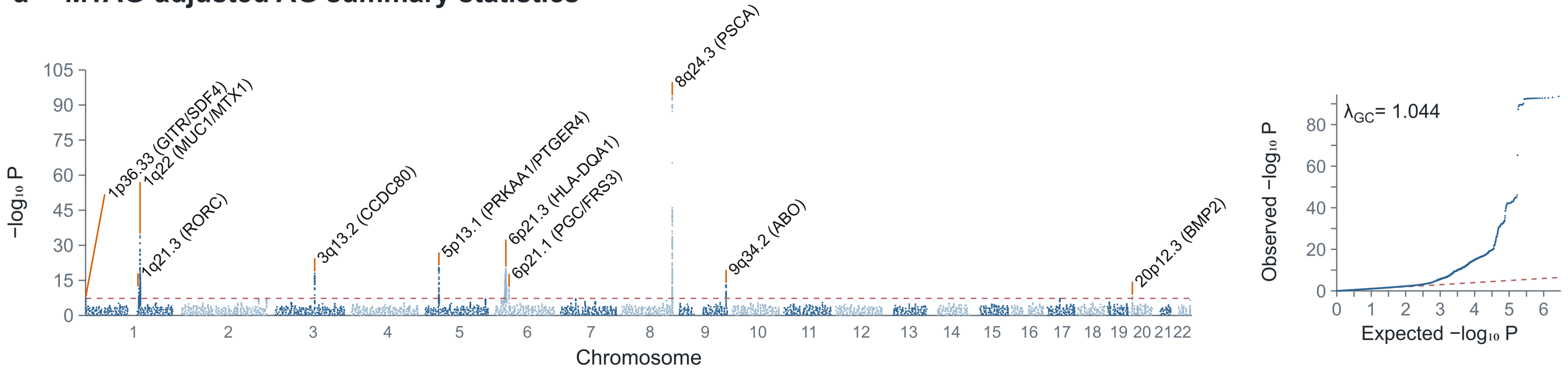


**b** **MTAG-adjusted GC summary statistics**

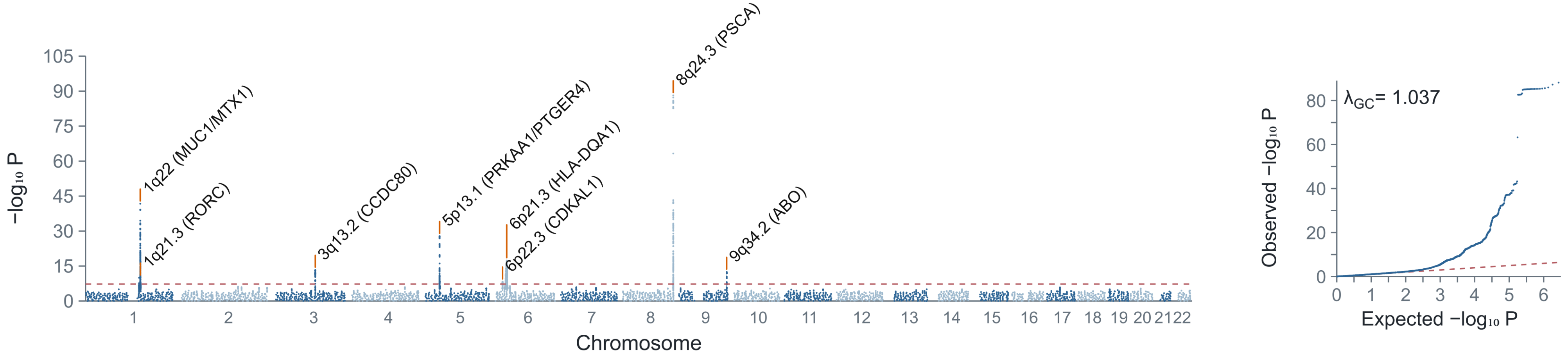

### Supplementary Figure 10. Manhattan and quantile-quantile plots for MTAG-adjusted AG and gastric cancer summary statistics.

Panels show MTAG-adjusted summary statistics for (a) AG and (b) gastric cancer. MTAG jointly analyzed BBJ-excluded Japanese AG summary statistics and BBJ gastric cancer summary statistics. The red dashed line indicates $P = 5 \times 10^{-8}$. Selected genome-wide significant loci are annotated. Genomic positions are based on GRCh37. The $\lambda_{GC}$ values were 1.044 and 1.037 for panels a and b, respectively. $\lambda_{GC}$ was calculated from the median chi-square statistic using all reported $P$ values in the corresponding source summary-statistics file. The corresponding genome-wide significant loci are listed in Supplementary Table 15.

### a Genomic SEM AG-GC shared component

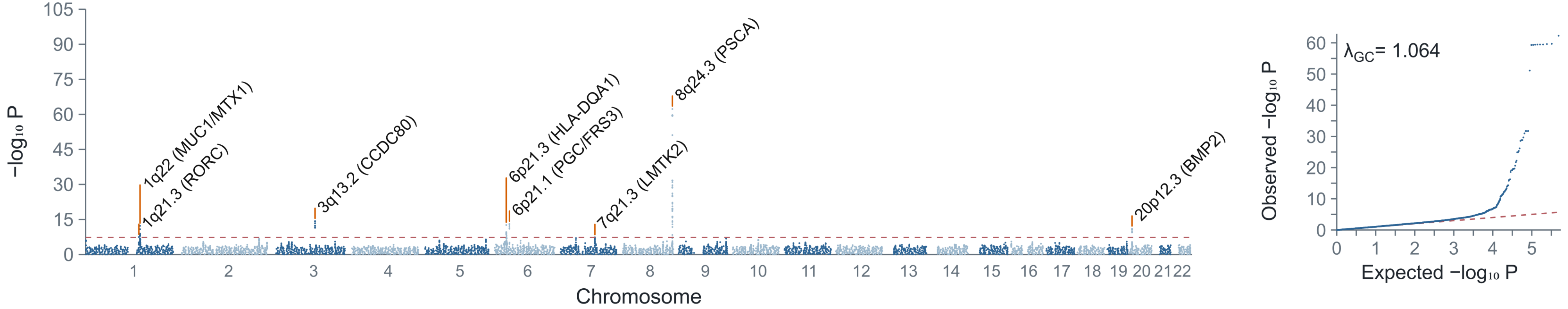


### b Genomic SEM GC-specific component

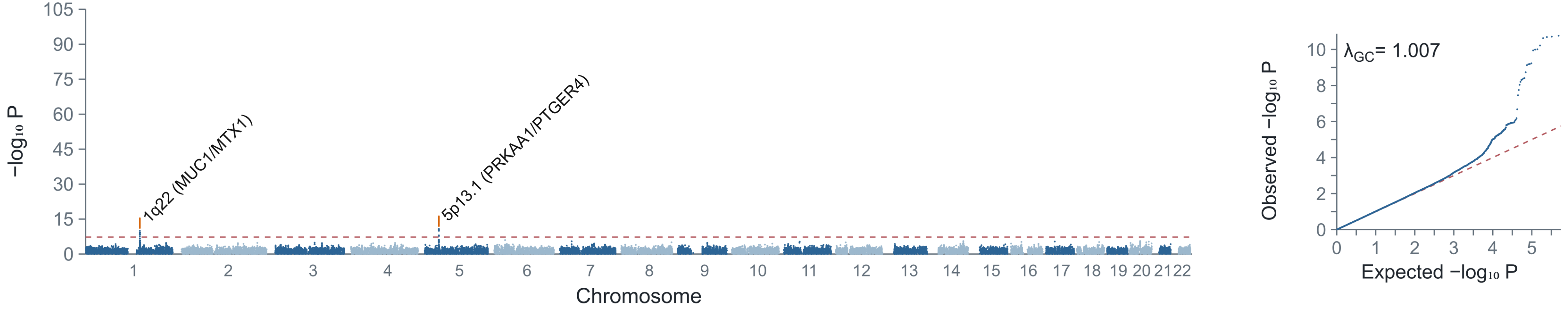

### Supplementary Figure 11. Manhattan and quantile-quantile plots for Genomic SEM GWAS-by-subtraction components.

Panels show (a) the AG-anchored shared component and (b) the GC-specific residual component from the Genomic SEM GWAS-by-subtraction model shown in Figure 3. The shared component represents SNP associations with the model-defined factor anchored to AG. The GC-specific component represents residual gastric cancer associations after accounting for that factor and does not imply complete absence of AG-related effects. The red dashed line indicates $P = 5 \times 10^{-8}$. The $\lambda_{GC}$ values were 1.064 and 1.007 for panels a and b, respectively. $\lambda_{GC}$ was calculated from the median chi-square statistic using all reported $P$ values in the corresponding source summary-statistics file. Genome-wide significant loci are listed in Supplementary Table 17.

**a** **Cross-ancestry SAG**

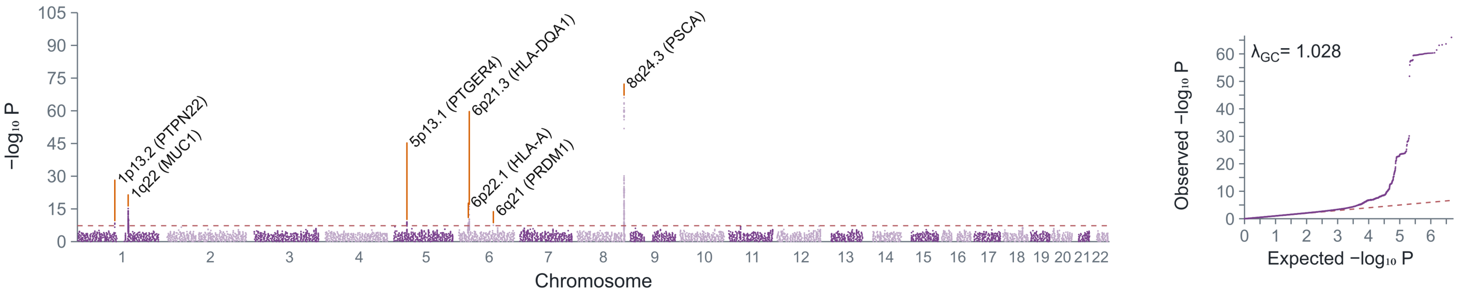


**b** **Cross-ancestry anti-H. pylori IgG-seropositive AG**

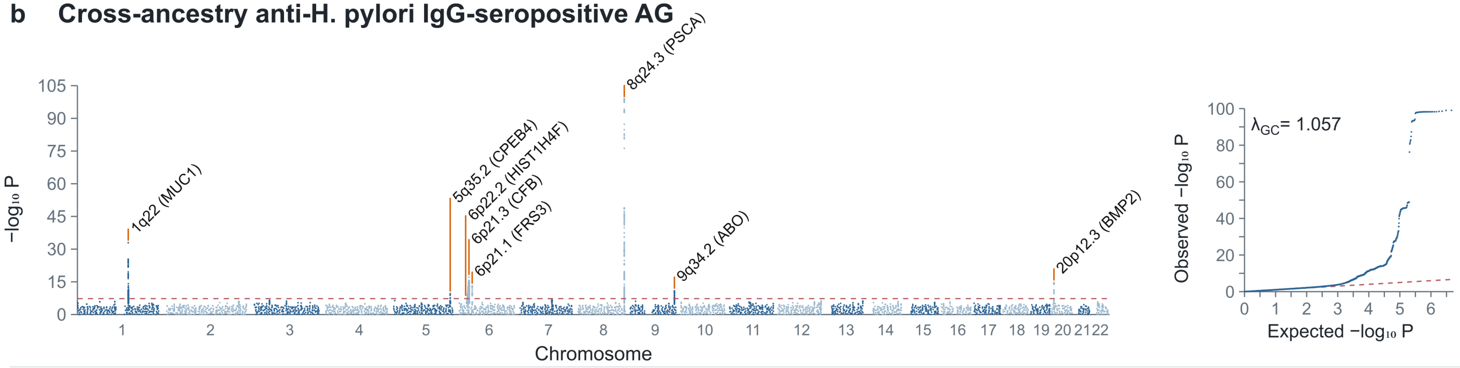


**c** **Cross-ancestry anti-H. pylori IgG-seropositive SAG**

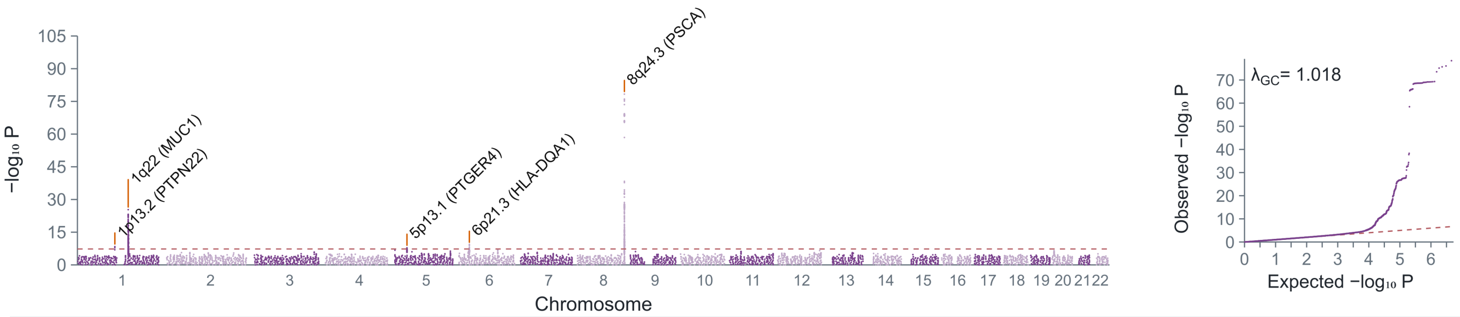


**d** **Cross-ancestry anti-H. pylori IgG-seronegative AG**

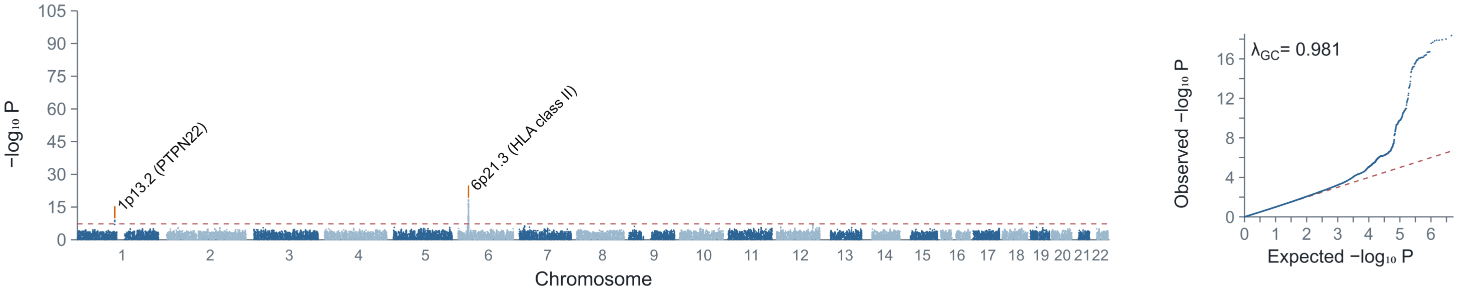

**Supplementary Figure 12. Supportive cross-ancestry meta-analyses for the remaining four AG/SAG phenotypes.**

Panels show MR-MEGA results for (a) overall SAG, (b) anti-*H. pylori* IgG-seropositive AG, (c) anti-*H. pylori* IgG-seropositive SAG, and (d) anti-*H. pylori* IgG-seronegative AG. Results for overall AG and anti-*H. pylori* IgG-seronegative SAG are shown in Figure 4. Japanese AG/SAG summary statistics were integrated with external chronic gastritis GWAS summary statistics from FinnGen, UK Biobank, and MVP. Because the external phenotype was chronic gastritis rather than serologically defined AG/SAG, these analyses were interpreted as supportive evidence. Manhattan plots show $-\log_{10}(P)$ values by genomic position; the red dashed line indicates $P = 5 \times 10^{-8}$. Selected genome-wide significant loci are annotated. Quantile-quantile plots compare observed and expected $-\log_{10}(P)$ values. Genomic positions are based on GRCh37. Anti-*H. pylori* IgG seronegativity was defined as <=3 U/mL and does not directly establish absence of current or previous infection. The $\lambda_{GC}$ values were 1.028, 1.057, 1.018, and 0.981 for panels a-d, respectively. $\lambda_{GC}$ was calculated from the median chi-square statistic using all reported $P$ values in the corresponding source summary-statistics file.
