## Supplementary Tables 1-18 for "Genome-wide association meta-analysis of atrophic gastritis reveals anti–*H. pylori* IgG serostatus-dependent associations and genetic overlap with gastric cancer"

### Supplementary Table 1. Cohort- and dataset-specific sample sizes and characteristics for the AG/SAG GWAS.

| Cohort | Dataset | Study | | Sample/data collection | | AG | | SAG | | HP-positive | | | | HP-negative | | | |
|---|---|---|---|---|---|---|---|---|---|---|---|---|---|---|---|---|---|
| | | | | | | | | | | AG | | SAG | | AG | | SAG | |
| | | D/R | No. of participants | From | To | Case | Control | Case | Control | Case | Control | Case | Control | Case | Control | Case | Control |
| TMM CommCohort | 1 | D | 47831 | 2013 | 2016 | 9956 | 37875 | 3013 | 44818 | 9283 | 14831 | 2620 | 21494 | 668 | 23032 | 391 | 23309 |
| TMM CommCohort | 2 | D | 9136 | 2013 | 2016 | 2510 | 6626 | 644 | 8492 | 2330 | 2833 | 572 | 4591 | 162 | 3778 | 68 | 3872 |
| TMM CommCohort | 3 | D | 11442 | 2013 | 2016 | 1972 | 9470 | 598 | 10844 | 1793 | 2949 | 489 | 4253 | 176 | 6512 | 107 | 6581 |
| TMM CommCohort | 4 | D | 6665 | 2013 | 2016 | 1206 | 5459 | 262 | 6403 | 1096 | 1988 | 217 | 2867 | 110 | 3471 | 45 | 3536 |
| TMM BirThree Cohort | 5 | R | 16552 | 2013 | 2017 | 836 | 15716 | 204 | 16348 | 765 | 3588 | 172 | 4181 | 71 | 12122 | 32 | 12161 |
| TMM BirThree Cohort | 6 | R | 14285 | 2013 | 2017 | 569 | 13716 | 120 | 14165 | 502 | 3349 | 94 | 3757 | 67 | 10362 | 26 | 10403 |
| Yamagata Study | 7 | R | 1591 | 2004 | 2006 | 746 | 845 | 278 | 1313 | 727 | 610 | 267 | 1070 | 19 | 235 | 11 | 243 |
| Yamagata Study | 8 | R | 2329 | 2009 | 2015 | 1049 | 1280 | 345 | 1984 | 997 | 656 | 323 | 1330 | 52 | 624 | 22 | 654 |
| Yamagata Study | 9 | R | 1214 | 2009 | 2015 | 448 | 766 | 124 | 1090 | 429 | 392 | 120 | 701 | 19 | 374 | 4 | 389 |
| JPHC | 10 | R | 5339 | 1993 | 1994 | 2206 | 3133 | 594 | 4745 | 2141 | 2048 | 555 | 3634 | 65 | 1085 | 39 | 1111 |
| J-MICC | 11 | R | 3472 | 2005 | 2012 | 600 | 2872 | 169 | 3303 | 546 | 1103 | 151 | 1498 | 53 | 1765 | 17 | 1801 |
| HERPACC | 12 | R | 620 | 2001 | 2005 | 176 | 444 | 53 | 567 | 160 | 120 | 44 | 236 | 16 | 324 | - | - |
| HERPACC | 13 | R | 2758 | 2001 | 2013 | 688 | 2070 | 263 | 2495 | 645 | 897 | 238 | 1304 | 43 | 1173 | 25 | 1191 |
| HERPACC | 14 | R | 1804 | 2001 | 2013 | 468 | 1336 | 153 | 1651 | 448 | 686 | 139 | 995 | 20 | 650 | 14 | 656 |
| BBJ | 15 | R | 69236 | 2003 | 2013 | 972 | 68264 | | | | | | | | | | |
| BBJ | 16 | R | 3277 | 2003 | 2013 | 24 | 3253 | | | | | | | | | | |
| BBJ | 17 | R | 17196 | 2013 | 2017 | 766 | 16430 | | | | | | | | | | |

Dataset numbers identify genotyping-array-specific analysis datasets. D and R denote discovery and replication datasets. AG and SAG were defined using serum pepsinogen criteria except in BBJ, where medically diagnosed AG was used; BBJ was included only in overall AG. Anti-*H. pylori* IgG seropositivity and seronegativity were defined as >3 U/mL and <=3 U/mL, respectively. AG, atrophic gastritis; BBJ, BioBank Japan; D, discovery; HERPACC, Hospital-based Epidemiologic Research Program at Aichi Cancer Center; J-MICC, Japan Multi-Institutional Collaborative Cohort Study; JPHC, Japan Public Health Center-based Prospective Study; R, replication; SAG, severe atrophic gastritis; TMM, Tohoku Medical Megabank.

Both BBJ1 and BBJ2 collected DNA and clinical information, and BBJ1 also collected serum.

**Supplementary Table 2. Genotyping, imputation, quality-control, and GWAS analysis settings by dataset.**

| Cohort | Dataset | Genotyping | | | Preimputation QC | | | | Imputation | | Post-imputation QC | | GWAS |
|---|---|---|---|---|---|---|---|---|---|---|---|---|---|
| | | Genotyping array | Calling algorithm | Sample call rate cutoff | other criteria | SNP call rate cutoff | SNP MAF cutoff | SNP HWE P cutoff | Reference | Software | SNP INFO or Rsq cutoff | SNP MAF or MAC cutoff | Software |
| TMM CommCohort | 1 | Japonica Array version 2 | Affymetrix Power Tools | 0.97 | sex mismatches, gastric cancer history | 0.99 | 0.01 | $1.00 \times 10^{-5}$ | 3.5KJPNv2 × 1000 Genomes Project phase 3 | SHAPEIT2 & IMPUTE4 | INFO >=0.4 | MAF >=0.01 | GCTA |
| TMM CommCohort | 2 | Japonica Array NEO | Affymetrix Power Tools | 0.97 | sex mismatches, gastric cancer history | 0.99 | 0.01 | $1.00 \times 10^{-5}$ | 3.5KJPNv2 × 1000 Genomes Project phase 3 | SHAPEIT2 & IMPUTE4 | INFO >=0.4 | MAF >=0.01 | GCTA |
| TMM CommCohort | 3 | Japonica Array version 2 | Affymetrix Power Tools | 0.97 | sex mismatches, gastric cancer history | 0.99 | 0.01 | $1.00 \times 10^{-5}$ | 3.5KJPNv2 × 1000 Genomes Project phase 3 | SHAPEIT2 & IMPUTE4 | INFO >=0.4 | MAF >=0.01 | GCTA |
| TMM CommCohort | 4 | Japonica Array version 2 | Affymetrix Power Tools | 0.97 | sex mismatches, gastric cancer history | 0.99 | 0.01 | $1.00 \times 10^{-5}$ | 3.5KJPNv2 × 1000 Genomes Project phase 3 | SHAPEIT2 & IMPUTE4 | INFO >=0.4 | MAF >=0.01 | GCTA |
| TMM BirThree Cohort | 5 | Japonica Array version 2 | Affymetrix Power Tools | 0.97 | sex mismatches, gastric cancer history | 0.99 | 0.01 | $1.00 \times 10^{-5}$ | 3.5KJPNv2 × 1000 Genomes Project phase 3 | SHAPEIT2 & IMPUTE4 | INFO >=0.4 | MAF >=0.01 | GCTA |
| TMM BirThree Cohort | 6 | Japonica Array NEO | Affymetrix Power Tools | 0.97 | sex mismatches, gastric cancer history | 0.99 | 0.01 | $1.00 \times 10^{-5}$ | 3.5KJPNv2 × 1000 Genomes Project phase 3 | SHAPEIT2 & IMPUTE4 | INFO >=0.4 | MAF >=0.01 | GCTA |
| Yamagata Study | 7 | Human660W-Quad | GenomeStudio | 0.97 | sex mismatches, gastric cancer history | 0.99 | 0.01 | $1.00 \times 10^{-5}$ | 1000 Genomes Project phase 3 | SHAPEIT2 & IMPUTE4 | INFO >=0.4 | MAF >=0.01 | GCTA |
| Yamagata Study | 8 | Japonica Array NEO | Affymetrix Power Tools | 0.97 | sex mismatches, gastric cancer history | 0.99 | 0.01 | $1.00 \times 10^{-5}$ | 3.5KJPNv2 × 1000 Genomes Project phase 3 | SHAPEIT2 & IMPUTE4 | INFO >=0.4 | MAF >=0.01 | GCTA |
| Yamagata Study | 9 | HumanOmniExpressExome | GenomeStudio | 0.97 | sex mismatches, gastric cancer history | 0.99 | 0.01 | $1.00 \times 10^{-5}$ | 1000 Genomes Project phase 3 | SHAPEIT2 & IMPUTE4 | INFO >=0.4 | MAF >=0.01 | GCTA |
| JPHC | 10 | HumanOmniExpressExome BeadChip | GenomeStudio | 0.98 | sex mismatches, gastric cancer history, related samples (IBD 0.1875) | 0.99 | 0.01 | $1.00 \times 10^{-6}$ | 1000 Genomes Project phase 3 | SHAPEIT2 & IMPUTE2 | NA | MAF >=0.01 | SNPTEST |
| J-MICC | 11 | HumanOmniExpressExome | GenomeStudio | 0.98 | sex mismatches, gastric cancer history, related samples (IBD 0.1875) | 0.98 | 0.01 | $1.00 \times 10^{-6}$ | 1000 Genomes Project phase 3 | SHAPEIT2 & IMPUTE2 | INFO >=0.4 | MAF >=0.01 | REGENIE |
| HERPACC | 12 | Human610-Quad | GenomeStudio | 0.98 | sex mismatches, gastric cancer history, related samples (IBD 0.1875) | 0.98 | 0.01 | $1.00 \times 10^{-6}$ | 1000 Genomes Project phase 3 | SHAPEIT2 & minimac3 | INFO >=0.4 | MAF >=0.01 | REGENIE |
| HERPACC | 13 | Infinium Asian Screening Array | GenomeStudio | 0.95 | sex mismatches, gastric cancer history, related samples (IBD 0.1875) | 0.98 | 0.01 | $1.00 \times 10^{-6}$ | 1000 Genomes Project phase 3 | SHAPEIT2 & minimac3 | INFO >=0.4 | MAF >=0.01 | REGENIE |
| HERPACC | 14 | HumanCoreExome | GenomeStudio | 0.98 | sex mismatches, gastric cancer history, related samples (IBD 0.1875) | 0.98 | 0.01 | $1.00 \times 10^{-6}$ | 1000 Genomes Project phase 3 | SHAPEIT2 & minimac3 | INFO >=0.4 | MAF >=0.01 | REGENIE |
| BBJ | 15 | HumanOmniExpressExome BeadChip or a combination of HumanOmniExpress and HumanExome BeadChip | GenomeStudio | 0.98 | sex mismatches, gastric cancer history, non-EAS individuals, related samples (PI_HAT > 0.175) | 0.99 | NA | $1.00 \times 10^{-6}$ | 1000 Genomes Project phase 3 | Eagle2 & Minimac3 | Rsq >=0.3 | MAC >=5 | REGENIE |
| BBJ | 16 | Infinium Asian Screening Array | GenomeStudio | 0.98 | sex mismatches, gastric cancer history, non-EAS individuals | 0.99 | NA | $1.00 \times 10^{-6}$ | 1000 Genomes Project phase 3 | Eagle2 & Minimac4 | Rsq >=0.3 | MAC >=5 | REGENIE |
| BBJ | 17 | Infinium Asian Screening Array | GenomeStudio | 0.98 | sex mismatches, gastric cancer history, non-EAS individuals | 0.99 | NA | $1.00 \times 10^{-6}$ | 1000 Genomes Project phase 3 | Eagle2 & Minimac4 | Rsq >=0.3 | MAC >=5 | REGENIE |

QC was performed separately for each cohort and genotyping-array dataset before and after imputation using study-specific criteria. Samples or variants below the stated call-rate, MAF, or imputation-quality threshold, and variants below the stated HWE P-value threshold, were excluded as applicable. The TMM cross-imputed reference panel combined 3.5KJPNv2 and 1000 Genomes Project Phase 3 data. Dataset-specific differences reflect the applicable study protocol and should not be interpreted as a single universal threshold. BBJ, BioBank Japan; EAS, East Asian; HERPACC, Hospital-based Epidemiologic Research Program at Aichi Cancer Center; HWE, Hardy-Weinberg equilibrium; IBD, identity by descent; INFO, imputation information score; J-MICC, Japan Multi-Institutional Collaborative Cohort Study; JPHC, Japan Public Health Center-based Prospective Study; MAC, minor allele count; MAF, minor allele frequency; QC, quality control; Rsq, imputation quality metric; TMM, Tohoku Medical Megabank.

BBJ variants with heterozygote count < 5 were excluded in datasets 15–17. In dataset 15, variants with concordance < 99.5% or NDR ≥ 0.5% versus WGS (n = 939) were also excluded.

### Supplementary Table 3. Lead SNPs from combined, discovery, and replication AG GWAS

| Analysis | Locus | Lead SNP | Position (GRCh37) | Effect allele | EAF | Beta | SE | OR | 95% CI | P-value | Nearest Gene | Annotation |
|---|---|---|---|---|---|---|---|---|---|---|---|---|
| Combined meta-GWAS | 1p36.33 | rs58980341 | chr1:1135107 | T | 0.18 | 0.096 | 0.014 | 1.101 | 1.071-1.131 | $3.35 \times 10^{-11}$ | TTLL10 | Intergenic |
| | 1p31.3 | rs75563559 | chr1:68422111 | T | 0.016 | 0.238 | 0.044 | 1.269 | 1.164-1.383 | $6.62 \times 10^{-8}$ | GNG12-AS1 | Intron |
| | 1q22 | rs760077 | chr1:155178782 | T | 0.855 | 0.126 | 0.015 | 1.134 | 1.101-1.168 | $1.45 \times 10^{-16}$ | MTX1 | Missense |
| | 3q13.2 | rs72943992 | chr3:112403577 | C | 0.63 | 0.097 | 0.011 | 1.102 | 1.078-1.126 | $8.19 \times 10^{-18}$ | CCDC80 | Intergenic |
| | 3q26.2 | rs28626343 | chr3:169526272 | C | 0.653 | 0.061 | 0.011 | 1.063 | 1.04-1.086 | $4.59 \times 10^{-8}$ | LRRC34 | Intron |
| | 6p22.1 | rs9260603 | chr6:29922754 | C | 0.592 | 0.081 | 0.011 | 1.084 | 1.061-1.108 | $9.20 \times 10^{-14}$ | HLA-A | Intergenic |
| | 6p21.3 | rs1130153 | chr6:32610947 | A | 0.94 | 0.202 | 0.025 | 1.224 | 1.165-1.285 | $2.69 \times 10^{-16}$ | HLA-DQA1 | Intron |
| | 6p21.1 | rs62396739 | chr6:41730412 | A | 0.266 | 0.098 | 0.012 | 1.103 | 1.077-1.129 | $4.23 \times 10^{-16}$ | FRS3 | Intergenic |
| | 7q21.3 | rs58880085 | chr7:97836465 | C | 0.278 | 0.068 | 0.012 | 1.07 | 1.045-1.096 | $3.05 \times 10^{-8}$ | LMTK2 | 3 Prime UTR |
| | 8q24.3 | rs2920279 | chr8:143762135 | C | 0.643 | 0.184 | 0.011 | 1.202 | 1.176-1.228 | $3.32 \times 10^{-61}$ | PSCA | Intron |
| | 9q34.2 | rs9411372 | chr9:136134068 | A | 0.216 | 0.077 | 0.013 | 1.08 | 1.053-1.108 | $8.19 \times 10^{-9}$ | ABO | Intron |
| | 20p12.3 | rs6038583 | chr20:6670473 | G | 0.249 | 0.086 | 0.012 | 1.09 | 1.064-1.116 | $2.74 \times 10^{-12}$ | BMP2 | Intergenic |
| Discovery GWAS | 1p36.33 | rs78717314 | chr1:1124974 | T | 0.155 | 0.115 | 0.018 | 1.122 | 1.083-1.162 | $4.20 \times 10^{-10}$ | TTLL10 | Intron |
| | 1p31.3 | rs75563559 | chr1:68422111 | T | 0.016 | 0.3 | 0.053 | 1.35 | 1.217-1.498 | $1.89 \times 10^{-8}$ | GNG12-AS1 | Intron |
| | 1q22 | rs760077 | chr1:155178782 | T | 0.862 | 0.114 | 0.019 | 1.121 | 1.08-1.163 | $3.32 \times 10^{-9}$ | MTX1 | Missense |
| | 3q13.2 | rs72943992 | chr3:112403577 | C | 0.623 | 0.096 | 0.014 | 1.101 | 1.071-1.131 | $2.28 \times 10^{-12}$ | CCDC80 | Intergenic |
| | 3q26.2 | rs77065455 | chr3:169490099 | A | 0.925 | 0.101 | 0.025 | 1.106 | 1.053-1.162 | $7.26 \times 10^{-5}$ | MYNN | Intergenic |
| | 6p22.1 | rs9260603 | chr6:29922754 | C | 0.591 | 0.095 | 0.014 | 1.1 | 1.07-1.13 | $1.84 \times 10^{-12}$ | HLA-A | Intergenic |
| | 6p21.3 | rs9271399 | chr6:32587510 | T | 0.527 | 0.105 | 0.013 | 1.111 | 1.083-1.139 | $1.91 \times 10^{-15}$ | HLA-DQA1 | Intergenic |
| | 6p21.1 | rs62396739 | chr6:41730412 | A | 0.267 | 0.102 | 0.015 | 1.107 | 1.075-1.14 | $1.40 \times 10^{-11}$ | FRS3 | Intergenic |
| | 7q21.3 | rs28457167 | chr7:97830704 | C | 0.279 | 0.079 | 0.015 | 1.082 | 1.051-1.114 | $8.84 \times 10^{-8}$ | LMTK2 | Intron |
| | 8q24.3 | rs2978981 | chr8:143759137 | T | 0.655 | 0.195 | 0.014 | 1.215 | 1.182-1.249 | $9.23 \times 10^{-44}$ | PSCA | Intron |
| | 9q34.2 | rs9411372 | chr9:136134068 | A | 0.209 | 0.071 | 0.016 | 1.074 | 1.04-1.108 | $1.68 \times 10^{-5}$ | ABO | Intron |
| | 20p12.3 | rs11087739 | chr20:6652824 | A | 0.582 | 0.061 | 0.013 | 1.063 | 1.036-1.09 | $6.24 \times 10^{-6}$ | BMP2 | Intergenic |
| Replication GWAS | 1p36.33 | rs113723849 | chr1:1133502 | T | 0.18 | 0.09 | 0.026 | 1.094 | 1.04-1.151 | $4.38 \times 10^{-4}$ | TTLL10 | Intron |
| | 1p31.3 | rs116938328 | chr1:68547919 | C | 0.025 | 0.197 | 0.061 | 1.218 | 1.081-1.372 | $1.29 \times 10^{-3}$ | GNG12-AS1 | Intron |
| | 1q22 | rs760077 | chr1:155178782 | T | 0.843 | 0.145 | 0.025 | 1.156 | 1.101-1.214 | $4.92 \times 10^{-9}$ | MTX1 | Missense |
| | 3q13.2 | rs12639600 | chr3:112392370 | G | 0.679 | 0.1 | 0.02 | 1.105 | 1.063-1.149 | $4.89 \times 10^{-7}$ | CCDC80 | Intergenic |
| | 3q26.2 | rs28626343 | chr3:169526272 | C | 0.644 | 0.091 | 0.018 | 1.095 | 1.058-1.134 | $7.32 \times 10^{-7}$ | LRRC34 | Intron |
| | 6p22.1 | rs2523969 | chr6:29938777 | A | 0.835 | 0.099 | 0.026 | 1.104 | 1.049-1.162 | $9.86 \times 10^{-5}$ | HCG9 | Intergenic |
| | 6p21.3 | rs1130153 | chr6:32610947 | A | 0.934 | 0.257 | 0.049 | 1.293 | 1.175-1.423 | $1.65 \times 10^{-7}$ | HLA-DQA1 | Intron |
| | 6p21.1 | rs149626650 | chr6:41777938 | C | 0.309 | 0.093 | 0.019 | 1.097 | 1.057-1.139 | $1.26 \times 10^{-6}$ | USP49 | Intron |
| | 7q21.3 | rs12539490 | chr7:97813089 | C | 0.739 | 0.075 | 0.022 | 1.078 | 1.032-1.125 | $7.79 \times 10^{-4}$ | LMTK2 | Intron |
| | 8q24.3 | rs2976384 | chr8:143752994 | C | 0.615 | 0.17 | 0.019 | 1.185 | 1.142-1.23 | $7.84 \times 10^{-20}$ | PSCA | Intron |
| | 9q34.2 | rs9411372 | chr9:136134068 | A | 0.229 | 0.089 | 0.023 | 1.093 | 1.045-1.143 | $9.92 \times 10^{-5}$ | ABO | Intron |
| | 20p12.3 | rs6038583 | chr20:6670473 | G | 0.253 | 0.118 | 0.021 | 1.125 | 1.08-1.173 | $7.25 \times 10^{-9}$ | BMP2 | Intergenic |

For each AG-associated region, the lead variant from the combined, discovery, and replication analyses is shown. A region was retained if it reached genome-wide significance (P-value $< 5 \times 10^{-8}$) in at least one analysis stage; therefore, not every stage-specific P value is genome-wide significant. Beta estimates are on the log-odds scale; ORs and 95% CIs were calculated as exp(beta) and exp(beta +/- 1.96 x SE). Positions are based on GRCh37. EAF, effect allele frequency; SE, standard error.

### Supplementary Table 4. Regional lead variants from the combined, discovery, and replication overall SAG GWAS.

| Analysis | Locus | Lead SNP | Position (GRCh37) | Effect allele | EAF | Beta | SE | OR | 95% CI | P-value | Nearest Gene | Annotation |
|---|---|---|---|---|---|---|---|---|---|---|---|---|
| Combined meta-GWAS | 1q22 | rs760077 | chr1:155178782 | T | 0.857 | 0.222 | 0.027 | 1.249 | 1.184-1.316 | $1.20 \times 10^{-16}$ | MTX1 | Missense |
| | 5p13.1 | rs28540420 | chr5:40687463 | C | 0.421 | 0.122 | 0.019 | 1.13 | 1.088-1.173 | $1.03 \times 10^{-10}$ | PTGER4 | Intron |
| | 6p22.1 | rs2523945 | chr6:29942324 | T | 0.586 | 0.134 | 0.022 | 1.143 | 1.095-1.194 | $5.03 \times 10^{-10}$ | HCG9 | 2KB Upstream |
| | 6p21.3 | rs9268399 | chr6:32340236 | A | 0.249 | 0.137 | 0.022 | 1.147 | 1.098-1.197 | $1.98 \times 10^{-10}$ | TSBP1 | 2KB Upstream |
| | 6q21 | rs4946699 | chr6:106263758 | A | 0.17 | 0.356 | 0.063 | 1.428 | 1.262-1.615 | $1.81 \times 10^{-8}$ | PRDM1 | Intergenic |
| | 8q24.3 | rs2920280 | chr8:143761144 | C | 0.503 | 0.326 | 0.019 | 1.385 | 1.335-1.438 | $1.43 \times 10^{-68}$ | PSCA | Intron |
| Discovery GWAS | 1q22 | rs822478 | chr1:155793969 | T | 0.805 | 0.16 | 0.028 | 1.174 | 1.111-1.24 | $1.42 \times 10^{-8}$ | GON4L | Intron |
| | 5p13.1 | rs6899032 | chr5:40839206 | C | 0.468 | 0.128 | 0.023 | 1.137 | 1.086-1.189 | $1.29 \times 10^{-8}$ | CARD6 | Intergenic |
| | 6p22.1 | rs2523945 | chr6:29942324 | T | 0.586 | 0.128 | 0.025 | 1.137 | 1.082-1.194 | $1.96 \times 10^{-7}$ | HCG9 | 2KB Upstream |
| | 6p21.3 | rs35656734 | chr6:32600153 | C | 0.699 | 0.147 | 0.024 | 1.158 | 1.105-1.214 | $1.63 \times 10^{-9}$ | HLA-DQA1 | Intron |
| | 6q21 | rs55687767 | chr6:106268447 | G | 0.986 | 0.273 | 0.098 | 1.314 | 1.084-1.592 | $5.37 \times 10^{-3}$ | PRDM1 | Intergenic |
| | 8q24.3 | rs2920280 | chr8:143761144 | C | 0.512 | 0.321 | 0.022 | 1.379 | 1.32-1.439 | $8.98 \times 10^{-47}$ | PSCA | Intron |
| Replication GWAS | 1q22 | rs760077 | chr1:155178782 | T | 0.845 | 0.32 | 0.048 | 1.377 | 1.253-1.513 | $1.70 \times 10^{-11}$ | MTX1 | Missense |
| | 5p13.1 | rs372374952 | chr5:40663259 | G | 0.02 | 0.59 | 0.177 | 1.804 | 1.275-2.552 | $8.35 \times 10^{-4}$ | PTGER4 | Intergenic |
| | 6p22.1 | rs9260093 | chr6:29909454 | C | 0.126 | 0.209 | 0.049 | 1.232 | 1.12-1.357 | $2.14 \times 10^{-5}$ | HLA-A | Intergenic |
| | 6p21.3 | rs77055720 | chr6:32396078 | T | 0.034 | 0.372 | 0.086 | 1.451 | 1.226-1.717 | $1.53 \times 10^{-5}$ | HLA-DRA | Intergenic |
| | 6q21 | rs4946699 | chr6:106263758 | A | 0.17 | 0.356 | 0.063 | 1.428 | 1.262-1.615 | $1.81 \times 10^{-8}$ | PRDM1 | Intergenic |
| | 8q24.3 | rs2717609 | chr8:143769252 | T | 0.475 | 0.347 | 0.034 | 1.415 | 1.324-1.512 | $1.45 \times 10^{-24}$ | PSCA | Intergenic |

Regional lead variants are shown for the combined, discovery, and replication analyses; lead variants can differ among stages. Beta and SE are reported for the displayed effect allele, and odds ratios and 95% confidence intervals are on the per-allele scale. Gene labels indicate nearby genes and do not imply causality. Positions are based on GRCh37. AG, atrophic gastritis; CI, confidence interval; EAF, effect allele frequency; OR, odds ratio; SAG, severe atrophic gastritis; SE, standard error.

### Supplementary Table 5. Regional lead variants from the combined, discovery, and replication anti-*H. pylori* IgG-seropositive AG GWAS.

| Analysis | Locus | Lead SNP | Position (GRCh37) | Effect allele | EAF | Beta | SE | OR | 95% CI | P-value | Nearest Gene | Annotation |
|---|---|---|---|---|---|---|---|---|---|---|---|---|
| Combined meta-GWAS | 1q22 | rs760077 | chr1:155178782 | T | 0.854 | 0.225 | 0.018 | 1.252 | 1.209-1.297 | $1.40 \times 10^{-34}$ | MTX1 | Missense |
| | 5q35.2 | rs359456 | chr5:173279808 | C | 0.428 | 0.086 | 0.013 | 1.09 | 1.062-1.118 | $2.71 \times 10^{-11}$ | CPEB4 | Intergenic |
| | 6p21.3 | rs12614 | chr6:31914179 | C | 0.934 | 0.237 | 0.028 | 1.267 | 1.2-1.339 | $9.25 \times 10^{-18}$ | CFB | Missense |
| | 6p21.1 | rs62396738 | chr6:41728997 | T | 0.279 | 0.112 | 0.014 | 1.119 | 1.088-1.15 | $9.15 \times 10^{-15}$ | PGC | Intergenic |
| | 7q21.3 | rs28457167 | chr7:97830704 | C | 0.282 | 0.083 | 0.015 | 1.087 | 1.055-1.119 | $3.10 \times 10^{-8}$ | LMTK2 | Intron |
| | 8q24.3 | rs2920279 | chr8:143762135 | C | 0.641 | 0.289 | 0.014 | 1.335 | 1.299-1.372 | $2.48 \times 10^{-101}$ | PSCA | Intron |
| | 9q34.2 | rs494242 | chr9:136145118 | T | 0.419 | 0.089 | 0.013 | 1.093 | 1.066-1.121 | $1.03 \times 10^{-11}$ | ABO | Intron |
| | 20p12.3 | rs6038583 | chr20:6670473 | G | 0.246 | 0.122 | 0.015 | 1.13 | 1.097-1.163 | $6.52 \times 10^{-16}$ | BMP2 | Intergenic |
| Discovery GWAS | 1q22 | rs760077 | chr1:155178782 | T | 0.859 | 0.197 | 0.022 | 1.218 | 1.166-1.271 | $7.41 \times 10^{-19}$ | MTX1 | Missense |
| | 5q35.2 | rs359457 | chr5:173279842 | T | 0.429 | 0.09 | 0.016 | 1.094 | 1.06-1.129 | $7.37 \times 10^{-9}$ | CPEB4 | Intergenic |
| | 6p21.3 | rs12614 | chr6:31914179 | C | 0.936 | 0.226 | 0.031 | 1.254 | 1.18-1.332 | $7.26 \times 10^{-13}$ | CFB | Missense |
| | 6p21.1 | rs2153878 | chr6:41773322 | A | 0.28 | 0.105 | 0.017 | 1.111 | 1.074-1.148 | $1.22 \times 10^{-9}$ | USP49 | Intron |
| | 7q21.3 | rs28457167 | chr7:97830704 | C | 0.28 | 0.091 | 0.017 | 1.095 | 1.059-1.132 | $1.25 \times 10^{-7}$ | LMTK2 | Intron |
| | 8q24.3 | rs2294008 | chr8:143761931 | T | 0.652 | 0.288 | 0.016 | 1.334 | 1.293-1.376 | $1.74 \times 10^{-70}$ | PSCA | Intron |
| | 9q34.2 | rs687621 | chr9:136137065 | G | 0.429 | 0.094 | 0.016 | 1.099 | 1.065-1.134 | $1.52 \times 10^{-9}$ | ABO | Intron |
| | 20p12.3 | rs6038583 | chr20:6670473 | G | 0.244 | 0.108 | 0.018 | 1.114 | 1.075-1.154 | $2.18 \times 10^{-9}$ | BMP2 | Intergenic |
| Replication GWAS | 1q22 | rs760077 | chr1:155178782 | T | 0.844 | 0.287 | 0.033 | 1.332 | 1.249-1.421 | $1.68 \times 10^{-18}$ | MTX1 | Missense |
| | 5q35.2 | rs2973351 | chr5:173320041 | T | 0.019 | 0.478 | 0.123 | 1.613 | 1.267-2.053 | $1.00 \times 10^{-4}$ | CPEB4 | Intron |
| | 6p21.3 | rs10947223 | chr6:31872383 | G | 0.927 | 0.266 | 0.054 | 1.305 | 1.174-1.45 | $7.76 \times 10^{-7}$ | C2 | Intron |
| | 6p21.1 | rs149626650 | chr6:41777938 | C | 0.309 | 0.141 | 0.026 | 1.151 | 1.094-1.212 | $3.99 \times 10^{-8}$ | USP49 | Intron |
| | 7q21.3 | rs56153131 | chr7:97847619 | T | 0.225 | 0.228 | 0.062 | 1.256 | 1.112-1.418 | $2.60 \times 10^{-4}$ | TECPR1 | Intron |
| | 8q24.3 | rs2920279 | chr8:143762135 | C | 0.617 | 0.293 | 0.025 | 1.34 | 1.276-1.408 | $1.03 \times 10^{-32}$ | PSCA | Intron |
| | 9q34.2 | rs9411372 | chr9:136134068 | A | 0.228 | 0.152 | 0.032 | 1.164 | 1.093-1.24 | $2.55 \times 10^{-6}$ | ABO | Intron |
| | 20p12.3 | rs6038586 | chr20:6673320 | A | 0.25 | 0.153 | 0.027 | 1.165 | 1.105-1.229 | $1.71 \times 10^{-8}$ | BMP2 | Intergenic |

Regional lead variants are shown for the combined, discovery, and replication analyses; lead variants can differ among stages. Beta and SE are reported for the displayed effect allele, and odds ratios and 95% confidence intervals are on the per-allele scale. Gene labels indicate nearby genes and do not imply causality. Positions are based on GRCh37. AG, atrophic gastritis; CI, confidence interval; EAF, effect allele frequency; OR, odds ratio; SAG, severe atrophic gastritis; SE, standard error. Anti-*H. pylori* IgG seropositivity was defined as >3 U/mL and does not directly establish active infection.

**Supplementary Table 6. Regional lead variants from the combined, discovery, and replication anti-*H. pylori* IgG-seropositive SAG GWAS.**

| Analysis | Locus | Lead SNP | Position (GRCh37) | Effect allele | EAF | Beta | SE | OR | 95% CI | P-value | Nearest Gene | Annotation |
|---|---|---|---|---|---|---|---|---|---|---|---|---|
| Combined meta-GWAS | 1q22 | rs760077 | chr1:155178782 | T | 0.854 | 0.318 | 0.029 | 1.374 | 1.298-1.455 | $4.10 \times 10^{-27}$ | MTX1 | Missense |
| | 5p13.1 | rs28540420 | chr5:40687463 | C | 0.421 | 0.124 | 0.021 | 1.132 | 1.086-1.18 | $1.78 \times 10^{-9}$ | PTGER4 | Intron |
| | 6p21.3 | rs6940063 | chr6:32428862 | C | 0.069 | 0.226 | 0.04 | 1.254 | 1.159-1.356 | $1.47 \times 10^{-8}$ | HLA-DRA | Intergenic |
| | 8q24.3 | rs2920280 | chr8:143761144 | C | 0.497 | 0.39 | 0.02 | 1.477 | 1.42-1.536 | $4.60 \times 10^{-81}$ | PSCA | Intron |
| Discovery GWAS | 1q22 | rs760077 | chr1:155178782 | T | 0.859 | 0.277 | 0.036 | 1.319 | 1.229-1.416 | $1.11 \times 10^{-14}$ | MTX1 | Missense |
| | 5p13.1 | rs28540420 | chr5:40687463 | C | 0.424 | 0.132 | 0.025 | 1.141 | 1.087-1.198 | $1.49 \times 10^{-7}$ | PTGER4 | Intron |
| | 6p21.3 | rs74729707 | chr6:32566501 | C | 0.723 | 0.137 | 0.028 | 1.147 | 1.086-1.212 | $6.52 \times 10^{-7}$ | HLA-DRB1 | Intergenic |
| | 8q24.3 | rs2920280 | chr8:143761144 | C | 0.506 | 0.387 | 0.025 | 1.473 | 1.402-1.547 | $3.45 \times 10^{-55}$ | PSCA | Intron |
| Replication GWAS | 1q22 | rs760077 | chr1:155178782 | T | 0.844 | 0.402 | 0.052 | 1.495 | 1.35-1.655 | $7.11 \times 10^{-15}$ | MTX1 | Missense |
| | 5p13.1 | rs372374952 | chr5:40663259 | G | 0.019 | 0.624 | 0.19 | 1.866 | 1.286-2.709 | $9.96 \times 10^{-4}$ | PTGER4 | Intergenic |
| | 6p21.3 | rs77055720 | chr6:32396078 | T | 0.034 | 0.398 | 0.093 | 1.489 | 1.241-1.787 | $1.71 \times 10^{-5}$ | HLA-DRA | Intergenic |
| | 8q24.3 | rs2717609 | chr8:143769252 | T | 0.468 | 0.403 | 0.037 | 1.496 | 1.392-1.609 | $2.33 \times 10^{-28}$ | PSCA | Intergenic |

Regional lead variants are shown for the combined, discovery, and replication analyses; lead variants can differ among stages. Beta and SE are reported for the displayed effect allele, and odds ratios and 95% confidence intervals are on the per-allele scale. Gene labels indicate nearby genes and do not imply causality. Positions are based on GRCh37. AG, atrophic gastritis; CI, confidence interval; EAF, effect allele frequency; OR, odds ratio; SAG, severe atrophic gastritis; SE, standard error. Anti-*H. pylori* IgG seropositivity was defined as >3 U/mL and does not directly establish active infection.

### Supplementary Table 7. Regional lead variants from the combined, discovery, and replication anti-*H. pylori* IgG-seronegative AG GWAS.

| Analysis | Locus | Lead SNP | Position (GRCh37) | Effect allele | EAF | Beta | SE | OR | 95% CI | P-value | Nearest Gene | Annotation |
|---|---|---|---|---|---|---|---|---|---|---|---|---|
| Combined meta-GWAS | 6p21.3 | rs9271065 | chr6:32575619 | C | 0.101 | 0.587 | 0.067 | 1.799 | 1.577-2.051 | $1.40 \times 10^{-18}$ | HLA-DRB1 | Intergenic |
| Discovery GWAS | 6p21.3 | rs9469110 | chr6:32398525 | T | 0.151 | 0.535 | 0.063 | 1.707 | 1.509-1.932 | $1.43 \times 10^{-17}$ | HLA-DRA | Intergenic |
| Replication GWAS | 6p21.3 | rs9269204 | chr6:32449797 | C | 0.256 | 0.51 | 0.107 | 1.665 | 1.35-2.054 | $1.71 \times 10^{-6}$ | HLA-DRB5 | Intergenic |

Regional lead variants are shown for the combined, discovery, and replication analyses; lead variants can differ among stages. Beta and SE are reported for the displayed effect allele, and odds ratios and 95% confidence intervals are on the per-allele scale. Gene labels indicate nearby genes and do not imply causality. Positions are based on GRCh37. AG, atrophic gastritis; CI, confidence interval; EAF, effect allele frequency; OR, odds ratio; SAG, severe atrophic gastritis; SE, standard error. Anti-*H. pylori* IgG seronegativity was defined as <=3 U/mL and does not directly establish absence of current or previous infection.

**Supplementary Table 8. Regional lead variants from the combined, discovery, and replication anti-*H. pylori* IgG-seronegative SAG GWAS.**

| Analysis | Locus | Lead SNP | Position (GRCh37) | Effect allele | EAF | Beta | SE | OR | 95% CI | P-value | Nearest Gene | Annotation |
|---|---|---|---|---|---|---|---|---|---|---|---|---|
| Combined meta-GWAS | 2q24.2 | rs11679244 | chr2:163082395 | A | 0.172 | 0.415 | 0.076 | 1.514 | 1.305-1.758 | $4.21 \times 10^{-8}$ | FAP | Intron |
| | 6p22.1 | rs9258242 | chr6:29716146 | G | 0.818 | 0.485 | 0.07 | 1.624 | 1.416-1.863 | $5.93 \times 10^{-12}$ | HLA-F-AS1 | Intron |
| | 6p21.3 | rs9269047 | chr6:32438783 | C | 0.096 | 0.863 | 0.089 | 2.37 | 1.991-2.822 | $4.74 \times 10^{-22}$ | HLA-DRA | Intergenic |
| | 6q27 | rs204295 | chr6:167500562 | C | 0.536 | 0.3 | 0.053 | 1.35 | 1.217-1.498 | $1.46 \times 10^{-8}$ | CCR6 | Intergenic |
| | 8q24.12 | rs370391108 | chr8:120562304 | G | 0.017 | 0.631 | 0.226 | 1.879 | 1.207-2.927 | $5.27 \times 10^{-3}$ | ENPP2 | Intergenic |
| | 19q13.12 | rs725986 | chr19:36057622 | T | 0.82 | 0.462 | 0.075 | 1.587 | 1.37-1.839 | $8.58 \times 10^{-10}$ | ATP4A | Intergenic |
| Discovery GWAS | 2q24.2 | rs11679244 | chr2:163082395 | A | 0.172 | 0.422 | 0.083 | 1.525 | 1.296-1.794 | $4.39 \times 10^{-7}$ | FAP | Intron |
| | 6p22.1 | rs9258242 | chr6:29716146 | G | 0.826 | 0.483 | 0.079 | 1.621 | 1.388-1.892 | $1.12 \times 10^{-9}$ | HLA-F-AS1 | Intron |
| | 6p21.3 | rs9469110 | chr6:32398525 | T | 0.151 | 0.796 | 0.086 | 2.217 | 1.873-2.624 | $2.24 \times 10^{-20}$ | HLA-DRA | Intergenic |
| | 6q27 | rs239933 | chr6:167413230 | G | 0.544 | 0.271 | 0.059 | 1.311 | 1.168-1.472 | $4.80 \times 10^{-6}$ | CEP43 | Intron |
| | 8q24.12 | rs149499850 | chr8:120576658 | G | 0.023 | 0.501 | 0.195 | 1.65 | 1.126-2.419 | $1.00 \times 10^{-2}$ | ENPP2 | Intron |
| | 19q13.12 | rs11882733 | chr19:36056253 | A | 0.819 | 0.44 | 0.082 | 1.553 | 1.322-1.823 | $9.05 \times 10^{-8}$ | ATP4A | 2KB Upstream |
| Replication GWAS | 2q24.2 | rs572746168 | chr2:163058336 | C | 0.015 | 1.723 | 0.586 | 5.601 | 1.776-17.665 | $3.26 \times 10^{-3}$ | FAP | Intron |
| | 6p22.1 | rs9258223 | chr6:29712175 | C | 0.784 | 0.514 | 0.149 | 1.672 | 1.249-2.239 | $5.79 \times 10^{-4}$ | HLA-F-AS1 | Intron |
| | 6p21.3 | rs3104373 | chr6:32600375 | T | 0.099 | 0.775 | 0.172 | 2.171 | 1.549-3.041 | $7.04 \times 10^{-6}$ | HLA-DQA1 | Intron |
| | 6q27 | rs6902119 | chr6:167505791 | C | 0.425 | 0.51 | 0.12 | 1.665 | 1.316-2.107 | $2.05 \times 10^{-5}$ | CCR6 | Intergenic |
| | 8q24.12 | rs78903306 | chr8:120712964 | T | 0.027 | 1.627 | 0.294 | 5.089 | 2.86-9.054 | $3.01 \times 10^{-8}$ | ENPP2 | Intergenic |
| | 19q13.12 | rs10424120 | chr19:36068881 | A | 0.515 | 0.368 | 0.111 | 1.445 | 1.162-1.796 | $9.13 \times 10^{-4}$ | ATP4A | Intergenic |

Regional lead variants are shown for the combined, discovery, and replication analyses; lead variants can differ among stages. Beta and SE are reported for the displayed effect allele, and odds ratios and 95% confidence intervals are on the per-allele scale. Gene labels indicate nearby genes and do not imply causality. Positions are based on GRCh37. AG, atrophic gastritis; CI, confidence interval; EAF, effect allele frequency; OR, odds ratio; SAG, severe atrophic gastritis; SE, standard error. Anti-*H. pylori* IgG seronegativity was defined as <=3 U/mL and does not directly establish absence of current or previous infection.

**Supplementary Table 9. FINEMAP results for genome-wide significant locus-analysis combinations across the 17 Japanese AG/SAG-associated loci.**

| Locus | Combined GWAS analysis | Lead variant (anchor) | GWAS P-value | Locus label | Anchor position (GRCh37) | Fine-mapping window (GRCh37) | Variants in LD matrix | Credible sets (n) | CS1 | CS2 | CS3 | CS4 | CS5 | Representative credible-set variant (highest r2 with anchor) | LD r2 between representative and anchor variant | Distance from anchor variant (bp) |
|---|---|---|---|---|---|---|---|---|---|---|---|---|---|---|---|---|
| 1p36.33 | AG | rs58980341 | $3.35 \times 10^{-11}$ | TTLL10 | chr1:1135107 | chr1:1000000-1300000 | 984 | 5 | rs77156226 (PP=0.9927; n=1) | rs57839043 (PP=1.0000; n=1) | rs2298210 (PP=1.0000; n=1) | rs59574701 (PP=0.5085; n=4) | rs3813200 (PP=1.0000; n=1) | rs59574701 (PP=0.5085; CS4; n=4) | 0.992 | 4,796 |
| 1q22 | AG | rs760077 | $1.45 \times 10^{-16}$ | MTX1 | chr1:155178782 | chr1:155000000-155300000 | 367 | 5 | rs1611774 (PP=1.0000; n=1) | rs2974937 (PP=1.0000; n=1) | rs2070803 (PP=1.0000; n=1) | rs7556304 (PP=1.0000; n=1) | rs4971091 (PP=1.0000; n=1) | rs2974937 (PP=1.0000; CS2; n=1) | 0.935 | 9,933 |
| | SAG | | $1.20 \times 10^{-16}$ | | | | 372 | 5 | rs423144 (PP=1.0000; n=1) | rs2974937 (PP=1.0000; n=1) | rs67579710 (PP=1.0000; n=1) | rs11264341 (PP=1.0000; n=1) | rs3766917 (PP=0.9977; n=1) | rs2974937 (PP=1.0000; CS2; n=1) | | |
| | HP+ AG | | $1.40 \times 10^{-34}$ | | | | 370 | 5 | rs4971091 (PP=1.0000; n=1) | rs7556304 (PP=1.0000; n=1) | rs12411216 (PP=0.9908; n=1) | rs58939130 (PP=1.0000; n=1) | rs4971099 (PP=1.0000; n=1) | rs12411216 (PP=0.9908; CS3; n=1) | 0.935 | 14,302 |
| | HP+ SAG | | $4.10 \times 10^{-27}$ | | | | 370 | 5 | rs4971091 (PP=1.0000; n=1) | rs7556304 (PP=1.0000; n=1) | rs12411216 (PP=0.9299; n=2) | rs4971099 (PP=1.0000; n=1) | rs141942982 (PP=1.0000; n=1) | rs12411216 (PP=0.9299; CS3; n=2) | | |
| 2q24.2 | HP- SAG | rs11679244 | $4.21 \times 10^{-8}$ | FAP | chr2:163082395 | chr2:162950000-163250000 | 486 | 3 | rs2111485 (PP=1.0000; n=1) | rs1990760 (PP=0.9999; n=1) | rs148412518 (PP=0.9532; n=1) | | | rs2111485 (PP=1.0000; CS1; n=1) | 0.786 | 28,141 |
| 3q13.2 | AG | rs72943992 | $8.19 \times 10^{-18}$ | CCDC80 | chr3:112403577 | chr3:112250000-112550000 | 873 | 5 | rs72943992 (PP=1.0000; n=1) | rs75439410 (PP=1.0000; n=1) | rs9876497 (PP=0.9935; n=1) | rs9870933 (PP=1.0000; n=1) | rs7630314 (PP=0.9999; n=1) | rs72943992 (PP=1.0000; CS1; n=1) | 1.000 | 0 |
| 3q26.2 | AG | rs28626343 | $4.59 \times 10^{-8}$ | LRRC34 | chr3:169526272 | chr3:169400000-169700000 | 671 | 1 | rs28626343 (PP=0.2086; n=9) | | | | | rs28626343 (PP=0.2086; CS1; n=9) | 1.000 | 0 |
| 5p13.1 | SAG | rs28540420 | $1.03 \times 10^{-10}$ | PTGER4 | chr5:40687463 | chr5:40600000-40900000 | 848 | 5 | rs78356883 (PP=0.5000; n=2) | rs2329353 (PP=1.0000; n=1) | rs62356468 (PP=1.0000; n=1) | rs462366 (PP=0.9902; n=1) | rs73084490 (PP=0.8767; n=2) | rs2329353 (PP=1.0000; CS2; n=1) | 0.992 | 60,805 |
| | HP+ SAG | | $1.78 \times 10^{-9}$ | | | | 848 | 5 | rs2329353 (PP=1.0000; n=1) | rs77567692 (PP=1.0000; n=1) | rs142676770 (PP=1.0000; n=1) | rs75671110 (PP=0.9937; n=1) | rs73084490 (PP=1.0000; n=1) | rs2329353 (PP=1.0000; CS1; n=1) | | |
| 5q35.2 | HP+ AG | rs359456 | $2.71 \times 10^{-11}$ | CPEB4 | chr5:173279808 | chr5:173150000-173450000 | 876 | 5 | rs111991836 (PP=1.0000; n=1) | rs2922298 (PP=0.6164; n=2) | rs17076724 (PP=0.9999; n=1) | rs359456 (PP=0.9997; n=1) | rs201122529 (PP=1.0000; n=1) | rs359456 (PP=0.9997; CS4; n=1) | 1.000 | 0 |
| 6p22.1 | AG | rs9260603 | $9.20 \times 10^{-14}$ | HLA-A | chr6:29922754 | chr6:29800000-30000000 | 3922 | 5 | rs393282 (PP=1.0000; n=1) | rs3823374 (PP=1.0000; n=1) | rs400488 (PP=1.0000; n=1) | rs4947244 (PP=1.0000; n=1) | rs1974574 (PP=1.0000; n=1) | rs393282 (PP=1.0000; CS1; n=1) | 0.833 | 19,743 |
| | SAG | rs2523945 | $5.03 \times 10^{-10}$ | | chr6:29942324 | | 3936 | 5 | rs56052612 (PP=1.0000; n=1) | rs6922640 (PP=1.0000; n=1) | rs6923856 (PP=1.0000; n=1) | rs9260918 (PP=0.9968; n=1) | rs3734836 (PP=0.7670; n=2) | rs56052612 (PP=1.0000; CS1; n=1) | 0.214 | 3,478 |
| | HP+ AG | rs75292910 | $1.17 \times 10^{-13}$ | | chr6:29807788 | | 3942 | 5 | rs6923856 (PP=1.0000; n=1) | rs11964113 (PP=1.0000; n=1) | rs9260959 (PP=0.8549; n=2) | rs9256989 (PP=1.0000; n=1) | rs9260740 (PP=1.0000; n=1) | rs11964113 (PP=1.0000; CS2; n=1) | 0.450 | 163,503 |
| 6p21.3 | HP+ AG | rs12614 | $9.25 \times 10^{-18}$ | CFB | chr6:31914179 | chr6:31764179-32064179 | 620 | 5 | rs12614 (PP=1.0000; n=1) | rs11751545 (PP=0.9224; n=2) | rs9501161 (PP=0.5126; n=2) | rs9283893 (PP=0.6429; n=2) | rs13194087 (PP=0.6474; n=2) | rs12614 (PP=1.0000; CS1; n=1) | 1.000 | 0 |
| | AG | rs73729294 | $4.78 \times 10^{-14}$ | HLA-DRA | chr6:32445447 | chr6:32400000-32500000 | 3134 | 5 | rs12204922 (PP=1.0000; n=1) | rs76733997 (PP=1.0000; n=1) | rs373105177 (PP=0.9871; n=1) | rs536151678 (PP=1.0000; n=1) | rs561623787 (PP=1.0000; n=1) | rs536151678 (PP=1.0000; CS4; n=1) | 0.129 | 8,096 |
| | SAG | rs2395184 | $6.22 \times 10^{-10}$ | | chr6:32432646 | | 3133 | 5 | rs17203444 (PP=0.5000; n=2) | rs1557548 (PP=0.7183; n=2) | rs575694449 (PP=0.5818; n=2) | rs2395184 (PP=1.0000; n=1) | rs62405568 (PP=1.0000; n=1) | rs2395184 (PP=1.0000; CS4; n=1) | 1.000 | 0 |
| | HP+ SAG | rs6940063 | $1.47 \times 10^{-8}$ | | chr6:32428862 | | 3150 | 5 | rs17203444 (PP=0.5000; n=2) | rs143909993 (PP=0.9374; n=2) | rs111524667 (PP=0.5928; n=2) | rs10947290 (PP=1.0000; n=1) | rs17209768 (PP=0.4014; n=4) | rs62405568 (PP=0.0959; CS5; n=4) | 0.610 | 24,547 |
| | HP- AG | rs9269047 | $3.25 \times 10^{-18}$ | | chr6:32438783 | | 3149 | 5 | rs9269047 (PP=0.9598; n=1) | rs111752979 (PP=0.3431; n=3) | rs67987819 (PP=1.0000; n=1) | rs57404925 (PP=1.0000; n=1) | rs9269236 (PP=1.0000; n=1) | rs9269047 (PP=0.9598; CS1; n=1) | 1.000 | 0 |
| | HP- SAG | | $4.74 \times 10^{-22}$ | | | | 3145 | 5 | rs9269047 (PP=0.7555; n=3) | rs146942103 (PP=0.3435; n=3) | rs57404925 (PP=1.0000; n=1) | rs67987819 (PP=1.0000; n=1) | rs9269236 (PP=1.0000; n=1) | rs9269047 (PP=0.7555; CS1; n=3) | | |
| 6p21.1 | AG | rs62396739 | $4.23 \times 10^{-16}$ | PGC | chr6:41730412 | chr6:41600000-41900000 | 575 | 5 | rs62396738 (PP=1.0000; n=1) | rs3761781 (PP=1.0000; n=1) | rs2153878 (PP=1.0000; n=1) | rs55967101 (PP=1.0000; n=1) | rs117356949 (PP=0.6265; n=3) | rs62396738 (PP=1.0000; CS1; n=1) | 0.918 | 1,415 |
| | HP+ AG | rs62396738 | $9.15 \times 10^{-15}$ | | chr6:41728997 | | 576 | 5 | rs62396739 (PP=0.9996; n=1) | rs3761781 (PP=1.0000; n=1) | rs68017625 (PP=0.7955; n=2) | rs11962743 (PP=1.0000; n=1) | rs55967101 (PP=0.9883; n=1) | rs62396739 (PP=0.9996; CS1; n=1) | 0.918 | 1,415 |
| 6q21 | SAG | rs4946699 | $1.81 \times 10^{-8}$ | PRDM1 | chr6:106263758 | chr6:106200000-106400000 | 641 | 5 | rs76770617 (PP=1.0000; n=1) | rs73510172 (PP=1.0000; n=1) | rs78758811 (PP=0.9385; n=2) | rs4946699 (PP=1.0000; n=1) | rs78615796 (PP=1.0000; n=1) | rs4946699 (PP=1.0000; CS4; n=1) | 1.000 | 0 |
| 6q27 | HP- SAG | rs204295 | $1.46 \times 10^{-8}$ | CCR6 | chr6:167500562 | chr6:167300000-167600000 | 1193 | 5 | rs137955076 (PP=1.0000; n=1) | rs9457265 (PP=0.8795; n=2) | rs7748224 (PP=0.5216; n=3) | rs387733 (PP=0.4138; n=11) | rs6920364 (PP=0.9264; n=2) | rs9457265 (PP=0.8795; CS2; n=2) | 0.984 | 19,753 |
| 7q21.3 | AG | rs58880085 | $3.05 \times 10^{-8}$ | LMTK2 | chr7:97836465 | chr7:97700000-98050000 | 861 | 5 | rs12536312 (PP=1.0000; n=1) | rs554599272 (PP=1.0000; n=1) | rs80327648 (PP=1.0000; n=1) | rs35943072 (PP=1.0000; n=1) | rs80021450 (PP=0.4460; n=3) | rs80021450 (PP=0.4460; CS5; n=3) | 0.965 | 4,476 |
| | HP+ AG | rs28457167 | $3.10 \times 10^{-8}$ | | chr7:97830704 | | 862 | 5 | rs10953258 (PP=1.0000; n=1) | rs1985593 (PP=0.7938; n=2) | rs6465667 (PP=0.9709; n=1) | rs7802794 (PP=0.4585; n=5) | rs10953254 (PP=0.6308; n=71) | rs28457167 (PP=0.0043; CS5; n=71) | 1.000 | 0 |
| 8q24.3 | AG | rs2920279 | $3.32 \times 10^{-61}$ | PSCA | chr8:143762135 | chr8:143600000-143900000 | 1000 | 5 | rs2920285 (PP=1.0000; n=1) | rs146194718 (PP=0.9970; n=1) | rs2978977 (PP=1.0000; n=1) | rs2585177 (PP=1.0000; n=1) | rs78563521 (PP=1.0000; n=1) | rs2920285 (PP=1.0000; CS1; n=1) | 0.991 | 5,917 |
| | HP+ AG | | $2.48 \times 10^{-101}$ | | | | 1002 | 5 | rs2920279 (PP=1.0000; n=1) | rs2976388 (PP=1.0000; n=1) | rs146194718 (PP=0.9978; n=1) | rs2294012 (PP=0.9847; n=1) | rs78146008 (PP=1.0000; n=1) | rs2920279 (PP=1.0000; CS1; n=1) | 1.000 | 0 |
| | SAG | rs2920280 | $1.43 \times 10^{-68}$ | | chr8:143761144 | | 1002 | 5 | rs2920279 (PP=1.0000; n=1) | rs2976388 (PP=1.0000; n=1) | rs73714659 (PP=1.0000; n=1) | rs141266717 (PP=1.0000; n=1) | rs189393533 (PP=1.0000; n=1) | rs2976388 (PP=1.0000; CS2; n=1) | 1.000 | 888 |
| | HP+ SAG | | $4.60 \times 10^{-81}$ | | | | 1001 | 5 | rs2920279 (PP=1.0000; n=1) | rs2976388 (PP=1.0000; n=1) | rs73714659 (PP=1.0000; n=1) | rs2294012 (PP=0.7790; n=3) | rs78146008 (PP=1.0000; n=1) | rs2976388 (PP=1.0000; CS2; n=1) | | |
| 9q34.2 | AG | rs9411372 | $8.19 \times 10^{-9}$ | ABO | chr9:136134068 | chr9:136050000-136200000 | 742 | 5 | rs1053878 (PP=0.9669; n=1) | rs9411473 (PP=0.9402; n=2) | rs61527302 (PP=1.0000; n=1) | rs9411476 (PP=1.0000; n=1) | rs587600964 (PP=0.7912; n=11) | rs1053878 (PP=0.9669; CS1; n=1) | 0.855 | 2,417 |
| | HP+ AG | rs494242 | $1.03 \times 10^{-11}$ | ABO | chr9:136145118 | | 744 | 5 | rs687621 (PP=1.0000; n=1) | rs7855466 (PP=0.5585; n=3) | rs13292932 (PP=0.5235; n=2) | rs10901251 (PP=1.0000; n=1) | rs9411365 (PP=0.9999; n=1) | rs687621 (PP=1.0000; CS1; n=1) | 0.942 | 8,053 |
| 19q13.12 | HP- SAG | rs725986 | $8.58 \times 10^{-10}$ | ATP4A | chr19:36057622 | chr19:35900000-36200000 | 844 | 5 | rs73597296 (PP=1.0000; n=1) | rs10422358 (PP=1.0000; n=1) | rs12979142 (PP=1.0000; n=1) | rs12973004 (PP=1.0000; n=1) | rs58800588 (PP=0.9616; n=1) | rs73597296 (PP=1.0000; CS1; n=1) | 0.958 | 3,881 |
| 20p12.3 | AG | rs6038583 | $2.74 \times 10^{-12}$ | BMP2 | chr20:6670473 | chr20:6550000-6850000 | 758 | 5 | rs6140071 (PP=1.0000; n=1) | rs3931450 (PP=1.0000; n=1) | rs235762 (PP=1.0000; n=1) | rs2876050 (PP=0.3107; n=9) | rs6117401 (PP=1.0000; n=1) | rs6038583 (PP=0.0737; CS4; n=9) | 1.000 | 0 |
| | HP+ AG | | $6.52 \times 10^{-16}$ | | | | 760 | 5 | rs6054459 (PP=0.4128; n=3) | rs6085649 (PP=0.6188; n=2) | rs1555163 (PP=0.9796; n=1) | rs1986180 (PP=1.0000; n=1) | rs990999 (PP=0.9669; n=1) | rs6054459 (PP=0.4128; CS1; n=3) | 0.783 | 92 |

FINEMAP was applied to 17 loci across six combined GWAS analyses (102 analyses) and to a separate CFB target across the same six analyses, yielding 108 analyses. The table reports the 35 locus-analysis combinations for which the anchor variant reached $P < 5 \times 10^{-8}$. LD was estimated from the 1000 Genomes Project Phase 3 East Asian reference panel, and the maximum number of causal components was set to five. PP is the posterior probability reported for the displayed representative variant within the corresponding credible set; n is the number of variants in that credible set. CS1–CS5 show one representative variant from each credible set, its within-set posterior probability (PP), and the number of variants in that set (n). The LD representative was selected from all variants belonging to the reported credible sets as the variant with the highest pairwise $r^2$ with the GWAS anchor. Distance is the absolute base-pair distance between the representative and the anchor. Reaching five credible sets indicates that the prespecified upper bound was reached and does not establish five independent causal variants. Fine-mapping prioritizes candidate variants but does not establish causality. Positions are based on GRCh37. CS, credible set; LD, linkage disequilibrium; PP, posterior probability.

### Supplementary Table 10. Odds ratios by HP antibody status and effect-allele dosage.

| Locus | Variant | Effect allele | Trait | Joint OR (common reference) | | | | Allele OR within each HP status | |
|---|---|---|---|---|---|---|---|---|---|
| | | | | HP antibody-negative | | HP antibody-positive | | HP antibody-negative | HP antibody-positive |
| | | | | 0 copies | 1 copy | 0 copies | 1 copy | | |
| *MTX1* | rs760077 | T | AG | 1.000 (ref) | 0.923 | 10.844 | 13.451 | 0.923 | 1.240 |
| *FAP* | rs11679244 | A | | 1.000 (ref) | 1.243 | 19.703 | 20.194 | 1.243 | 1.025 |
| *ACKR2* | rs150595426 | A | | 1.000 (ref) | 1.636 | 18.705 | 15.391 | 1.636 | 0.823 |
| *ZNRD1-AS1* | 6:29968499:C:G | G | | 1.000 (ref) | 1.352 | 19.846 | 20.969 | 1.352 | 1.057 |
| *HLA-C* | rs3130425 | C | | 1.000 (ref) | 1.174 | 21.452 | 21.793 | 1.174 | 1.016 |
| *TSBP1* | rs9268399 | A | | 1.000 (ref) | 1.374 | 21.299 | 21.906 | 1.374 | 1.029 |
| *HLA-DRA* | rs9469110 | T | | 1.000 (ref) | 1.509 | 21.296 | 19.695 | 1.509 | 0.925 |
| *HLA-DRA* | rs2027856 | A | | 1.000 (ref) | 1.509 | 21.299 | 19.687 | 1.509 | 0.924 |
| *PSCA* | rs2920280 | C | | 1.000 (ref) | 1.224 | 17.115 | 22.182 | 1.224 | 1.296 |
| *ATP4A* | rs725985 | A | | 1.000 (ref) | 1.232 | 27.762 | 26.500 | 1.232 | 0.955 |
| *MTX1* | rs760077 | T | SAG | 1.000 (ref) | 0.799 | 2.095 | 2.907 | 0.799 | 1.388 |
| *FAP* | rs11679244 | A | | 1.000 (ref) | 1.440 | 6.113 | 6.761 | 1.440 | 1.106 |
| *ACKR2* | rs150595426 | A | | 1.000 (ref) | 1.540 | 5.624 | 4.899 | 1.540 | 0.871 |
| *ZNRD1-AS1* | 6:29968499:C:G | G | | 1.000 (ref) | 1.523 | 6.039 | 6.890 | 1.523 | 1.141 |
| *HLA-C* | rs3130425 | C | | 1.000 (ref) | 1.522 | 9.227 | 9.357 | 1.522 | 1.014 |
| *TSBP1* | rs9268399 | A | | 1.000 (ref) | 1.698 | 7.376 | 7.774 | 1.698 | 1.054 |
| *HLA-DRA* | rs9469110 | T | | 1.000 (ref) | 1.885 | 7.119 | 6.714 | 1.885 | 0.943 |
| *HLA-DRA* | rs2027856 | A | | 1.000 (ref) | 1.884 | 7.118 | 6.714 | 1.884 | 0.943 |
| *PSCA* | rs2920280 | C | | 1.000 (ref) | 1.218 | 4.365 | 6.471 | 1.218 | 1.482 |
| *ATP4A* | rs725985 | A | | 1.000 (ref) | 1.754 | 14.279 | 14.099 | 1.754 | 0.987 |

Joint ORs are approximate model-derived estimates, relative to HP antibody-negative participants with 0 copies of the effect allele. Environmental coefficients were converted using dataset-specific standardization and pooled by inverse variance. FAP, ZNRD1-AS1 and ATP4A include 4 datasets (summed N = 69,753); other variants include 6 (89,878). HP, anti-*H. pylori* IgG antibody status; AG, atrophic gastritis; SAG, severe atrophic gastritis; OR, odds ratio.

**Supplementary Table 11. Associations of genetically inferred ABO blood groups with AG/SAG and anti-*H. pylori* IgG seropositivity.**

| Analysis | Comparison | Reference | OR | 95% CI | Unadjusted P-value | Overall 3-df LRT unadjusted P-value | Bonferroni-adjusted P-value | Overall 3-df LRT Bonferroni-adjusted P-value |
|---|---|---|---|---|---|---|---|---|
| All AG | A | O | 1.087 | 1.038-1.137 | $3.40 \times 10^{-4}$ | $3.04 \times 10^{-5}$ | $7.13 \times 10^{-3}$ | $2.13 \times 10^{-4}$ |
| | B | | 1.001 | 0.951-1.054 | 0.965 | | 1.000 | |
| | AB | | 1.131 | 1.055-1.212 | $5.52 \times 10^{-4}$ | | 0.012 | |
| All SAG | A | | 1.046 | 0.967-1.132 | 0.257 | 0.157 | 1.000 | 1.000 |
| | B | | 1.005 | 0.920-1.099 | 0.909 | | 1.000 | |
| | AB | | 1.136 | 1.009-1.279 | 0.035 | | 0.725 | |
| Seropositive AG | A | O | 1.139 | 1.081-1.201 | $1.27 \times 10^{-6}$ | $1.72 \times 10^{-7}$ | $2.67 \times 10^{-5}$ | $1.20 \times 10^{-6}$ |
| | B | | 1.08 | 1.017-1.147 | 0.012 | | 0.255 | |
| | AB | | 1.219 | 1.123-1.324 | $2.27 \times 10^{-6}$ | | $4.77 \times 10^{-5}$ | |
| Seropositive SAG | A | | 1.074 | 0.985-1.171 | 0.108 | 0.186 | 1.000 | 1.000 |
| | B | | 1.077 | 0.976-1.188 | 0.139 | | 1.000 | |
| | AB | | 1.133 | 0.992-1.294 | 0.065 | | 1.000 | |
| Seronegative AG | A | O | 0.995 | 0.859-1.153 | 0.947 | 0.049 | 1.000 | 0.341 |
| | B | | 0.867 | 0.731-1.028 | 0.100 | | 1.000 | |
| | AB | | 1.196 | 0.967-1.479 | 0.098 | | 1.000 | |
| Seronegative SAG | A | | 0.98 | 0.800-1.201 | 0.845 | 0.019 | 1.000 | 0.131 |
| | B | | 0.822 | 0.648-1.043 | 0.107 | | 1.000 | |
| | AB | | 1.35 | 1.021-1.786 | 0.035 | | 0.741 | |
| Anti-*H. pylori* IgG seropositivity | A | O | 0.994 | 0.954-1.035 | 0.765 | $8.15 \times 10^{-3}$ | 1.000 | 0.057 |
| | B | | 0.949 | 0.906-0.994 | 0.027 | | 0.559 | |
| | AB | | 0.913 | 0.856-0.973 | $5.46 \times 10^{-3}$ | | 0.115 | |

ABO blood groups were genetically inferred from rs505922, rs8176746, and rs8176747 as described in Methods. O blood group was the reference category. Logistic regression models were adjusted for age, sex, and principal components 1-10. The overall association of ABO blood group was evaluated by a 3-df likelihood-ratio test comparing models with and without the A, B, and AB indicators. Anti-*H. pylori* IgG seropositivity was defined as >3 U/mL. ORs and 95% CIs are shown for each blood group relative to O. AG, atrophic gastritis; CI, confidence interval; LRT, likelihood-ratio test; OR, odds ratio; SAG, severe atrophic gastritis.

Bonferroni correction was applied separately across 21 blood-group contrasts and seven omnibus tests. The 95% CIs are unadjusted.

**Supplementary Table 12. Regional lead variants from the BBJ-excluded Japanese overall AG meta-analysis used in AG-gastric cancer shared-genetic analyses.**

| Locus | Lead SNP | Position (GRCh37) | Effect allele | EAF | Beta | SE | OR | 95% CI | P-value | Nearest Gene | Annotation |
|---|---|---|---|---|---|---|---|---|---|---|---|
| 1p36.33 | rs78717314 | chr1:1124974 | T | 0.155 | 0.109 | 0.016 | 1.115 | 1.081-1.151 | $9.71 \times 10^{-12}$ | TTLL10 | Intron |
| 1p31.3 | rs75563559 | chr1:68422111 | T | 0.016 | 0.253 | 0.045 | 1.288 | 1.179-1.408 | $2.26 \times 10^{-8}$ | GNG12-AS1 | Intron |
| 1q21.3 | rs12030974 | chr1:151798295 | A | 0.379 | 0.069 | 0.012 | 1.071 | 1.046-1.097 | $9.96 \times 10^{-9}$ | RORC | Intron |
| 1q22 | rs760077 | chr1:155178782 | T | 0.857 | 0.137 | 0.016 | 1.146 | 1.111-1.183 | $1.14 \times 10^{-17}$ | MTX1 | Missense |
| 3q13.2 | rs72943992 | chr3:112403577 | C | 0.629 | 0.099 | 0.012 | 1.104 | 1.079-1.130 | $3.41 \times 10^{-17}$ | CCDC80 | Intergenic |
| 6p22.1 | rs9260603 | chr6:29922754 | C | 0.592 | 0.088 | 0.012 | 1.092 | 1.067-1.116 | $2.29 \times 10^{-14}$ | HLA-A | Intergenic |
| 6p21.3 | rs9270664 | chr6:32566149 | A | 0.497 | 0.091 | 0.011 | 1.095 | 1.071-1.120 | $6.35 \times 10^{-16}$ | HLA-DRB1 | Intergenic |
| 6p21.1 | rs62396739 | chr6:41730412 | A | 0.267 | 0.104 | 0.013 | 1.11 | 1.083-1.138 | $1.51 \times 10^{-16}$ | FRS3 | Intergenic |
| 7q21.3 | rs11531574 | chr7:97827833 | T | 0.278 | 0.073 | 0.013 | 1.075 | 1.048-1.103 | $2.16 \times 10^{-8}$ | LMTK2 | Intron |
| 8q24.3 | rs2920279 | chr8:143762135 | C | 0.645 | 0.201 | 0.012 | 1.223 | 1.195-1.251 | $1.95 \times 10^{-65}$ | PSCA | Intron |
| 9q34.2 | rs9411372 | chr9:136134068 | A | 0.214 | 0.089 | 0.014 | 1.094 | 1.064-1.124 | $2.42 \times 10^{-10}$ | ABO | Intron |
| 20p12.3 | rs6038583 | chr20:6670473 | G | 0.247 | 0.09 | 0.013 | 1.095 | 1.067-1.123 | $3.32 \times 10^{-12}$ | BMP2 | Intergenic |

The Japanese overall AG meta-analysis excluded BBJ to avoid sample overlap with the BBJ gastric cancer GWAS. This BBJ-excluded summary-statistics dataset was used in LDSC, MTAG, LAVA, and Genomic SEM analyses of AG and gastric cancer. Regional lead variants reaching $P < 5 \times 10^{-8}$ are shown. Beta and SE are reported for the displayed effect allele; gene labels indicate nearby genes and do not imply causality. Positions are based on GRCh37. AG, atrophic gastritis; BBJ, BioBank Japan; CI, confidence interval; EAF, effect allele frequency; OR, odds ratio; SE, standard error.

**Supplementary Table 13. SNP heritability and genome-wide genetic correlations between AG/SAG phenotypes and gastric cancer estimated by LDSC.**

| Trait pair | AG/SAG summary statistics | | | GC summary statistics | | | cross trait intercept | genetic correlation | SE | Unadjusted P value | Bonferroni-adjusted P-value |
|---|---|---|---|---|---|---|---|---|---|---|---|
| | N | heritability | intercept | N | heritability | intercept | | | | | |
| AG vs GC | 125,038 | 0.031 | 1.034 | 167,122 | 0.0201 | 0.9698 | -0.006 | 0.529 | 0.137 | $1.00 \times 10^{-4}$ | $6.00 \times 10^{-4}$ |
| SAG vs GC | 125,038 | 0.018 | 1.007 | | | | -0.004 | 0.436 | 0.183 | 0.018 | 0.105 |
| Seronegative AG vs GC | 67,120 | 0.019 | 0.990 | | | | 0.003 | 0.079 | 0.178 | 0.656 | 1.000 |
| Seronegative SAG vs GC | 66,780 | 0.025 | 0.980 | | | | 0.006 | -0.145 | 0.157 | 0.355 | 1.000 |
| Seropositive AG vs GC | 57,918 | 0.060 | 1.028 | | | | -0.007 | 0.447 | 0.162 | $5.60 \times 10^{-3}$ | 0.034 |
| Seropositive SAG vs GC | 57,918 | 0.035 | 0.997 | | | | -0.006 | 0.492 | 0.207 | 0.018 | 0.106 |

SNP heritability, univariate LDSC intercepts, cross-trait LDSC intercepts, and genome-wide genetic correlations were estimated using LD scores derived from the 1000 Genomes Project Phase 3 East Asian reference panel. AG/SAG summary statistics excluded BBJ to avoid sample overlap with the BBJ gastric cancer GWAS. The same gastric cancer summary statistics (N = 167,122) were used for all trait pairs. The univariate intercept quantifies inflation within each GWAS, whereas the cross-trait intercept captures covariance attributable to sample overlap or correlated confounding across the two GWASs. Unadjusted P values test the null hypothesis that the genetic correlation equals zero. AG, atrophic gastritis; BBJ, BioBank Japan; GC, gastric cancer; LDSC, linkage disequilibrium score regression; SAG, severe atrophic gastritis; SE, standard error.

Adjusted P-values use Bonferroni correction across six trait pairs.

### Supplementary Table 14. Summary of pairwise MTAG analyses of AG/SAG phenotypes and gastric cancer.

| Phenotype | Maximum N | GWAS mean $\chi^2$ | MTAG mean $\chi^2$ | GWAS-equivalent maximum N | Relative N gain, % | Omega correlation |
|---|---|---|---|---|---|---|
| AG | 125,038 | 1.081 | 1.112 | 173,632 | 38.9 | 0.694 |
| SAG | 125,038 | 1.043 | 1.059 | 171,034 | 36.8 | 0.514 |
| Seronegative AG | 67,120 | 1.032 | 1.032 | 67,180 | 0.1 | NA |
| Seronegative SAG | 66,780 | 1.063 | 1.063 | 66,851 | 0.1 | NA |
| Seropositive AG | 57,918 | 1.076 | 1.101 | 76,802 | 32.6 | 0.593 |
| Seropositive SAG | 57,918 | 1.038 | 1.058 | 88,081 | 52.1 | 0.586 |

Each of the six AG/SAG phenotypes was analyzed jointly with BBJ gastric cancer using MTAG. BBJ was excluded from the overall AG input to avoid sample overlap. Maximum N denotes the maximum SNP-level sample size in the input summary statistics. GWAS-equivalent maximum N is an estimated equivalent sample size, not an observed participant count. Relative N gain was calculated as 100 × (GWAS-equivalent maximum N / maximum N − 1). Omega correlation is the correlation implied by the estimated MTAG Omega matrix and is distinct from the LDSC genetic correlation. NA indicates that the Omega correlation was not estimable. AG, atrophic gastritis; BBJ, BioBank Japan; GC, gastric cancer; GWAS, genome-wide association study; MTAG, multi-trait analysis of GWAS; SAG, severe atrophic gastritis.

### Supplementary Table 15. Genome-wide significant loci in MTAG-adjusted AG and gastric cancer summary statistics.

| Analysis | Locus | Lead SNP | Position (GRCh37) | Effect allele | EAF | Beta | SE | P-value | Nearest Gene | Annotation |
|---|---|---|---|---|---|---|---|---|---|---|
| MTAG-adjusted AG | 1p36.33 | rs58980341 | chr1:1135107 | T | 0.137 | 0.025 | 0.004 | $3.89 \times 10^{-8}$ | TTLL10 | Intergenic |
| | 1q21.3 | rs12030974 | chr1:151798295 | A | 0.395 | 0.025 | 0.004 | $3.22 \times 10^{-12}$ | RORC | Intron |
| | 1q22 | rs140081212 | chr1:155184975 | G | 0.767 | 0.083 | 0.007 | $9.29 \times 10^{-35}$ | GBAP1 | Exon |
| | 3q13.2 | rs72943992 | chr3:112403577 | C | 0.616 | 0.031 | 0.004 | $1.15 \times 10^{-18}$ | CCDC80 | Intergenic |
| | 5p13.1 | rs10078575 | chr5:40704074 | A | 0.447 | 0.054 | 0.006 | $3.59 \times 10^{-21}$ | PTGER4 | Intergenic |
| | 6p21.3 | rs1130153 | chr6:32610947 | A | 0.954 | 0.068 | 0.007 | $1.15 \times 10^{-20}$ | HLA-DQA1 | Intron |
| | 6p21.1 | rs62396739 | chr6:41730412 | A | 0.196 | 0.027 | 0.004 | $5.53 \times 10^{-12}$ | FRS3 | Intergenic |
| | 8q24.3 | rs2920279 | chr8:143762135 | C | 0.342 | 0.074 | 0.004 | $2.93 \times 10^{-94}$ | PSCA | Intron |
| | 9q34.2 | rs2519093 | chr9:136141870 | T | 0.191 | 0.029 | 0.004 | $9.66 \times 10^{-14}$ | ABO | Intron |
| | 20p12.3 | rs6038583 | chr20:6670473 | G | 0.251 | 0.023 | 0.004 | $1.22 \times 10^{-8}$ | BMP2 | Intergenic |
| MTAG-adjusted GC | 1q21.3 | rs12030974 | chr1:151798295 | A | 0.395 | 0.019 | 0.003 | $1.05 \times 10^{-10}$ | RORC | Intron |
| | 1q22 | rs140081212 | chr1:155184975 | G | 0.767 | 0.060 | 0.004 | $1.83 \times 10^{-42}$ | GBAP1 | Exon |
| | 3q13.2 | rs72943992 | chr3:112403577 | C | 0.616 | 0.022 | 0.003 | $4.63 \times 10^{-14}$ | CCDC80 | Intergenic |
| | 5p13.1 | rs59585832 | chr5:40744038 | C | 0.447 | 0.031 | 0.003 | $1.45 \times 10^{-28}$ | TTC33 | Intron |
| | 6p22.3 | rs7752780 | chr6:20666022 | G | 0.599 | 0.017 | 0.003 | $4.88 \times 10^{-9}$ | CDKAL1 | Intron |
| | 6p21.3 | rs1130153 | chr6:32610947 | A | 0.954 | 0.048 | 0.005 | $3.99 \times 10^{-18}$ | HLA-DQA1 | Intron |
| | 8q24.3 | rs2976397 | chr8:143764613 | T | 0.226 | 0.056 | 0.003 | $6.70 \times 10^{-89}$ | PSCA | 470 bp downstream |
| | 9q34.2 | rs2519093 | chr9:136141870 | T | 0.191 | 0.023 | 0.003 | $3.42 \times 10^{-13}$ | ABO | Intron |

MTAG jointly analyzed harmonized BBJ-excluded Japanese AG summary statistics and BBJ gastric cancer summary statistics. The table shows trait-specific MTAG-adjusted regional lead variants at loci reaching $P < 5 \times 10^{-8}$. Positions are based on GRCh37. Regional lead variants can differ between the original GWAS and MTAG-adjusted analyses; locus-level changes in significance should therefore be interpreted using both the variant identifier and P value. EAFs were obtained from the 1000 Genomes Project Phase 3 East Asian reference population. AG, atrophic gastritis; BBJ, BioBank Japan; CI, confidence interval; EAF, effect allele frequency; GC, gastric cancer; MTAG, multi-trait analysis of GWAS; OR, odds ratio; SE, standard error.

### Supplementary Table 16. Significant local genetic correlations between AG and gastric cancer estimated by LAVA.

#### A. Non-MHC regions significant in the genome-wide analysis (218 tests)

| Locus / region | Position (GRCh37) | Region size (bp) | Representative genes | No. of SNPs | Local h² (AG) | Local h² (GC) | Local genetic correlation (rho) | 95% CI | P-value | Bonferroni-adjusted P-value |
|---|---|---|---|---|---|---|---|---|---|---|
| 8q24.3 | chr8:143611462-144934288 | 1,322,826 | *PSCA* | 1,369 | $5.34 \times 10^{-3}$ | $3.05 \times 10^{-3}$ | 0.937 | 0.876–0.999 | $4.30 \times 10^{-64}$ | $9.37 \times 10^{-62}$ |
| 1q22 | chr1:154685546-156813845 | 2,128,299 | *MUC1/MTX1* | 851 | $1.97 \times 10^{-3}$ | $3.15 \times 10^{-3}$ | 1.000* | 0.907–1.000 | $2.84 \times 10^{-32}$ | $6.19 \times 10^{-30}$ |
| 3p21.31 | chr3:45844192-47588461 | 1,744,269 | *LZTFL1/CCR1/CCR2/CCR5/SETD2* | 1,765 | $9.38 \times 10^{-4}$ | $2.05 \times 10^{-4}$ | 1.000* | 0.498–1.000 | $8.48 \times 10^{-6}$ | $1.85 \times 10^{-3}$ |
| 5p13.1 | chr5:40290852-41279590 | 988,738 | *PTGER4/PRKAA1/CARD6* | 1,869 | $4.32 \times 10^{-4}$ | $1.13 \times 10^{-3}$ | 0.844 | 0.489–1.000 | $2.26 \times 10^{-5}$ | $4.93 \times 10^{-3}$ |
| 8q24.3 | chr8:142611890-143611461 | 999,571 | *ADGRB1/TSNARE1* | 1,136 | $7.24 \times 10^{-4}$ | $2.88 \times 10^{-4}$ | 0.853 | 0.465–1.000 | $1.42 \times 10^{-4}$ | 0.031 |

#### B. MHC regional analysis (3 tests)

| Locus / region | Position (GRCh37) | Region size (bp) | Representative genes | No. of SNPs | Local h² (AG) | Local h² (GC) | Local genetic correlation (rho) | 95% CI | P-value | Bonferroni-adjusted P-value |
|---|---|---|---|---|---|---|---|---|---|---|
| MHC class I | chr6:29500000-31498002 | 1,998,002 | — | 8,663 | $2.40 \times 10^{-3}$ | $1.90 \times 10^{-3}$ | 0.819 | 0.681–0.963 | $3.23 \times 10^{-19}$ | $9.69 \times 10^{-19}$ |
| MHC class III | chr6:31498003-32191844 | 693,841 | — | 615 | $2.01 \times 10^{-3}$ | $9.30 \times 10^{-4}$ | 0.615 | 0.418–0.821 | $1.33 \times 10^{-7}$ | $4.00 \times 10^{-7}$ |
| MHC class II | chr6:32191845-33500000 | 1,308,155 | — | 8,294 | $3.22 \times 10^{-3}$ | $1.67 \times 10^{-3}$ | 0.689 | 0.547–0.843 | $2.06 \times 10^{-15}$ | $6.18 \times 10^{-15}$ |

Local genetic correlations were estimated between BBJ-excluded Japanese AG and BBJ gastric cancer using LAVA and the 1000 Genomes Project Phase 3 East Asian LD reference. Regions with sufficient univariate local SNP heritability in both traits were tested. Panel A shows non-MHC regions significant after Bonferroni correction across 218 eligible regions ($P < 0.05/218 = 2.29 \times 10^{-4}$). Panel B shows post hoc MHC regional analyses, corrected separately across three tests ($P < 0.05/3 = 0.0167$). Asterisks indicate correlation estimates capped at 1. Confidence intervals reaching the parameter boundary of 1 should be interpreted cautiously. Gene labels indicate nearby genes and do not imply causality. AG, atrophic gastritis; BBJ, BioBank Japan; CI, confidence interval; GC, gastric cancer; LD, linkage disequilibrium; LAVA, Local Analysis of [co]Variant Association; MHC, major histocompatibility complex; rho, local genetic correlation.

### Supplementary Table 17. Genome-wide significant loci in the AG-anchored shared and GC-specific Genomic SEM components.

| Analysis | Locus | Lead SNP | Position (GRCh37) | Effect allele | EAF | Component beta | SE | P-value | Nearest Gene | Annotation |
|---|---|---|---|---|---|---|---|---|---|---|
| AG-GC shared component | 1q21.3 | rs939595 | chr1:151783688 | A | 0.298 | -0.111 | 0.020 | $3.36 \times 10^{-8}$ | RORC | Intronic |
| | 1q22 | rs2075571 | chr1:155174106 | T | 0.184 | 0.150 | 0.021 | $5.60 \times 10^{-13}$ | THBS3 | Intronic |
| | 3q13.2 | rs1249910 | chr3:112391174 | A | 0.41 | -0.128 | 0.016 | $4.98 \times 10^{-15}$ | CCDC80 | Intergenic |
| | 6p21.32 | rs17843604 | chr6:32620283 | T | 0.497 | 0.118 | 0.016 | $2.32 \times 10^{-13}$ | HLA-DQA1 | Intergenic |
| | 6p21.32 | rs86715 | chr6:33480435 | G | 0.093 | -0.162 | 0.028 | $1.11 \times 10^{-8}$ | ZBTB9 | Intergenic |
| | 6p21.1 | rs11753329 | chr6:41788380 | G | 0.311 | 0.130 | 0.017 | $7.64 \times 10^{-14}$ | USP49 | Intronic |
| | 7q21.3 | rs3735254 | chr7:97838247 | G | 0.286 | 0.100 | 0.018 | $4.53 \times 10^{-8}$ | LMTK2 | 3' UTR |
| | 8q24.3 | rs2976394 | chr8:143763622 | T | 0.367 | 0.280 | 0.017 | $5.21 \times 10^{-63}$ | PSCA | 3' UTR |
| | 20p12.3 | rs6054471 | chr20:6674887 | T | 0.246 | 0.127 | 0.019 | $9.67 \times 10^{-12}$ | BMP2 | Intergenic |
| GC-specific component | 1q22 | rs2075571 | chr1:155174106 | T | 0.184 | 0.166 | 0.026 | $1.13 \times 10^{-10}$ | THBS3 | Intronic |
| | 5p13.1 | rs13361707 | chr5:40791884 | T | 0.42 | -0.136 | 0.020 | $1.72 \times 10^{-11}$ | PRKAA1 | Intronic |

The two-trait Genomic SEM model used BBJ-excluded AG and BBJ gastric cancer summary statistics. AG was the anchor trait: its loading on the shared factor F was fixed to 1 and its residual genetic variance was constrained to 0. The shared component therefore represents SNP association with an AG-anchored factor and is not a symmetric intersection of AG and gastric cancer. The GC-specific component represents residual gastric cancer association after accounting for F and does not imply complete absence of AG-related effects. Regional lead variants reaching $P < 5 \times 10^{-8}$ are shown. Positions are based on GRCh37. AG, atrophic gastritis; BBJ, BioBank Japan; CI, confidence interval; GC, gastric cancer; OR, odds ratio; SEM, structural equation modeling; SE, standard error.

**Supplementary Table 18. Genome-wide significant loci from cross-ancestry meta-analyses integrating Japanese AG/SAG and external chronic gastritis GWAS.**

| Analysis | Locus | Lead SNP | Position (GRCh37) | Effect allele | EAF | Beta_0 | SE_0 | P-value | Heterogeneity P | Nearest Gene | Annotation |
|---|---|---|---|---|---|---|---|---|---|---|---|
| Japanese AG + cross-ancestry chronic gastritis meta-analysis | 1p36.33 | rs58980341 | chr1:1135107 | T | 0.086577 | 0.07367 | 0.008455 | $1.22 \times 10^{-10}$ | $3.22 \times 10^{-11}$ | TTLL10 | Intergenic |
| | 1q22 | rs760077 | chr1:155178782 | T | 0.663944 | 0.096005 | 0.014326 | $5.11 \times 10^{-16}$ | $1.34 \times 10^{-16}$ | MTX1 | Exonic |
| | 3q13.2 | rs72943992 | chr3:112403577 | T | 0.095256 | -0.074148 | 0.007907 | $4.10 \times 10^{-17}$ | $1.39 \times 10^{-17}$ | CCDC80 | Intergenic |
| | 6p21.3 | rs1130153 | chr6:32610947 | C | 0.061539 | -0.15452 | 0.016697 | $1.34 \times 10^{-15}$ | $3.43 \times 10^{-16}$ | HLA-DQA1 | Intron |
| | 6p21.1 | rs62396739 | chr6:41730412 | A | 0.164829 | 0.074996 | 0.009991 | $2.62 \times 10^{-15}$ | $6.66 \times 10^{-16}$ | FRS3 | Intergenic |
| | 8q24.3 | rs2920279 | chr8:143762135 | C | 0.483809 | 0.141521 | 0.012021 | $3.86 \times 10^{-60}$ | $6.94 \times 10^{-18}$ | PSCA | Intron |
| | 9q34.2 | rs9411372 | chr9:136134068 | A | 0.220828 | -0.162359 | 0.405781 | $3.58 \times 10^{-8}$ | 0.303 | ABO | Intron |
| | 20p12.3 | rs6038583 | chr20:6670473 | A | 0.723955 | -0.066331 | 0.012606 | $7.65 \times 10^{-12}$ | $2.92 \times 10^{-12}$ | BMP2 | Intergenic |
| Japanese SAG + cross-ancestry chronic gastritis meta-analysis | 1p13.2 | rs2476601 | chr1:114377568 | A | 0.106988 | 1.64883 | 0.390397 | $3.59 \times 10^{-9}$ | $4.81 \times 10^{-10}$ | PTPN22 | Exonic |
| | 1q22 | rs760077 | chr1:155178782 | T | 0.647581 | 0.164931 | 0.019967 | $6.79 \times 10^{-16}$ | $1.91 \times 10^{-16}$ | MTX1 | Exonic |
| | 5p13.1 | rs28540420 | chr5:40687463 | T | 0.254152 | -0.09073 | 0.014718 | $1.00 \times 10^{-9}$ | $1.37 \times 10^{-10}$ | PTGER4 | Intron |
| | 6p22.1 | rs3129030 | chr6:29661695 | A | 0.237243 | 0.132205 | 0.019739 | $3.29 \times 10^{-9}$ | $1.11 \times 10^{-9}$ | ZFP57 | Intergenic |
| | 6p21.3 | rs34004019 | chr6:32626403 | G | 0.318444 | -0.110396 | 0.018486 | $1.39 \times 10^{-15}$ | 0.018 | HLA-DQB1 | Downstream |
| | 6q21 | rs4946699 | chr6:106263758 | G | 0.832744 | -2.53584 | 1.26462 | $3.22 \times 10^{-8}$ | 0.093 | PRDM1 | Intergenic |
| | 8q24.3 | rs2920280 | chr8:143761144 | C | 0.434005 | 0.243805 | 0.014646 | $1.00 \times 10^{-66}$ | $<1.00 \times 10^{-16}$ | PSCA | Intron |
| Japanese HP-positive AG + cross-ancestry chronic gastritis meta-analysis | 1q22 | rs760077 | chr1:155178782 | T | 0.630667 | 0.168068 | 0.015651 | $1.18 \times 10^{-33}$ | $6.94 \times 10^{-18}$ | MTX1 | Exonic |
| | 5q35.2 | rs359456 | chr5:173279808 | C | 0.557068 | 0.064449 | 0.010911 | $2.55 \times 10^{-10}$ | $3.85 \times 10^{-11}$ | CPEB4 | Intergenic |
| | 6p22.2 | rs3734533 | chr6:26240624 | G | 0.172671 | -0.090544 | 0.013352 | $2.63 \times 10^{-8}$ | $1.56 \times 10^{-7}$ | HIST1H4F | 5' UTR |
| | 6p21.3 | rs12614 | chr6:31914179 | T | 0.124355 | -0.183385 | 0.028921 | $6.44 \times 10^{-18}$ | $<1.00 \times 10^{-16}$ | CFB | Exonic |
| | 6p21.1 | rs62396739 | chr6:41730412 | A | 0.154055 | 0.084432 | 0.011723 | $6.76 \times 10^{-14}$ | $1.72 \times 10^{-14}$ | FRS3 | Intergenic |
| | 8q24.3 | rs2920279 | chr8:143762135 | C | 0.467214 | 0.216567 | 0.010127 | $9.28 \times 10^{-100}$ | $<1.00 \times 10^{-16}$ | PSCA | Intron |
| | 9q34.2 | rs8176645 | chr9:136149098 | A | 0.370803 | 0.064895 | 0.013542 | $1.25 \times 10^{-11}$ | $2.16 \times 10^{-11}$ | ABO | Intron |
| | 20p12.3 | rs6038583 | chr20:6670473 | A | 0.721783 | -0.091319 | 0.013554 | $2.16 \times 10^{-15}$ | $7.13 \times 10^{-16}$ | BMP2 | Intergenic |
| Japanese HP-positive SAG + cross-ancestry chronic gastritis meta-analysis | 1p13.2 | rs2476601 | chr1:114377568 | A | 0.107128 | 1.69314 | 0.413815 | $4.11 \times 10^{-9}$ | $5.53 \times 10^{-10}$ | PTPN22 | Exonic |
| | 1q22 | rs760077 | chr1:155178782 | T | 0.630667 | 0.23537 | 0.023248 | $4.49 \times 10^{-26}$ | $<1.00 \times 10^{-16}$ | MTX1 | Exonic |
| | 5p13.1 | rs28540420 | chr5:40687463 | T | 0.238731 | -0.092438 | 0.019586 | $1.55 \times 10^{-8}$ | $2.26 \times 10^{-9}$ | PTGER4 | Intron |
| | 6p21.3 | 6:32603563:C:G | chr6:32603563 | G | 0.280943 | -0.077348 | 0.020525 | $1.50 \times 10^{-9}$ | 0.660 | HLA-DQA1 | Intron |
| | 8q24.3 | rs2920280 | chr8:143761144 | C | 0.430481 | 0.290811 | 0.01747 | $4.59 \times 10^{-79}$ | $<1.00 \times 10^{-16}$ | PSCA | Intron |
| Japanese HP-negative AG + cross-ancestry chronic gastritis meta-analysis | 1p13.2 | rs2476601 | chr1:114377568 | A | 0.107113 | 1.71669 | 0.31915 | $1.24 \times 10^{-9}$ | $1.62 \times 10^{-10}$ | PTPN22 | Exonic |
| | 6p21.3 | rs9271065 | chr6:32575619 | G | 0.8676 | -0.445562 | 0.041094 | $4.37 \times 10^{-19}$ | $<1.00 \times 10^{-16}$ | HLA-DRB1 | Intergenic |
| Japanese HP-negative SAG + cross-ancestry chronic gastritis meta-analysis | 1p13.2 | rs2476601 | chr1:114377568 | A | 0.107113 | 1.69549 | 0.321206 | $1.26 \times 10^{-9}$ | $1.64 \times 10^{-10}$ | PTPN22 | Exonic |
| | 2q24.2 | rs11679244 | chr2:163082395 | A | 0.418791 | 0.309238 | 0.045181 | $1.63 \times 10^{-8}$ | $7.59 \times 10^{-9}$ | FAP | Intron |
| | 6p22.1 | 6:29692622:C:G | chr6:29692622 | G | 0.137671 | -0.334267 | 0.041598 | $3.26 \times 10^{-11}$ | $3.70 \times 10^{-12}$ | HLA-F | Intron |
| | 6p21.3 | rs9269047 | chr6:32438783 | G | 0.861603 | -0.631245 | 0.046489 | $1.52 \times 10^{-22}$ | $<1.00 \times 10^{-16}$ | HLA-DRA | Intergenic |
| | 6q27 | rs204295 | chr6:167500562 | T | 0.470416 | -0.220588 | 0.035277 | $1.19 \times 10^{-8}$ | $7.35 \times 10^{-9}$ | CCR6 | Intergenic |
| | 19q13.12 | rs725986 | chr19:36057622 | C | 0.151306 | -0.335212 | 0.049366 | $4.41 \times 10^{-9}$ | $7.59 \times 10^{-10}$ | ATP4A | Intergenic |
| | 22q11.1 | rs2186377 | chr22:17128707 | G | 0.019665 | 1.31599 | 0.259295 | $3.79 \times 10^{-8}$ | 0.064 | TPTEP1 | Intron |

MR-MEGA integrated each Japanese AG/SAG GWAS with chronic gastritis GWAS summary statistics from FinnGen, UK Biobank, and European and African American or Afro-Caribbean MVP datasets. Beta_0 and SE_0 are the estimated intercept effect and its standard error from the meta-regression; the P-value column reports the association P value. Heterogeneity P is the PH field reported in the source MR-MEGA output. EAF is the effect-allele frequency reported in the harmonized source data. Anti-*H. pylori* IgG-seropositive and seronegative strata were defined as >3 U/mL and <=3 U/mL, respectively. Because the external phenotype was chronic gastritis rather than serologically defined AG/SAG, these loci are supportive cross-ancestry findings and are not assigned the same evidence tier as the 17 Japanese loci. Positions are based on GRCh37. AG, atrophic gastritis; EAF, effect allele frequency; MVP, Million Veteran Program; SAG, severe atrophic gastritis; SE, standard error.
